# Multimodal gene prioritization reveals nonlinear regulatory architecture in childhood-onset asthma

**DOI:** 10.64898/2026.07.14.26357983

**Authors:** Nan Huang, Michelle F. Ragsac, Xiaoyu Gui, Kelan G. Tantisira, Tiffany Amariuta

## Abstract

Asthma is a heritable complex disease that disproportionately burdens minority and admixed populations in the US. However, the causal genes and regulatory mechanisms governing inherited risk remain largely unresolved. We performed a European-ancestry meta-analysis of 141,894 cases and 1,361,846 controls drawn from the Trans-national Asthma Genetic Consortium (TAGC) and Global Biobank Meta-analysis Initiative (GBMI), yielding an estimated *h^2^_SNP_* of 0.056 (SE = 0.0038) and 275 independently associated loci. To enhance mechanistic inference beyond variant-level associations, we developed a multimodal framework to predict asthma risk integrating GWAS summary statistics, bulk tissue expression quantitative trait loci (eQTL) data from the Genotype-Tissue Expression (GTEx) project, and single-cell gene eQTL data from the OneK1K Project. We performed transcriptome-wide association studies (TWAS) and subsequently applied probabilistic fine-mapping with FOCUS to prioritize putative causal genes expressed in bulk tissues and higher resolution immune cell populations. Fine-mapping asthma-associated genes implicated barrier-immune and metabolic-endocrine tissues alongside adaptive T-cell subsets as the primary mediators of asthma genetic risk, resolving canonical CD4+ Th2 effector genes including *IL1RL1*, *TSLP*, *STAT6*, and *GATA3*. Using these prioritized genes, we constructed a polygenic transcriptome risk score (PTRS) using random forest to integrate gene-level effects across critical tissues and cell types. Evaluated in two ancestrally distinct pediatric asthma cohorts, the Childhood Asthma Management Program (CAMP) and the Genetics of Asthma in Costa Rica Study (GACRS), our PTRS demonstrated improved transferability over the standard variant-level and gene-level baseline models. While modest common variant heritability limits the discriminative power of our models, we estimated a theoretical maximum achievable area under the receiver operating characteristic (AUROC) curve of 0.64. Our integrative nonlinear model of PRS-CSx and cross-modal (bulk tissue and single cell) FOCUS PTRS resulted in the best cross-cohort performance (CAMP AUC = 0.632, sd = 0.04, 3.55 case/control odds ratio in top vs. bottom quartiles), representing an increase of +0.118 AUC over PRS-CSx, +0.067 AUC over tissue-specific TWAS pruning and thresholding, and +0.041 AUC over cell-type-specific FOCUS PTRS. Our results demonstrate that modeling nonlinear interactions between variant- and gene-level effects across both bulk tissue and single cell eQTL data improves our ability to determine high-risk individuals and to explain the likely mechanisms driving genetic susceptibility of childhood-onset asthma.

**Highlights:**

- Multimodal putative causal gene prioritization integrates GWAS summary statistics, bulk tissue, and single-cell expression data to resolve effector genes underlying childhood-onset asthma.
- Gene-level fine-mapping of underlying asthma GWAS loci reveals CD4+ Th2 effector genes, which are known drivers of eosinophilic airway inflammation, across multiple tissue and cell types including CD4+ cytotoxic T cells, naïve CD4+ T cells, and natural killer cells.
- We constructed and evaluated variant-level multi-ancestry asthma PRS and gene-level PTRS for genes in fine-mapped credible sets.
- We identified tissues and cell types that consistently helped discriminate asthma cases from controls, namely esophagus mucosa and CD4+ naïve T cells.
- We found that nonlinear modeling of both variant-level and gene-level effects across top tissues and cell types, tested across various integration strategies, substantially outperformed linear models in the distinction of asthma cases from controls.
- The model that most effectively distinguished cases from controls was a cross-modal model of PRS-CSx combined with the esophagus mucosa and CD4+ naïve T cell PTRS models, resulting in an AUC = 0.632 ± 0.040 and case/control enrichment odds ratio of 3.55 in top/bottom quartiles.
- Overall modest discrimination between cases and controls with genetic predictors supports asthma as a complex disease with a substantial non-genetic component.

**Graphical Abstract:** 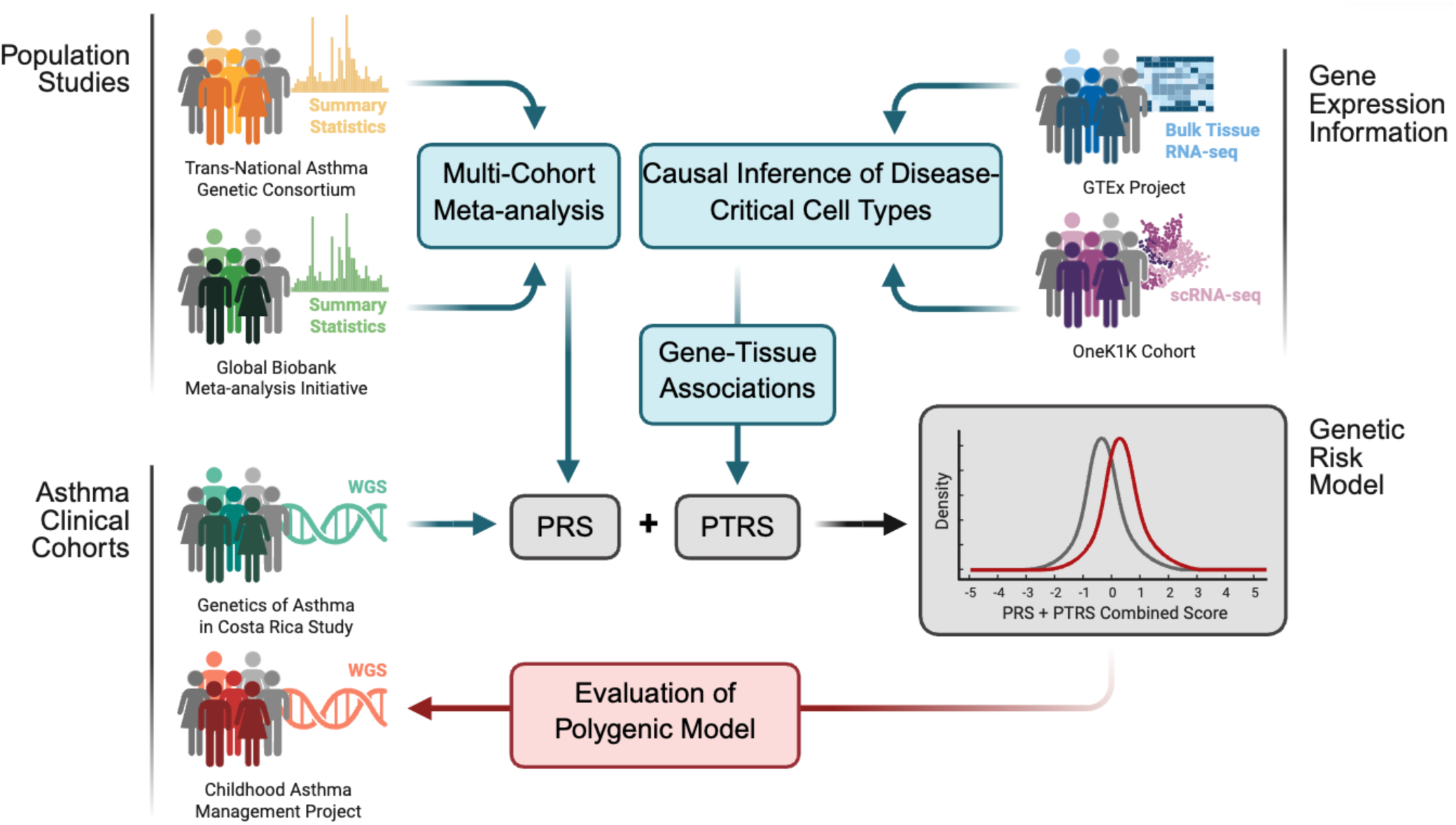

## Introduction

Asthma is a complex, heterogeneous respiratory disease encompassing multiple molecular endotypes, characterized by chronic airway inflammation, bronchial hyperresponsiveness, and variable airflow obstruction. Affected individuals present with recurrent respiratory symptoms that can fluctuate in frequency and severity, including wheezing, shortness of breath, chest tightness, and cough^1–3^. Asthma is classified as a major noncommunicable disease by the World Health Organization, and in 2021, the disease affected an estimated 260 million individuals worldwide^4^, with 24.9 million in the United States^5^. Although asthma affects individuals of all ages, onset most commonly occurs during childhood, coinciding with critical periods of immune and pulmonary development during which genetic susceptibility and environmental exposures may have lasting effects on respiratory health^1,6–8^. In the United States, however, this burden is not distributed equally: children from historically underrepresented populations, particularly Latino, African American and Caribbean American groups, experience higher asthma prevalence, increased disease severity, and elevated hospitalization risk, reflecting the disproportionate influence of socio-environmental factors and structural determinants of health^5,6,8^. These disparities are further compounded by persistent underrepresentation of non-European and admixed American populations in genomic research, limiting the generalizability of existing genetic findings and equitable development of precision medicine approaches for childhood asthma.

Alongside socio-environmental influences, asthma has a strong genetic component: twin studies estimate asthma heritability between 35% and 75%, with childhood-onset disease showing a stronger heritable component relative to adult-onset disease^1,9–11^. Large-scale genome-wide association studies (GWAS) have also identified hundreds of loci associated with asthma and other allergic traits^12–14^. Among these, variants at the *17q12-21* locus harboring *ORMDL3* and *GSDMB* represent the most widely replicated asthma association across diverse populations, with particularly strong effects in childhood-onset disease^15,16^. Other consistently replicated loci include variants near *IL33*, *IL1RL1, IL1RL2, IL18R1*, SLC25A46, *TSLP*, *IL13, RAD50*, *HLA-DR*, and *HLA-DQ*, implicating immune regulation, epithelial barrier integrity, and airway inflammation as central pathogenic mechanisms^13,15^. Despite this progress, individually identified variants such as single nucleotide polymorphisms (SNPs) carry small effect sizes and predominantly reside in noncoding regulatory regions whose regulatory targets, relevant cell types, and downstream effector genes remain largely unknown^17,18^. This interpretability gap reflects a fundamental limitation of variant-level association mapping: without resolving which genes are causally dysregulated in disease, in which cell types, and in which regulatory mechanisms, associated loci cannot be linked to tractable biological hypotheses or directly inform the development of mechanistically grounded interventions or treatments for childhood-onset asthma^19^. This interpretability gap is further exacerbated by the fact that the majority of GWAS have been conducted primarily in individuals of European ancestry, limiting the portability of their findings to the populations most severely burdened by disease^20,21^. Together, these limitations underscore the critical need for integrative, biologically grounded approaches that move beyond variant-level associations to resolve the causal genes, cell types, and regulatory mechanisms underlying disease, particularly in ancestrally diverse populations.

Resolving this biological complexity requires frameworks that operate at multiple resolutions of genetic and regulatory factors. Polygenic risk scores (PRS) capture the contribution of common variants to disease by aggregating the effects of GWAS loci to quantify inherited susceptibility to complex diseases such as asthma^22–27^. Furthermore, Bayesian approaches such as PRS-CS and PRS- CSx have improved PRS cross-ancestry portability by applying shrinkage priors on SNP effect sizes^28,29^. However, variant-level scores alone cannot identify which genes are dysregulated in which cell types, limiting biological interpretability despite improved predictive performance.

Transcriptome-wide association studies (TWAS) partially address this by linking predicted tissue- specific gene expression to trait variation, providing a gene-level resolution layer that variant- level scores lack^30,31^. Polygenic transcriptome risk scores (PTRS) extend this framework by leveraging genetically predicted gene expression as a gene-based complement to traditional PRS, incorporating functional genomics information to improve the biological resolution and cross- ancestry portability of genetic risk scores^32^. However, because TWAS does not distinguish putative causal genes from co-regulated, or tagging, genes and tissues/cell types, the biological specificity and predictive utility of TWAS-derived features is limited^33^. Probabilistic fine-mapping frameworks such as FOCUS address this by computing posterior inclusion probabilities (PIPs) over candidate causal genes within credible sets, leveraging ancestry-specific differences in genetic correlation structure and gene expression regulation to enable more principled selection of putative causal genes at a given locus^34,35^.

Here, we present a multimodal framework that integrates variant-level PRS with gene-regulatory information from PTRS to resolve the putative causal genes, tissues, cell types, and regulatory mechanisms underlying childhood-asthma susceptibility. First, we establish training and validation cohorts using the Genetics of Asthma in Costa Rica Study (GACRS) cohort and the Childhood Asthma Management Program (CAMP)^36,37^. Then, we performed a GWAS meta-analysis of the two largest asthma biobanks, the Trans-National Asthma Genetic Consortium (TAGC) and the Global Biobank Meta-analysis Initiative (GBMI)^13,14^. We establish a variant-level PRS baseline using PRS-CS(x)^29^ and a gene-level baseline using the TWAS pruning and thresholding approach introduced by Liang and colleagues^32^. Then, we conduct TWAS using tissue-level eQTL data from GTEx and pseudobulked single cell eQTL data from OneK1K^38,39^, while specific immune cell populations may better capture disease-critical regulatory contexts than heterogeneous bulk tissues^40^. We hypothesized that because standard PTRS construction is susceptible to inflation from co-regulated, non-causal transcriptomic features^33^, we apply the probabilistic fine-mapping framework FOCUS^34,35^ to prioritize putative causal gene-tissue and gene-cell-type pairs. In our study, we evaluate different PTRS integration approaches including nonlinear classification models and cross-modal feature sets; we further assess the interpretability and concordance of variant-level and gene-level findings. We generally found that FOCUS-informed PTRS under various nonlinear training frameworks outperformed the baseline PRS and PTRS models and extracted features with greater biological plausibility. For example, using the GACRS cohort for training, the best tissue-level (esophagus mucosa) and cell-type-level (CD4+ naïve T) FOCUS PTRS outperformed the best tissue-level (spleen) and cell-type-level (NK 56^bright^) TWAS PTRS in distinguishing CAMP cases from controls, while also nominating tissues and cell types with more established roles in asthma pathogenesis. Our results suggest that fine-mapping asthma-associated gene loci may help remove tagging genes and tissues/cell types that attenuate cross-cohort generalizability. We subsequently constructed various cross-modal PTRS consisting of top-performing tissue-level and cell-type-level FOCUS PTRS and variant-level PRS-CS(x) evaluated across different nonlinear training architectures. Overall, our approach provides insight regarding the distinct disease risk captured in comparative models of (1) variant-level and gene-level effects, (2) tissue-level and cell-type-level gene effects, and (3) linear and nonlinear feature integration models.

## Results

### Cross-biobank asthma GWAS meta-analysis

To establish the variant-level genetic signal underlying asthma susceptibility, we performed a meta-analysis combining asthma GWAS summary statistics from two large cohorts: the Trans- National Asthma Genetic Consortium (TAGC) and the Global Biobank Meta-analysis Initiative (GBMI)^13,14^. Although neither consortium is restricted to pediatric cases, childhood-onset and adult-onset asthma share substantial common genetic architecture, particularly in established loci^8,12,13^. Our meta-analysis yielded a combined dataset of 141,894 cases and 1,361,846 controls (total N = 1,503,740). We identified 2,604 genome-wide significant variants (p ≤ 5 × 10^-8^) at 275 independent loci (**Figure 1A**, STAR★Methods, **Supplementary Table S1**). Among the independent loci, 264 represented noncoding regulatory regions, thus motivating further gene prioritization using transcriptome-wide association studies (TWAS)^30,31^ and subsequent fine-mapping.

**Figure 1.**
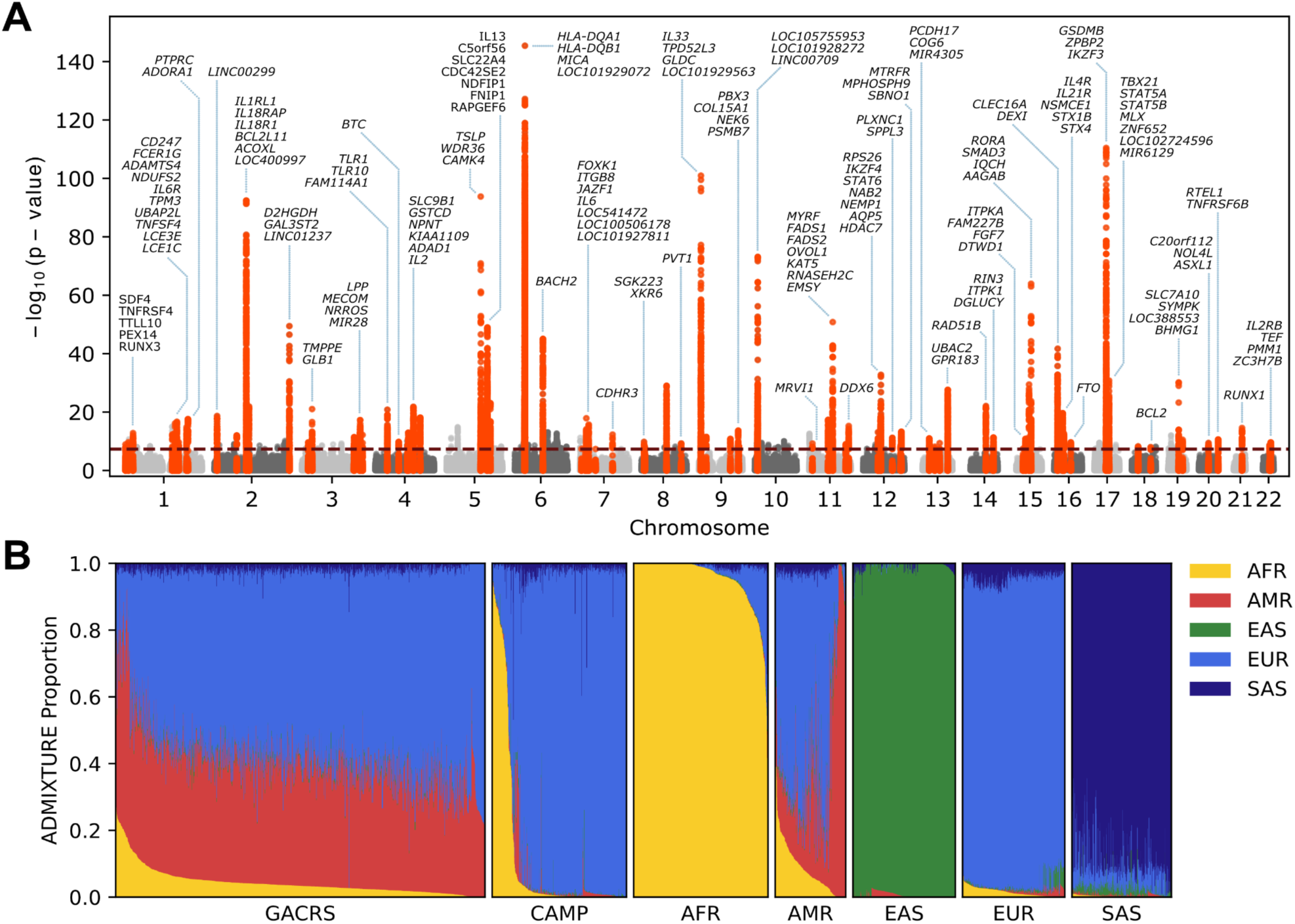
Genetic landscape of asthma and study cohort characterization. (**A**) GWAS meta-analysis of European-ancestry summary statistics from TAGC and GBMI. Each point represents a SNP and all SNPs within 500 kb of a genome-wide significant index variant (p ≤ 5 × 10^-8^) are highlighted in red, while all remaining SNPs are colored light or dark gray, alternating by chromosome. The dashed horizontal line indicates the genome-wide significance threshold. For each locus, up to three gene symbols are annotated, selected from the most significant variants within the 500 kb flanking window. Where adjacent loci fell within 500 kb of one another, annotations were merged into a single label to minimize crowding. Complete locus annotations are provided in **Supplementary Table S1**. (**B**) ADMIXTURE plots illustrating the estimated global ancestry proportions for each individual (along the x-axis) in the GACRS and CAMP cohorts, analyzed jointly with the 1000 Genomes Project (Phase 3) reference individuals. Known 1000 Genomes population labels were used to interpret the predicted ancestry components (*K*=5). The five rightmost panels (AFR, AMR, EAS, EUR, and SAS) represent the 1000 Genomes reference superpopulations. Each vertical bar represents one individual and colors are assigned based on the most representative feature of each superpopulation.

Restricting to pediatric asthma cases/controls in these GWAS would have resulted in notably smaller sample size (N = 27,375 for TAGC^13^ and N = 314,633 from the UK Biobank^41^. Therefore, we elected to maximize sample size and thus power to detect variant-level effects, which we hypothesized would better help identify asthma-associated genes and effector tissues/cell types through TWAS and gene-level fine-mapping. Furthermore, as both consortia consist of predominantly European-ancestry individuals, we restricted our meta-analysis to biobank- specific summary statistics of European participants to ensure compatibility with linkage disequilibrium (LD) panels required for downstream polygenic risk score (PRS) and previously constructed eQTL panels used for TWAS.

We anticipated that our increased GWAS sample size would improve power to detect loci of small effect and yield more precise SNP-heritability estimates, which directly informs PRS model evaluation by establishing the proportion of phenotypic variance explainable by common genetic variation^44,45^. We estimated the liability-scale SNP-heritability (*h*^2^) of asthma in our European- ancestry meta-analysis summary statistics with linkage disequilibrium score regression (LDSC)^42^, yielding *h*^2^ = 0.056 (standard error (SE) = 0.0038). This value (and corresponding certainty of the measurement, i.e., the Z-score) exceeds the genome-wide SNP heritabilities and Z-scores reported for the constituent asthma GWAS used in our meta-analysis. Given that TAGC and GBMI aggregate samples from multiple countries, we used a global estimate of asthma prevalence in individuals of European ancestry (5.28%) rather than a country-specific estimate^43^. As discussed below, the modest genome-wide SNP-heritability of asthma means that risk prediction models constructed from common variants will have limited ability to distinguish cases from controls. We estimated the theoretical ceiling of such models to be 0.64 AUC (STAR★Methods), which is thus a critical value in which to contextualize the performance of the models we evaluate below. This ceiling reflects the proportion of asthma liability captured by common SNPs in the European- ancestry GWAS meta-analysis, rather than contributions from rare variants, family history, environmental exposures, or social determinants of health, which are not modeled in this study.

### Pediatric asthma case-control training and validation cohorts

While the European-ancestry meta-analysis provides the variant-level discovery foundation for gene-level prioritization, the biological interpretability and clinical relevance of these signals must ultimately be evaluated in the populations most affected by childhood-onset asthma^8,20,21^. The overwhelming majority of published polygenic scores train on European-ancestry individuals and subsequently demonstrate reduced performance in admixed or non-European populations^20^; however, these are the populations that bear the greatest burden of childhood-onset asthma^4,5^. To address this, we constructed case-control cohorts of childhood-onset asthma from two well-characterized cohorts: the Genetics of Asthma in Costa Rica Study (GACRS) and the Childhood Asthma Management Project (CAMP)^36,37^.

Cases were defined as unrelated pediatric individuals with physician-diagnosed asthma, whereas controls were unrelated individuals without an asthma diagnosis drawn from within each cohort. First-degree relatives were identified and excluded using pre-computed KING kinship coefficients^36,37^.

To characterize the ancestry composition of each cohort, we performed ADMIXTURE analysis^46^ at *K*=5 using genotyping data from unrelated GACRS and CAMP individuals run jointly alongside 1000 Genomes Project (Phase 3) individuals^47^ labeled with their assigned superpopulation: AFR, AMR, EAS, EUR, or SAS (**Figure 1B**, STAR★Methods). The ancestry composition of GACRS reflects a highly admixed population with predominantly European and admixed American ancestry proportions with a smaller African ancestry component, consistent with the known demographic history of the Costa Rican population, reflecting ancestral origins of Indigenous American, European and African individuals^48,49^. In contrast, CAMP consists of individuals with predominantly European or African American ancestry, reflecting the broader North American recruitment of this clinical trial cohort^37^.

Following cohort characterization with ADMIXTURE, we defined two cohorts for feature selection and model evaluation: one derived solely from GACRS samples (974 cases, 844 controls) and one derived solely from CAMP samples (597 cases, 64 controls). The severe case enrichment in CAMP reflects its original design as a clinical trial of pediatric asthma management rather than a population-based cohort study^37^, and reflects a meaningful constraint on our ability to use this cohort for modeling training and evaluation. Ultimately, we decided to perform all model training in the admixed GACRS cohort and performed evaluations in a held-out fold of GACRS as well as in the independent CAMP cohort. However, we accounted for the case-control imbalance in CAMP by constructing CAMP-Balanced, in which CAMP cases were bootstrap-downsampled to match the 64 CAMP controls across 100 iterations. This approach allowed us to calculate not only AUROC values across models, but also the uncertainty of these metrics via confidence intervals.

While we considered recruiting proxy controls from other studies, complications resulting from cohort-of-origin confounding would make it difficult to interpret the performance results. We hypothesized that another advantage of training in the GACRS cohort would be to minimize the selection of European-specific effects in our risk models, resulting in hopefully greater generalizability and portability across cohorts and diverse patient populations^20^.

### Multimodal risk score construction integrates variant-level and gene-level genetic signals

Asthma is a complex, polygenic disease driven by coordinated dysregulation across multiple tissues, immune cell types, and regulatory pathways such that no single level of biological resolution captures the full spectrum of inherited susceptibility^1,2,13^. In particular, variant-level approaches alone are insufficient for capturing the causal genes, cell types, and mechanisms driving pathogenesis^17^. Thus, resolving the gene-regulatory architecture of childhood-onset asthma susceptibility requires leveraging multiple levels of biological resolution: from variant-level effects identified by GWAS to the potential genes mediating these associations at the bulk tissue- level or cell-type-level resolved by single-cell gene expression data and modeled by TWAS. In this work, we evaluated the ability of variant-level and gene-level approaches to resolve effector genes, cell types, tissues, and mechanisms driving childhood-onset asthma susceptibility.

#### Variant-level polygenic risk score construction

We established variant-level baselines for comparison with our gene-level approaches by constructing PRS-CS models, which apply continuous shrinkage priors on SNP effect sizes to improve polygenic signal recovery^28^ and PRS-CSx models, which extend PRS-CS to a multi- ancestry framework by jointly modeling effect sizes across populations with coupled shrinkage priors^29^. For PRS-CS, we utilized the pre-computed European-ancestry LD reference panel constructed from 1000 Genomes Project (Phase 3) individuals. Then for PRS-CSx, in addition to the European-ancestry panel, we utilized pre-computed LD reference panels from the other four major superpopulations from the 1000 Genomes Project (Phase 3) individuals to ensure compatibility with the ancestrally diverse composition of GACRS and CAMP (STAR★Methods).

While the performance of these models was modest, as discussed below, we were motivated to pursue the multi-modal gene-level integration framework proposed next.

#### Transcriptome-wide association studies across bulk and cell-type-resolved tissues identify asthma-associated gene sets

Polygenic transcriptome risk scores (PTRS) leverage genetically predicted gene expression derived from transcriptome-wide association study (TWAS) frameworks to model transcriptome- mediated genetic risk, providing gene-level biological interpretation and potentially improving cross-ancestry portability relative to conventional PRS^32^. Aggregating predicted gene expression across multiple tissues consistently outperforms single-tissue models^32,38^, yet across published applications of PTRS in respiratory and allergic disease^50,51^, whole blood has served as the default reference tissue, and the optimal selection of disease-relevant tissues for PTRS construction in asthma has yet to be established. Bulk whole blood expression profiles are unlikely to capture the entirety of disease-relevant regulatory programs driving genetic risk, given the coordinated immune dysregulation across airway epithelial cells, mast cells, and adaptive immune populations that characterize asthma pathophysiology^2^. Consistent with this, prior work found that nasal airway transcriptomic models captured genetically driven asthma risk signals that were absent from whole blood^52^. We therefore hypothesized that expanding the PTRS feature space from bulk tissue-derived models to single-cell, cell-type-resolved expression models would improve the biological resolution of transcriptome-mediated genetic risk for asthma, with potential gains in cross-ancestry portability.

To test this, we leveraged *cis-*genetic gene expression prediction models created with the FUSION TWAS framework^31^ applied to two complementary gene expression resources: bulk tissue-derived models and cell-type-resolved models (**Figure 2A**). For the former, FUSION models were constructed from bulk RNA-seq data derived from 39 GTEx tissues each with matched genotyping data across N = 320 individuals^38,53^. For the latter, FUSION models were built from pseudobulked single-cell RNA-seq (scRNA-seq) data from 17 peripheral blood mononuclear cell (PBMC) subtypes identified from the OneK1K cohort with matched genotyping data ranging from N = 809 - 969 individuals^39,54^. We applied these gene models to our European-ancestry GWAS meta-analysis summary statistics to compute gene-tissue and gene-cell-type association statistics (**Figure 2B**; **Supplementary Figures S1-S2**; STAR★Methods). Gene–tissue and gene–cell-type pairs were retained for downstream analysis based on three quality filters applied uniformly across both pipelines: nonzero TWAS Z-score, positive *cis-*heritability, and TWAS cross-validation R^2^ > 0 (STAR★Methods)^54^. This filtered set served as input to two complementary gene-prioritization approaches.

**Figure 2.**
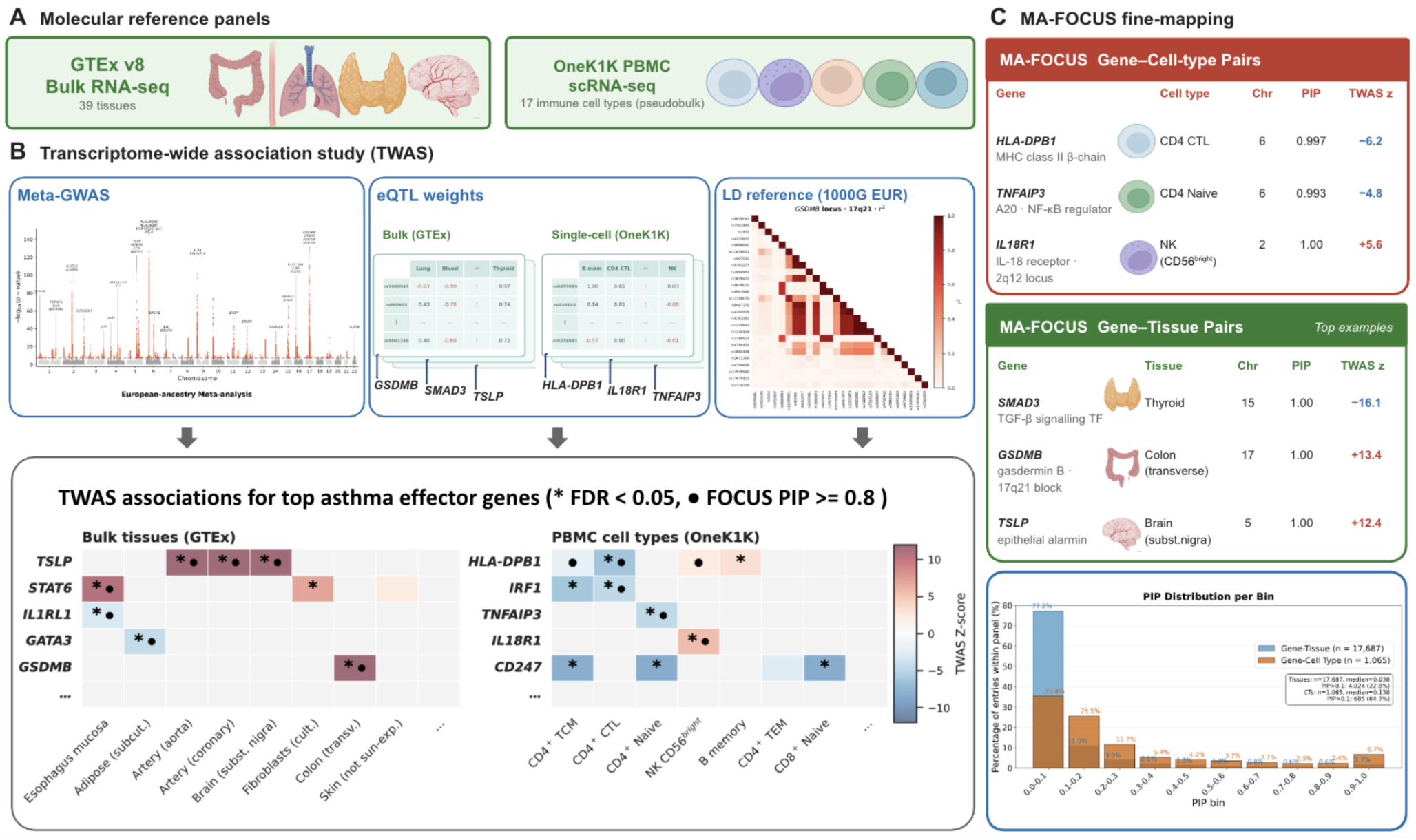
Overview of molecular data and gene-level prioritization scheme for asthma risk models. (**A**) Molecular reference panels comprising bulk RNA-seq data from 39 GTEx v8 tissues and pseudobulked single-cell RNA-seq data from 17 OneK1K peripheral blood mononuclear cell (PBMC) types. (**B**) Transcriptome-wide association studies (TWAS) integrated European-ancestry asthma meta-GWAS summary statistics, context-specific *cis-*eQTL prediction weights, and a European-ancestry linkage disequilibrium (LD) reference panel. Heatmaps show TWAS Z-scores for representative asthma-associated genes across bulk tissues and PBMC cell types; asterisks indicate FDR-significant associations, and circles indicate gene–context pairs with FOCUS posterior inclusion probability (PIP) ≥ 0.8. (**C**) Representative gene-context pairs prioritized by multi-ancestry FOCUS (MA-FOCUS), with chromosome, PIP, and TWAS Z-score shown. The histogram summarizes PIP distributions across fine-mapped gene–context pairs. Red and blue denote positive and negative TWAS Z-scores, respectively. Abbreviations: eQTL, expression quantitative trait locus; GWAS, genome-wide association study; PBMC, peripheral blood mononuclear cell; PIP, posterior inclusion probability; TWAS, transcriptome-wide association study.

First, a baseline TWAS pruning and thresholding (TWAS P+T) approach was applied, in which genes within each tissue or cell type were ranked by TWAS p-value and clumped at the gene level using a squared Pearson correlation threshold of R^2^ < 0.1. This correlation reflects the similarity between pairs of genes’ *cis-*genetically predicted expression vectors estimated in the reference panel, and is therefore distinct from SNP-level LD. Genes surviving clumping were then thresholded at four significance levels (θ ∈ {5 × 10^-5^, 5 × 10^-4^, 5 × 10^-3^, 5 × 10^-2^})^32^, yielding four nested candidate gene sets per genomic feature. Second, probabilistic fine-mapping was performed with FOCUS^34^, which uses nominal TWAS association (p < 0.05) as a recommended input criterion and assigns posterior inclusion probabilities (PIPs) to candidate causal transcripts within each locus (**Figure 2C**).

Across 39 GTEx tissues, FUSION TWAS identified 39,828 nominally significant (and 12,018 FDR 5% significant) gene-tissue pairs from 226,122 tested pairs. Across the 17 OneK1K PBMC pseudobulked cell types, FUSION TWAS identified 2,170 nominally significant (and 791 FDR 5% significant) gene- cell-type pairs from 19,055 tested pairs (**Figure 3A-B**, **Supplementary Figure S1-S2**; STAR★Methods). Despite larger sample size in the OneK1K cohort, we detected fewer associated genes, likely due to the increased noise and sparsity of single cell gene expression data. However, associated genes by GTEx and OneK1K are likely biologically complementary, as genes associated with specific immune cell types may represent regulatory programs that are obscured by heterogeneous bulk expression profiles^40^, motivating the inclusion of both modalities in downstream FOCUS fine-mapping and PTRS construction.

**Figure 3.**
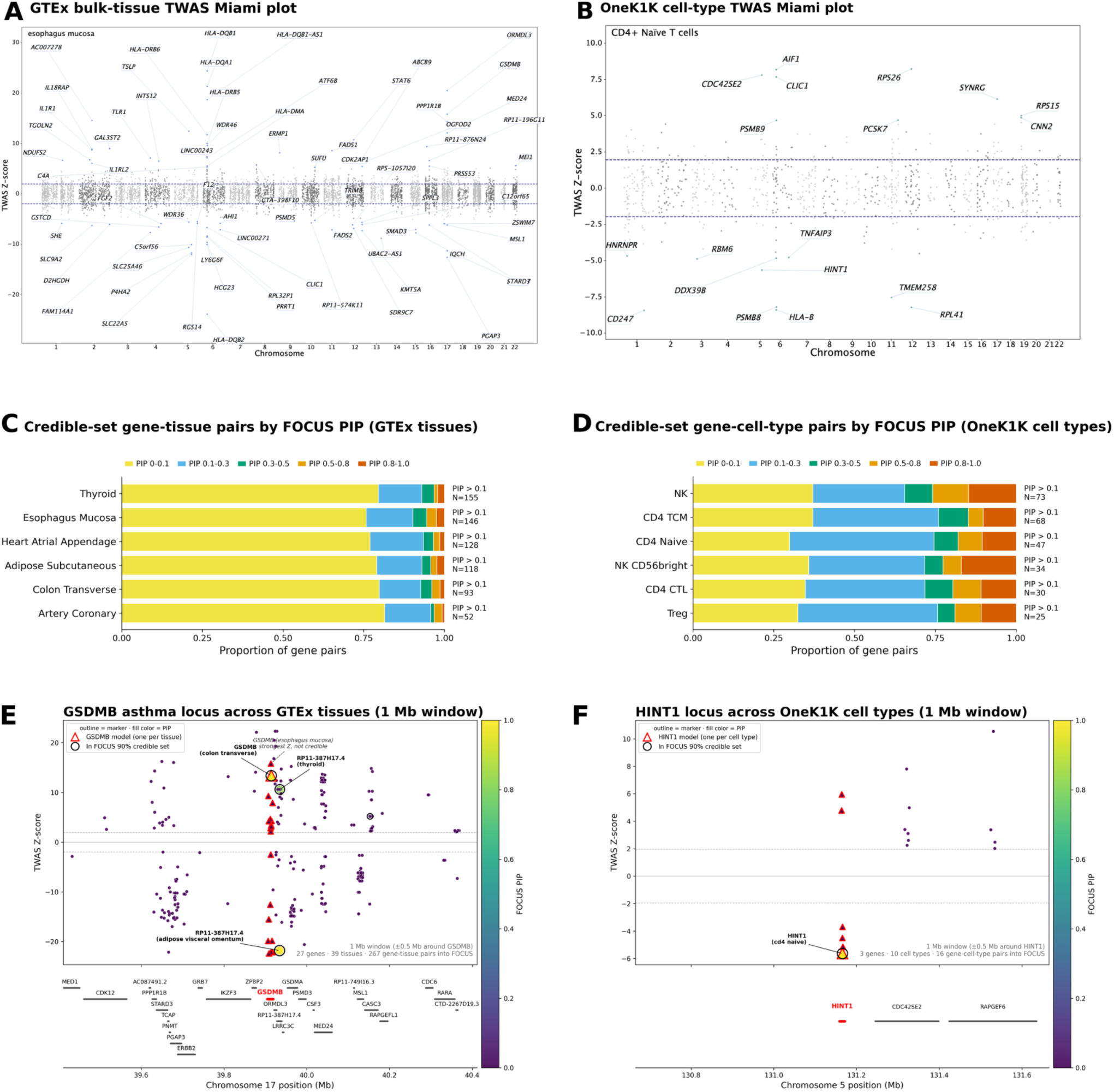
Gene-level genetic architecture of asthma at tissue and cell type resolution. (**A-B**) Miami plots displaying TWAS results with (**A**) GTEx bulk-tissue and (**B**) OneK1K pseudobulked PBMC cell type gene expression models (representative panels: **A**, esophagus mucosa; **B**, CD4+ naïve T cells), where each point represents a gene. Horizontal dashed lines mark the transcriptome-wide significance threshold. (**C-D**) Stacked bar plots displaying the proportion of gene-tissue pairs or gene-cell-type pairs in 90% credible sets according to their FOCUS PIP across 6 GTEx bulk tissues and 6 OneK1K pseudobulked PBMC cell types, respectively. (**E-F**) Locus-specific Miami plots centered on (**E**) the 17q21 asthma locus in GTEx tissues (focal gene *GSDMB*) and (**F**) the *HINT1* locus on chromosome 5 in OneK1K cell types. Each point represents one gene × tissue/cell-type model. Horizontal dashed lines mark the transcriptome-wide significance threshold.

To further evaluate whether the associated genes reflect asthma-relevant biology prior to fine- mapping with FOCUS, we identified unique genes among the FDR 5% significant pairs above and performed Gene Ontology (GO) term enrichment analysis with GOATOOLS^55–57^ against background genes also tested by TWAS to assess whether these gene sets were enriched for pathways relevant to asthma pathophysiology. Specifically, we assessed enrichment across 3,524 unique genes from the GTEx tissue TWAS analysis and 479 unique genes from the OneK1K single cell TWAS analysis **(Supplementary Figure S3**). Both modalities showed significant enrichment for peptide antigen assembly with major histocompatibility complex (MHC) class II protein complex (GO:0002503; tissue fold-enrichment = 5.70, FDR = 1.53 × 10^-6^; cell-type fold-enrichment = 5.53, FDR = 1.69 × 10^-3^), positive regulation of immune response (GO:0050778; tissue fold-enrichment = 3.39, FDR = 4.80 × 10^-4^; cell-type fold-enrichment = 5.12, FDR = 3.17 × 10^-3^), antigen processing and presentation of exogenous peptide antigen via MHC class II (GO:0019886; tissue fold-enrichment = 3.72, FDR = 9.66 × 10^-4^; cell-type fold-enrichment = 4.30, FDR = 7.30 × 10^-3^), and antigen processing and presentation (GO:0019882; tissue fold-enrichment = 3.19, FDR = 9.66 × 10^-4^; cell- type fold-enrichment = 3.75, FDR = 7.30 × 10^-3^) (**Supplementary Figure S4**). In addition to biological processes, our gene sets were enriched for cellular component terms such as MHC class II protein complex (GO:0042613; tissue fold-enrichment = 5.30, FDR = 3.02 × 10^-6^; cell-type fold-enrichment = 5.11, FDR = 6.16 × 10^-4^) and molecular function terms such as peptide antigen binding activity (GO:0042605; tissue fold-enrichment = 4.11, FDR = 1.43 × 10^-5^; cell-type fold-enrichment = 5.59, FDR = 7.99 × 10^-8^), implicating MHC class II-mediated antigen presentation as the primary biological signal in both modalities. Tissue-specific enrichment was also observed for interleukin-1 receptor activity (GO:0004908; fold-enrichment = 5.70, FDR = 2.30 × 10^-2^), consistent with the genetic association of *IL33*/*IL1RL1* with asthma susceptibility^15,58^, and the established mechanistic role of these factors in driving airway inflammation^59^. Chromatin remodeling (GO:0006338; fold-enrichment = 1.63, FDR = 2.12 × 10^-2^) also displayed tissue-specific enrichment, likely reflecting broader transcriptional regulatory activity in the tissue-level gene set rather than asthma-specific biology. Fold-enrichment values reflect the respective TWAS-tested gene backgrounds for each modality (24,866 unique genes for tissues; 3,571 unique genes for cell types) and are not directly comparable across analyses.

The convergent enrichment of MHC class II antigen presentation and positive regulation of immune response across both modalities is consistent with the established role of MHC class II- restricted CD4+ T helper cell activation in allergic airway inflammation. Asthma GWAS have long identified HLA loci among the most significantly associated genomic regions^1,15^, and mechanistic studies have demonstrated that MHC class II-mediated antigen presentation to CD4+ T helper 2 (Th2) cells drives the IgE-mediated and eosinophilic responses central to asthma pathophysiology^60^. Accordingly, endotype frameworks characterize Th2-high asthma as driven by Th2 cell-mediated inflammation, the initiation of which is contingent on MHC class II-mediated antigen presentation^2^. Together, these findings support the biological relevance of the TWAS- identified gene sets for downstream fine-mapping with FOCUS.

#### Probabilistic fine-mapping improves PTRS

Having established the biological relevance of TWAS-identified gene sets, we next sought to refine the feature space for transcriptomic risk score construction through statistical gene-level fine-mapping. TWAS signals at a given locus frequently span multiple co-regulated transcripts, reflecting shared *cis-*genetic regulation among nearby genes and among co-regulated tissues rather than independent causal effects. Consequently, retaining all TWAS-significant gene models as PTRS features risks introducing redundancy that dilutes true causal associations^33,34^. Pruning and thresholding (P+T) approaches exacerbate this problem by discarding potentially informative signals and do not explicitly model correlation structure among genes, whereas fine-mapping methods probabilistically assign PIPs to independent causal signals to retain a more accurate and non-redundant feature set. This distinction has practical consequences for model performance: fine-mapping of GWAS associations has been shown to improve PRS accuracy and cross-cohort transferability by prioritizing putative causal effects over tagging effects^61^. We therefore hypothesized that an analogous improvement could be achieved at the transcriptomic level. We applied FOCUS probabilistic fine-mapping, which jointly models co-regulation between all gene models within a locus and computes PIPs over candidate causal transcripts, incorporating ancestry-specific LD patterns and transcriptional regulation to disentangle co-regulated transcripts^35^.

We applied FOCUS at the locus level to every TWAS-significant (nominal TWAS p-value < 0.05) gene-context pair separately for the 39-tissue GTEx panel and the 17-cell-type OneK1K panel. This threshold is deliberately more lenient than the FDR < 5% cut-off used for the **Figure 3A-B** GO enrichment, as FOCUS leverages nearby co-regulated gene-tissue and gene-cell type pairs in order to perform inference. For each focal gene, all gene models with transcription start sites located within ±1 Mb of its transcription start site (TSS) were included in the fine-mapping locus, irrespective of their individual TWAS p-values. FOCUS then jointly accounted for co-regulation among these models and estimated their posterior inclusion probabilities (PIPs) (STAR★Methods). Gene models satisfying FOCUS posterior support were retained as causal features for PTRS construction. We applied asymmetric inclusion criteria to the two panels: in the bulk-tissue (GTEx) panel, we retained gene-tissue pairs with PIP ≥ 0.1 and residing in a 90% FOCUS credible set, whereas in the cell-type-resolved (OneK1K) panel, we retained all gene-cell-type pairs within a 90% FOCUS credible set without a PIP threshold (STAR★Methods). This panel-specific strategy reflects differences in the number of TWAS associated genes and the degree of co-regulation across tissues and cell types. For example, GTEx produced a large, highly co-regulated set of TWAS-significant gene-tissue pairs, where a single putative causal gene often generated nonzero PIPs across multiple tissues. Consequently, GTEx 90% credible sets contained many genes with low PIP (median PIP ≈ 0.04; 77% with PIP < 0.1; STAR★Methods). Applying a threshold of PIP ≥ 0.1 reduced the number of co-regulated genes while retaining 4,033 high-confidence gene-tissue features (**Supplementary Figure 5**). By contrast, OneK1K yielded fewer gene-cell-type pairs at these same criteria (*n* = 688), consistent with highly variable numbers of cells per donor and lower signal to noise ratio^54^. However, OneK1K credible sets were smaller and more concentrated than GTEx credible sets (median PIP ≈ 0.14; only 35% with PIP < 0.1; STAR★Methods) and thus did not require a PIP threshold to address the large credible set issue. Therefore, we elected to retain the full OneK1K 90% credible set and remove the PIP threshold resulting in 1,065 high-confidence gene-cell-type pairs (**Supplementary Figure 5**). As a result, for each modality, we were able to preserve a few thousand fine-mapped genes as features for our PTRS models discussed below. We note that we did not perform FOCUS fine-mapping jointly on gene models derived from GTEx and OneK1K due to different statistical power, gene model performance, and sample size. In **Figure 3C-D**, we show the distribution of PIPs across the six tissues and six cell types with the most gene-context pairs with PIP ≥ 0.1. In **Supplementary Table S2**, we report all fine-mapping results including PIPs and credible set assignments.

Furthermore, probabilistic fine-mapping implicated a biologically coherent set of genes across tissues and cell types, although these should be interpreted as potentially relevant regulatory contexts (or at least best proxy tissues) rather than definitive primary mediators of asthma risk. Across bulk GTEx tissues, FOCUS resolved canonical Th2-associated genes at the top of multiple credible sets, including *TSLP* in coronary artery, *IL1RL1* and *STAT6* in esophageal mucosa, *IL18R1* in heart atrial appendage, and *GATA3* in subcutaneous adipose tissue^2,15,60^. As discussed above, Th2 cells are the principal driver of eosinophilic airway inflammation, defining the two major endotypes of severe asthma as Th2-high (eosinophilic) and Th2-low (non-eosinophilic)^2^. This recovery of canonical Th2 genes supports the utility of fine-mapping for prioritizing established asthma susceptibility genes. Multiple effector genes were related to alarmin cytokine signaling: *TSLP* (PIP = 1.00, coronary artery, Z = 8.5); the *IL-33/IL-18* receptor cluster on chromosome 2 represented by *IL1RL1*^15^ (PIP = 1.00, esophagus mucosa, Z = -5.1) and *IL18R1* (PIP = 1.00, heart atrial appendage, Z = 15.0); the master Th2 transcription factor by *STAT6* (PIP = 0.998, esophagus mucosa, Z = 10.3); and the Th2 lineage-defining transcription factor *GATA3* (PIP = 0.974, adipose subcutaneous, Z = -4.2).

The canonical *17q12–21* childhood-asthma locus^15,16^ was resolved to *GSDMB* (PIP = 1.00, colon transverse, Z = 12.7; **Figure 3E**). This locus illustrates the resolution gained through fine-mapping over marginal TWAS association: within the 1 Mb window, 267 gene models representing 27 genes across 39 tissues were evaluated. We note that not all pairs of genes and tissues can be tested, as only 267 produced a *cis-*genetic predictive model with FUSION. Several nearby genes, including *ORMDL3*, *GSDMA*, and *PGAP3*, showed strong TWAS association strength, yet FOCUS assigned high PIPs exclusively to *GSDMB* and two less well-characterized pseudogenes. Notably, *GSDMB* was most strongly associated with asthma via the esophagus mucosa (Z = 14.8; **Figure 3A**), but this tissue model received negligible posterior inclusion probability (PIP = 0), underscoring that marginal TWAS significance is not necessarily a proxy for identifying putative causal genes. In other loci, FOCUS prioritized *SMAD3* (PIP = 1.00, thyroid, Z = −15.4), a central mediator of *TGF-β* signaling. *TGF-β* contributes to asthma-associated airway remodeling by promoting fibroblast activation, extracellular-matrix deposition, and structural changes in the airway wall^62^. FOCUS also prioritized *BACH2* (PIP = 1.00, pancreas, Z = 13.4), a transcriptional regulator that restrains *Th2* effector differentiation and supports regulatory T cell homeostasis^63^, *RUNX3 (PIP = 1.00, testis, Z = −6.1),* a transcription factor required for CD8+ cytotoxic T cell lineage development^64^. Additional signals included the MHC class II genes *HLA-DQA1* (PIP = 1.00, spleen, Z = 2.7) and *HLA-DQB1* (PIP = 1.00, brain substantia nigra, Z = −10.2), as well as the antigen-processing gene *TAP2* (PIP = 1.00, stomach, Z = −8.9). *HLA-DQB1* encodes a component of the *HLA-DQ* complex that presents extracellular, including allergen-derived, peptides to CD4+ T cells, and its genomic region is an established asthma susceptibility locus^15^.

The distribution of these signals across barrier epithelia (esophagus mucosa, skin, stomach, and transverse colon), vascular tissue (coronary artery), and metabolic, lymphoid, endocrine, and neural contexts (subcutaneous adipose, spleen, pancreas and thyroid, and brain substantia nigra, respectively) may partly reflect limitations of steady-state expression reference panels. If the true mediating tissue is absent from our panel, FOCUS may assign high PIPs to the best proxy tissues. Although asthma has systemic effects and has been associated with neuroinflammatory changes and an increased risk of Parkinson’s disease, current evidence does not establish the substantia nigra as a direct mediator of asthma susceptibility or explain the *HLA-DQB1* association in this tissue^65,66^. The brain substantia nigra assignment may therefore reflect cross-tissue co-regulation, resident immune-cell expression, or the absence of a better-matched airway or immune context.

On the other hand, our results may indeed reveal a more systemic dysregulation of gene expression across different tissues of the human body in asthma pathogenesis.

Among cell-type-resolved OneK1K models, FOCUS identified distinct asthma signals within specific immune lineages. *IL18R1* was fine-mapped to natural killer (NK) CD56^bright^ cells (PIP = 1.00) localizing this IL-18 receptor signal to an innate lymphocyte subset that constitutively expresses receptors for IL-12 and IL-18 and is specialized for cytokine production^67,68^. This cell-type resolution was not available from bulk tissue, where *IL18R1* was instead prioritized in heart atrial appendage. *IRF1*, a transcription factor gene in the IL-33/Th2 pathway, was fine-mapped to CD4+ cytotoxic T cells (PIP = 0.997, Z = -6.6). *TNFAIP3*, which encodes A20, was localized to naïve CD4+ T cells (PIP = 0.993, Z = −4.7). A20 restrains TCR-induced NF-κB activation and supports T-cell homeostasis, providing a plausible basis for this cell-type assignment^69^. MHC class II prioritizations were resolved by T cell subset: *HLA-DPB1* in CD4+ central memory T cells (PIP = 1.00) and *HLA-DPA1* in CD4+ cytotoxic T cells (PIP = 0.997), while *TAPBP* was resolved to CD4+ regulatory T cells (PIP = 1.00), implicating antigen-presentation machinery within known T cell lineages. Additional well-known asthma genes appeared in biologically expected cell types but at lower PIP. These included *RORA* in CD4+ central memory T cells (PIP = 0.61), *RUNX3* in NK cells (PIP = 0.70), *IL4R* in naïve B cells (PIP = 0.14), and *ETS1* in naïve CD4+ T cells (PIP = 0.26). *RORA* is an established asthma locus and regulates type 2 immune-cell differentiation^15^, whereas *IL4R* mediates IL-4 and IL-13 signaling and contains variants associated with asthma susceptibility and severity^70^. *RUNX3* regulates cytotoxic lymphocyte differentiation and Th1–Th2 balance^64^, while *ETS1* contributes to naïve T-cell development and immune homeostasis. These genes were included in their locus-specific 90% credible sets but were not the models with the highest PIPs, indicating that posterior support was shared across multiple co-regulated candidates, although this was not generally representative of applying FOCUS to OneK1K gene models, whose credible sets were on average more concentrated than those from GTEx, as discussed above (**Figure 3E–F**).

Beyond recovering canonical effector genes, cell-type-resolved fine-mapping also highlighted additional candidate signals. At a chromosome 5 locus, FOCUS evaluated 16 gene models representing three genes across 10 cell types and prioritized *HINT1* in naïve CD4+ T cells (PIP = 0.99, Z = −5.5; **Figure 3F**) over its co-regulated neighbors *CDC42SE2* and *RAPGEF6* (PIP < 0.006). *HINT1* is involved in allergy-relevant mast-cell biology: in FcεRI-activated mast cells, the second messenger Ap4A disrupts the *HINT1–MITF* interaction, releasing *MITF*-dependent transcriptional programs^71^. This signal is local and cell-type-specific, distinct from the canonical 5q31 Th2 cytokine cluster (*IL4/IL5/IL13/RAD50*), which lies outside the fine-mapped window. Establishing its independence from those variants would require further conditional analysis. Taken together, performing fine-mapping on cell-type-resolved models does not just recapitulate bulk tissue-level findings, but assigns the relevant asthma-effector genes to specific immune lineages to provide greater mechanistic resolution that tissue-level TWAS alone cannot provide.

#### Nonlinear modeling of gene-level features improves biological resolution of PTRS

While conventional PTRS predicts disease status using linear combinations of genetically predicted gene expression values scaled by gene-level effect sizes^32^, this approach assumes that each gene contributes independently and additively to disease risk. However, this assumption may not hold in complex diseases such as asthma, where coordinated interactions between cell types and regulatory pathways may produce nonlinear effects on disease risk that additive models cannot capture^1,2^. To test this hypothesis, we compare linear variant-level PRS models, linear gene-level PTRS models, and a panel of nonlinear classifiers (grid-searched random forest, gradient boosting, and other tree- and ensemble-based methods; full set in STAR★Methods) constructed from FOCUS fine-mapped TWAS associations of a single modality (either tissue-level or cell-type-level gene effects) (**Figure 4A**). For these analyses, we conducted model training in the GACRS train set (60%), performed hyperparameter tuning in the GACRS validation set (20%) and used this set to filter out models for which asthma controls received higher risk scores on average than asthma cases, evaluated within-cohort performance on the held-out GACRS test set (20%), and evaluated cross-cohort performance on the independent CAMP Balanced set. While CAMP-Balanced is entirely independent of the training cohort, we elect to report AUROC (Area Under the Receiver Operating Characteristic Curve) results predominantly for this cohort, or AUC for brevity, rather than also for GACRS-test. AUC values should be interpreted relative to an approximate theoretical ceiling of 0.64 for a common variant genetic predictor of asthma, calculated from our SNP heritability (0.056) and assumed population prevalence (5.28%) under the liability-threshold framework^72^. In the next section, we evaluate cross-modal PTRS involving the trio of variant-level effects combined with fine-mapped tissue- and cell-type-level gene effects.

**Figure 4.**
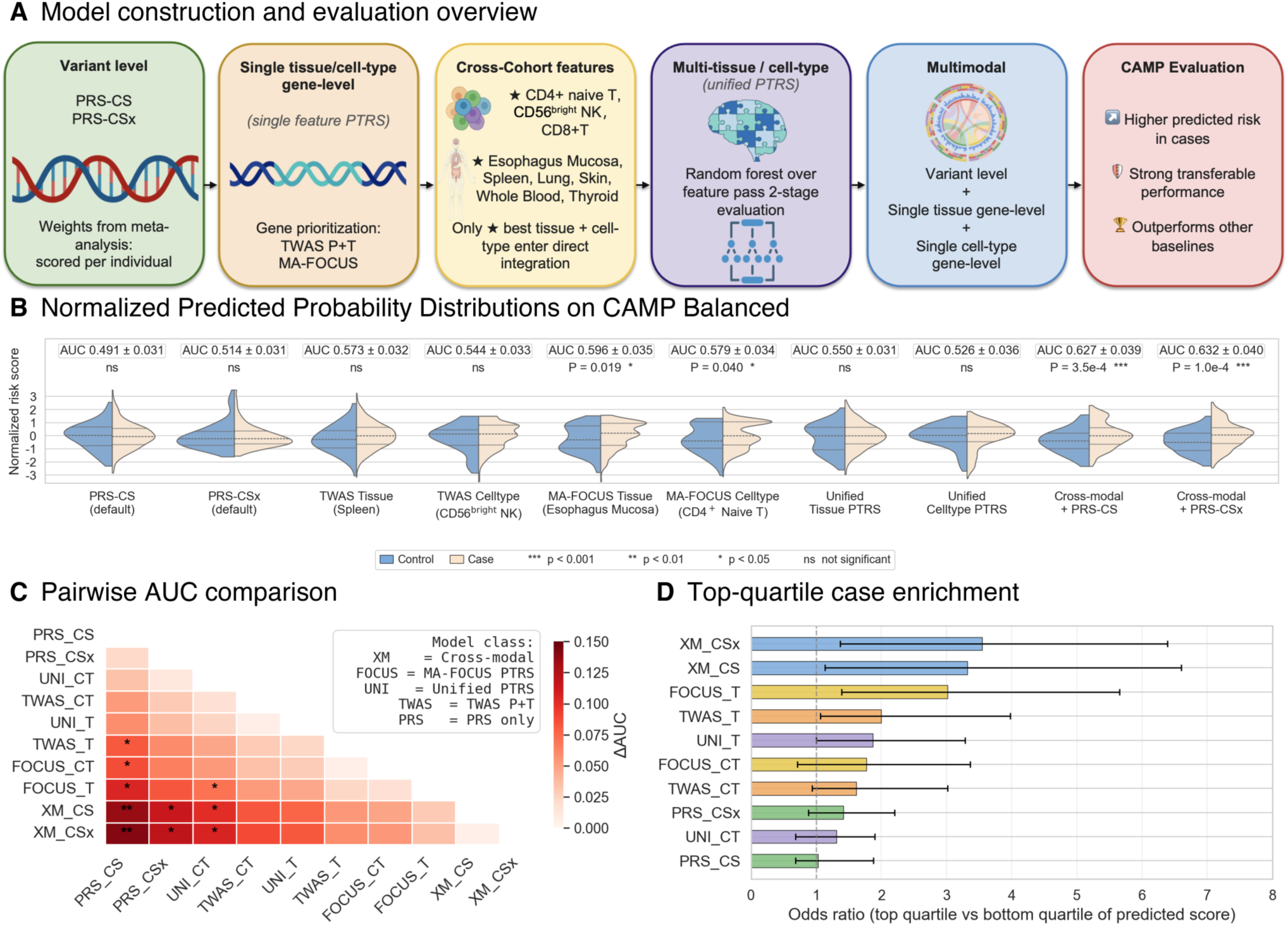
Cross-modal integration of gene-level PTRS and variant-level PRS for pediatric asthma risk stratification on the CAMP-Balanced cohort. (**A**) Model construction and evaluation overview. All models are trained on GACRS and evaluated on a CAMP-only sample (n = 597 cases, n = 64 controls). (**B**) Normalized predicted probability distributions on CAMP. Predicted probabilities are z-normalized per model before plotting so violin shapes and case-versus-control separation are visually comparable across classifiers with different raw score ranges. Split-violin plots of predicted asthma probability for cases (right half, beige) and controls (left half, blue) under the best of the ten evaluated models. Inner horizontal lines mark the 25th, 50th, and 75th percentiles of predicted values within each class. Above each violin: AUC = mean ± SD across 100 case-matched 64-vs-64 bootstrap iterations of the same CAMP cohort (the "CAMP-Balanced" evaluation); Welch’s two-sided t-test p-value = case-vs-control comparison on the full-cohort distributions shown (the t-test on means is robust to class imbalance and benefits from the full sample). Significance markers: *** p < 0.001, ** p < 0.01, * p < 0.05, ns = not significant. (**C**) Pairwise AUC comparison. Lower-triangular heatmap of ΔAUC = AUC(y-axis model) − AUC(x-axis model) computed across the paired CAMP-Balanced bootstrap iterations defined in (**A**); rows and columns are sorted in ascending order of mean CAMP-Balanced AUC. Significance is the empirical paired-bootstrap one-sided p-value (fraction of iterations with ΔAUC ≤ 0): * p < 0.05 (ΔAUC > 0 in > 95 % of iterations); ** p < 0.01 (> 99 %). Inset key: XM = Cross-modal, FOCUS = MA-FOCUS PTRS, UNI = Unified PTRS, TWAS = TWAS P+T, PRS = variant-level PRS only. (**D**) Top-quartile case enrichment. Horizontal bar chart of the asthma-case odds ratio (OR; top vs. bottom quartile of each model’s predicted-probability score on CAMP-Balanced), ranked by mean OR. Bars are the mean OR and error bars the 95 % bootstrap CI (2.5th–97.5th percentile) across the bootstrap iterations defined in (A); the dashed grey line marks OR = 1 (no enrichment). Bar color encodes model class: XM_CS or XM_CSx: Cross-modal (red); FOCUS_CT or FOCUS_T: MA-FOCUS PTRS (green); UNI_CT or UNI_T: Unified PTRS (brown); TWAS_CT or TWAS_T: TWAS P+T (grey); PRS_CS or PRS_CSx: PRS only (blue). Best-performing tissue- and cell-type-specific PTRS models are shown; comprehensive results across all 39 GTEx tissues and 17 OneK1K cell types are in **Supplementary Tables S4-S5**.

To establish the variant-level baseline, we evaluated PRS-CS and PRS-CSx (PRS-CS at ϕ = auto with the 1000 Genomes EUR LD reference; PRS-CSx at ϕ = auto with the META cross-population LD reference panel)^28,29^, fit as a logistic regression with the five 1000 Genomes super-population admixture proportions (AFR, AMR, EAS, EUR, SAS) as covariates (STAR★Methods). These baselines achieved only modest performance on the within-cohort held-out GACRS test set (PRS-CS AUC = 0.591; PRS-CSx AUC = 0.578) and weaker cross-cohort transfer to the independent CAMP- Balanced 64-vs-64 × 100-bootstrap evaluation (PRS-CS AUC = 0.491 ± 0.031; PRS-CSx AUC = 0.514 ± 0.031) (**Figure 4B**, left). For these models, there was no significant difference in the means of the score distributions between cases and controls (Welch t-test p > 0.05).

To establish the gene-level baseline, we evaluated TWAS PTRS using additive linear classifiers (ridge logistic regression at three regularization strengths, lasso logistic regression, and elastic- net logistic regression), consistent with the original PTRS framework^32^. We constructed TWAS PTRS separately for each of the 39 GTEx tissues and 17 OneK1K cell types and performed pruning and thresholding on their gene-level associations at four candidate p-value thresholds (TWAS p < 5×10⁻⁵, 5×10⁻⁴, 5×10⁻³, 5×10⁻²; LD-clumped at r² < 0.1 on predicted-expression correlation; full details in STAR★Methods). We refer to these gene-level scores as single-feature TWAS pruning and thresholding (P+T) PTRS. We observed that tissues or cell types that performed best in GACRS demonstrated poor transferability to the CAMP Balanced cohort. In an effort to utilize tissues and cell types that were meaningful across cohorts, we determined a subset of tissues and cell types that passed a “cross-cohort consistency filter” in which we require a weakly predictive AUC for both the GACRS test set and the CAMP Balanced cohort (STAR★Methods), enabling us to report the tissues or cell types that demonstrated cross-cohort transferability rather than those potentially overfitting to the GACRS cohort.

After applying the cross-cohort consistency filter, 6 of 39 GTEx tissues and 3 of 17 OneK1K cell types survived across at least one of the four TWAS p-value thresholds tested (**Supplementary Table S3**). Among these, the strongest tissue-level PTRS was derived from Spleen at TWAS p < 0.00005 using ridge logistic regression (CAMP-Balanced AUC = 0.5730 ± 0.032; **Figure 4B**). The strongest cell-type-level PTRS was derived from CD56^bright^ NK cells at TWAS p < 0.005, also using ridge logistic regression (AUC = 0.544 ± 0.034; **Figure 4B**, middle-left). However, for these models, we again note that there was no significant difference in the means of the score distributions between cases and controls (Welch t-test p > 0.05). Performance of models for other tissues and cell types are reported in **Supplementary Tables S4-S5**, which are correspond to their optimal significance threshold across the candidate thresholds tested (STAR★Methods). The importance of both spleen and NK cells to asthma pathophysiology are supported by previous studies.

Previous work implicated the spleen in asthma TWAS^52,73^. There is more compelling support for CD56^bright^ NK cells, which are an immunoregulatory, cytokine-producing subset and have been implicated in allergic airway inflammation^74^.

With baseline models established, we next sought to construct a PTRS consisting of fine-mapped TWAS associations at tissue- and cell-type-levels, separately. For both modalities, we employed the MA-FOCUS fine-mapped feature sets described above, trained various nonlinear models including random forest, and selected the top tissue or cell type based on performance on the held out GACRS test set and independent CAMP Balanced cohort. These FOCUS PTRS models respectively outperformed their corresponding linear TWAS PTRS model, described in the previous paragraph. Among the 39 GTEx tissues and across all classifiers evaluated (STAR★Methods), the strongest FOCUS PTRS was derived from esophageal mucosa using gradient boosting (CAMP-Balanced AUC = 0.596 ± 0.035; case–control Welch’s t-test p = 0.019; **Figure 4B**, middle). This prioritization does not necessarily indicate a direct contribution of the esophagus to asthma pathogenesis. Instead, it may reflect regulatory programs shared across type 2 inflammatory mucosal diseases. Eosinophilic esophagitis (EoE) is a chronic allergic disease of the esophageal mucosa that frequently co-occurs with asthma and similarly involves epithelial barrier dysfunction, eosinophilic inflammation, and IL-4/IL-13 signaling^75,76^. Because *cis-*regulatory effects are partly shared across tissues^38^, the well-powered esophageal mucosa panel may serve as a proxy for regulatory programs active in airway epithelium, whose involvement in asthma is well established^77^. This interpretation remains a mechanistic hypothesis rather than evidence that esophageal tissue directly mediates asthma risk. Among the 17 OneK1K cell types, the strongest FOCUS PTRS was derived from naïve CD4+ T cells using gradient boosting (AUC = 0.579 ± 0.034; Welch’s t-test P = 0.040; **Figure 4B**, middle). This finding is consistent with the role of naïve CD4+ T cells as precursors of allergen-responsive Th2 cells, which produce IL-4, IL-5, and IL-13 and coordinate eosinophilic airway inflammation^60^. Both models exceeded their corresponding linear TWAS P+T baselines, spleen for tissues and CD56^bright^ NK cells for cell types, in terms of absolute AUC and in terms of producing scores that were significantly larger in cases than controls. Rather than comparing only the top performing models, if we consider the tissues and cell types that passed the cross-cohort consistency filter, we found that TWAS P+T PTRS and FOCUS PTRS prioritize different features. For example, only salivary gland was shared at the tissue-level and CD4+ cytotoxic T cells were shared at the cell-type-level. Minor salivary glands are epithelial secretory organs that contribute to oral mucosal defense through secretory IgA, antimicrobial factors, and resident immune-cell populations^78,79^. These epithelial, secretory, and immune- regulatory properties provide a plausible basis for their prioritization in both approaches. The CD4+ cytotoxic T-cell overlap has more direct disease relevance: cytotoxic CD4+ tissue-resident memory cells are enriched in the airways of patients with severe asthma and express pro- inflammatory and cytotoxic programs implicated in persistent inflammation and airway remodeling^80^. Although these airway-resident cells are not equivalent to the peripheral-blood CD4+ cytotoxic population modeled by OneK1K, they support the broader relevance of this lineage to asthma. While the strength of TWAS associations is driven both by causal signals and co- regulated tagging genes and tissues, it is reasonable that these two approaches result in different prioritized genes and corresponding tissues or cell types.

Next, we trained a set of nonlinear models leveraging fine-mapped genes from multiple tissues or multiple cell types, separately. We refer to these as tissue-level or cell-type-level unified PTRS models. To this end, we leveraged the top 8 GTEx tissues (esophageal muscularis, esophageal mucosa, sun-protected skin, subcutaneous adipose, pituitary, minor salivary gland, aorta, and adrenal gland) and 3 OneK1K cell types (CD8+ central memory T cells, CD4+ cytotoxic T lymphocytes, and naïve CD4+ T cells) that were each predictive as single-feature models in both the within-cohort GACRS test set and the out-of-cohort CAMP Balanced cohort and thus passed the cross-cohort consistency filter (**Figure 4B**, middle-right). The tissue-level unified PTRS achieved an AUC of 0.550 ± 0.031, although it yielded no significant mean difference between the cases and controls (Welch t-test P > 0.05). The cell-type-level unified PTRS achieved an AUC of 0.526 ± 0.036, similarly with no significant difference between the average predictions of cases vs. controls. So, although we hypothesized that modeling multiple predictive tissues or cell types simultaneously would improve discrimination between cases and controls, these models did not outperform the more parsimonious single-feature FOCUS PTRS models discussed in the previous paragraph.

We then considered the predictive value of jointly modeling variant-level baselines from PRS- CS(x) within these unified models. Adding the variant-level PRS-CSx baseline to the tissue-level and cell-type-level FOCUS unified PTRS modestly improved performance in CAMP-Balanced. The combined PRS-CSx and tissue-level PTRS features was provided to a gradient boosting classifier, achieving an AUC of 0.563 ± 0.0315, corresponding to an increase of 0.013 AUC over the tissue- level unified PTRS and 0.049 AUC over PRS-CSx alone. By contrast, applying gradient boosting to the combined PRS-CSx and cell-type-level unified PTRS features produced a larger relative improvement over the CT unified PTRS (AUC = 0.554 ± 0.033; ΔAUC = 0.028 over unified PTRS, 0.040 over PRS-CSx alone). Analogous integrations with PRS-CS yielded uniformly weaker performance in this cohort, with no classifier beating the corresponding unified PTRS baseline (tissue-level best: random forest AUC = 0.530 ± 0.034; cell-type-level best: random forest AUC = 0.503 ± 0.038), indicating that PRS-CS does not compose additively with the unimodal transcriptomic signal in this cohort. These findings suggest that, for both modalities, the unified PTRS captured most of the discriminative signal achieved by within-modality integration with variant-level PRS-CSx providing only modest gains over the corresponding unified PTRS alone (ΔAUC = 0.013 for tissue and 0.028 for cell type). Nevertheless, the combined PRS–PTRS model performed better for the tissue modality, consistent with its stronger unified PTRS baseline. These results motivated our subsequent evaluation of a cross-modal architecture integrating selected tissue-level and cell-type-level PTRS features with variant-level PRS.

### Cross-modal integration of PRS and FOCUS-derived PTRS improves risk stratification and biological interpretability

While the unified within-modality PTRS architectures, each based on the three OneK1K cell types or eight GTEx tissues that passed the cross-cohort consistency filter, demonstrated rather poor transferability to our CAMP Balanced cohort, we set out to leverage both modalities simultaneously. Rather than expanding the within modality feature set, which underperformed the strongest single feature from each modality, we tested a deliberately sparse two-feature cross- modal design. Feature-count ablation analyses using this pair as the anchor showed that adding a third consistent-shortlist feature reduced CAMP-Balanced AUC in all experiments (mean ΔAUC = −0.031; **Supplementary Table S6**). Moreover, because these models demonstrated modest performance overall, we shifted our focus from developing a highly discriminative score to an evaluation of the prioritized tissues and cell types and the value of modeling their gene-level features using nonlinear approaches. To this end, we constructed a cross-modal nonlinear model that combines features of the top PTRS models with PRS-CS(x) and trains a hyperparameter- tuned random forest in the GACRS training cohort. These top PTRS models were the cell-type- level FOCUS PTRS (naïve CD4+ T cells) and the tissue-level FOCUS PTRS (esophageal mucosa). Full architectural and PRS configuration details are included in the STAR★Methods. The cross-modal architecture yielded the highest cross-cohort predictions observed in our study (AUC = 0.627 ± 0.039, OR = 3.32 for model with PRS-CS, AUC = 0.632 ± 0.040, OR=3.55 for model with PRS-CSx; **Figure 4B**, right). The latter was the top-performing model across the entire architecture search and the sole configuration in which the CAMP-Balanced AUC remained competitive with the GACRS within-cohort test AUC (0.598), which suggests that these features are transferable between cohorts and thus biologically meaningful. In **Supplementary Table S7**, we report the performance of all tested models. However, we note that these more sophisticated models only nominally outperform the tissue-level FOCUS PTRS for esophageal mucosa (AUC = 0.596 ± 0.035; ΔAUC = +0.036, empirical one-sided p-value = 0.13). Overall, our results suggest that jointly modeling variant-level and gene-level risk factors are important, as not all genetic risk is mediated by gene expression levels (especially those measured at steady state in limited cellular contexts).

To complement our AUC-based evaluation with an interpretable measure of risk stratification, we quantified asthma enrichment as the odds ratio (OR) comparing individuals in the top versus bottom quartile of each model’s predicted probability score (**Figure 4D**, STAR★Methods). For example, the 95% CI of PRS-CS includes an OR of 1, indicating that higher variant-level PRS are not significantly enriched for asthma cases and therefore does not meaningfully stratify asthma risk. In contrast, cross-modal integration of tissue-level and cell-type-level PTRS with variant-level PRS recovers strong top-quartile enrichment (with PRS-CSx OR = 3.55), notably exceeding the top tissue-level FOCUS PTRS (OR = 3.02), the top cell-type-level FOCUS PTRS (OR = 1.77), and both within-modality unified PTRS models evaluated alone, without PRS integration (tissue-level OR = 1.87; cell-type-level OR = 1.31). Adding variant-level PRS-CSx to either uni-modal unified PTRS yielded only marginal improvements in stratification (tissue level with PRS-CSx OR = 1.70; cell-type level with PRS-CSx OR = 1.66), confirming that cross-modal integration of tissue and cell-type level features is what drives the substantial top-quartile enrichment.

Because CAMP-Balanced included only 128 individuals per bootstrap iteration, we compared the top and bottom quartiles rather than deciles or 5% tails, which would produce sparse contingency-table cells and potentially unstable or biased odds-ratio estimates^81^. To contextualize our cross-modal OR of 3.55 with other studies, we compared to a multi-ancestral pediatric asthma PRS study, in which top-versus-bottom 5% ORs ranged from 2.80 to 5.82 across three cohorts^82^, but was not as enriched as the top-versus-bottom-decile (OR = 8.3, 95% CI: 7.4–9.3) reported for childhood-onset asthma in UK Biobank^25^. We caution that these estimates may not be directly comparable because of differences in cohort size, ancestry composition, phenotype definition, PRS construction, and tail threshold.

## Discussion

While GWAS identifies associated loci with high statistical confidence, the majority of lead variants reside in non-coding regulatory regions whose effector genes, relevant cell types, and downstream regulatory targets cannot be determined from variant-level associations alone. TWAS partially addresses this by linking genetically predicted gene expression to trait variation, providing gene-level resolution that is beyond what variant-level GWAS can achieve. However, because TWAS does not distinguish causal genes from those co-regulated with them, signals at a given locus are frequently distributed across correlated predicted expression profiles, limiting biological specificity. Fine-mapping approaches, such as FOCUS, resolve this by computing PIPs over candidate causal transcripts to enable prioritization of the most likely causal gene-tissue or gene-cell type pair at each locus. However, this resolution remains constrained by the tissues and cell types represented in available reference panels.

In this study we conducted a European GWAS meta-analysis of 1,503,740 asthma cases and controls. Integrating this data with the transcriptome-wide association study (TWAS) framework we identified 12,018 and 791 genes at 5% FDR across GTEx tissues and OneK1K cell types, respectively. These genes were enriched for CD4+ Th2 and MHC class II-mediated antigen presentation regulatory programs, which have a well-established role in asthma pathophysiology. We performed gene-level TWAS fine-mapping to account for co-regulation across genes and tissues or cell types. Identifying 4,033 and 1,065 high confidence genes among tissue-level and cell-type-level datasets, we incorporated these genes into polygenic transcriptome risk scores (PTRS). We modeled these prioritized genes across various tissues and cell types using nonlinear classifiers and different integration approaches. Ultimately, we determined that a cross-modal combination of variant-level PRS, tissue-level PTRS, and cell-type-level PTRS resulted in the best performance between our two evaluation cohorts (held-out GACRS and CAMP Balanced). Our work revealed that allowing nonlinear interactions between asthma risk factors improves our ability to distinguish cases from controls. However, while the genome-wide SNP heritability of asthma is limited (even with this work representing the largest asthma GWAS meta-analysis to date), the absolute performance of these models was modest. Rather, the remaining variance of asthma disease status is likely driven by genetic factors not well tagged by common variants and by environmental factors or exposures.

We recognize there are several key limitations of our study. First, we did not model social determinants of health, which are associated with childhood asthma prevalence and disparities^83,84^. Their omission may limit model calibration and portability across GACRS and CAMP. However, our objective was to evaluate the biological resolution and cross-cohort transferability of genetic features, rather than construct a comprehensive clinical risk model. Future studies should integrate genetic, social, and environmental predictors. Second, PTRS models are gene-level linear combinations of largely the same SNPs that are used to construct standard PRS, which inherently limits the possible increase in predictive power. However, we have shown that our PTRS construction allows more appropriate gene-level effect sizes and groupings to be assigned to those SNPs, including non-linear interactions between genes and cell types. In our analysis, the cross-modal PRS-CSx model achieved an AUC of 0.632, compared with 0.514 for PRS-CSx alone, although its improvement over the strongest tissue-level FOCUS PTRS was modest and not statistically significant (ΔAUC = 0.036; P = 0.13). Third, even with multi-ancestry weights provided with PRS-CSx, our TWAS weights and FOCUS causal gene prioritization depends predominantly on European-ancestry GWAS signals through the meta-analysis. As larger population-scale and single-cell eQTL datasets become available in non-European populations, they should provide better-matched inputs for constructing more portable PTRS. Fourth, the optimal protocol for incorporating multiple cell types and tissues into a unified cross-modal risk score remains an open question. As pediatric asthma cohorts become larger and there is more paired single cell and bulk tissue eQTL data, data driven approaches can enhance feature selection procedures. Fifth, our study does not consider how environmental exposures (like viral infections or pollution) or other risk factors like family history influence asthma risk. These are well recognized risk factors, but such characterizations at the individual-level were not present for our cohort subjects^85,86^. Moreover, we did not analyze how these well-known non-genetic risk factors interact with the genetic component of asthma and how much orthogonal variance of disease risk they explain.

Despite these limitations, in this study, we identified tissue-level and cell-type-level features that are most consistent at distinguishing cases from controls between different cohorts with substantial admixture and learned which nonlinear integration schemes optimize their ability to distinguish cases from controls. Our study ultimately revealed that even sophisticated representations of genetic effects and their propagating effects on gene expression only result in modest discrimination between asthma cases and controls. This suggests that in order to build a model that captures the remainder of asthma’s highly complex risk profile, future risk scores should consider other risk factors such as environmental exposures and family history, which play a critical role that may better characterize an individual’s risk.

## Resource availability

### Lead contact

Further information and requests for resources should be directed to and will be fulfilled by the lead contact, Tiffany Amariuta.

### Materials availability

This study did not generate new unique reagents.

### Data and code availability

All original code has been deposited in a public repository made available on GitHub (https://github.com/AmariutaLab/asthma-prs-study-public/) and on Zenodo (https://zenodo.org/records/21342711). URLs and other relevant repositories are listed in the **Key Resources Table.** Any additional information required to reanalyze the data reported in this paper is available from the lead contact upon request.

## Data Availability

All original code has been deposited in a public repository made available on GitHub (https://github.com/AmariutaLab/asthma-prs-study-public/) and on Zenodo (https://zenodo.org/records/21342711). URLs and other relevant repositories are listed in the Key Resources Table. Any additional information required to reanalyze the data reported in this paper is available from the lead contact upon request.

https://github.com/AmariutaLab/asthma-prs-study-public/

## Acknowledgments

This work was supported by funding from the Hartwell Foundation (2023 Individual Biomedical Researcher Award awarded to T.A.), National Science Foundation (NSF) (Award #2336469 awarded to T.A.), and the National Institutes of Health (NIH) (NHGRI R01HG013671 awarded to T.A.). The funders played no role in study design, data collection and analysis, decision to publish or preparation of the manuscript. We would like to thank the GTEx, OneK1K, CAMP, and GACRS cohort participants. The Genotype-Tissue Expression (GTEx) Project was supported by the Common Fund of the Office of the Director of the National Institutes of Health, and by NCI, NHGRI, NHLBI, NIDA, NIMH, and NINDS. GACRS was supported by grants HL04370 and HL66289 from the U.S. National Institutes of Health. We thank all subjects for their ongoing participation in this study. We acknowledge the CAMP investigators, including Dr. Scott Weiss, and the research team, supported by NHLBI, for collection of CAMP Genetic Ancillary Study data. All work on data collected from the CAMP Genetic Ancillary Study was conducted at the Channing Laboratory of the Brigham and Women’s Hospital under appropriate CAMP policies and human subject’s protections. The CAMP Genetics Ancillary Study is supported by U01 HL075419, U01 HL65899, P01 HL083069, R01 HL 086601, R37 HL066289 and T32 HL07427 from the National Heart, Lung and Blood Institute, National Institutes of Health. We acknowledge the GACRS investigators, including Dr. Scott Weiss, and the research team, supported by NHLBI grant R37 HL066289. We wish to acknowledge the investigators at the Channing Division of Network Medicine at Brigham and Women’s Hospital, the investigators at the Hospital Nacional de Niños in San José, Costa Rica and the study subjects and their extended family members who contributed samples and genotypes to the study, and the NIH/NHLBI for its support in making this project possible. Molecular data for the Trans-Omics in Precision Medicine (TOPMed) program was supported by the National Heart, Lung and Blood Institute (NHLBI). Genome sequencing for “NHLBI TOPMed: Childhood Asthma Management Program (CAMP)” (phs001726.v3.p1) was performed at the Northwest Genomics Center (HHSN268201600032I). Genome sequencing for “NHLBI TOPMed: The Genetic Epidemiology of Asthma in Costa Rica” (phs000988.v6.p1) was performed at the Northwest Genomics Center (3R37HL066289-13S1, HHSN268201600032I). Core support including centralized genomic read mapping and genotype calling, along with variant quality metrics and filtering were provided by the TOPMed Informatics Research Center (3R01HL-117626-02S1; contract HHSN268201800002I). Core support including phenotype harmonization, data management, sample-identity QC, and general program coordination were provided by the TOPMed Data Coordinating Center (R01HL-120393; U01HL-120393; contract HHSN268201800001I). We gratefully acknowledge the studies and participants who provided biological samples and data for TOPMed. This work used the Expanse HPC server at the San Diego Supercomputer Center (SDSC) through allocation BIO230210 from the Advanced Cyberinfrastructure Coordination Ecosystem: Services & Support (ACCESS) program, which is supported by National Science Foundation grants #2138259, #2138286, #2138307, #2137603, and #2138296. The graphical abstract was created with the help of Biorender.com under a postdoctoral license for M.F.R.

## Author Contributions

N.H. and T.A. conceived the original study. N.H., M.F.R., and T.A. designed and performed analyses. N.H., M.F.R., and X.G. processed data. M.F.R., N.H., and T.A. interpreted results. M.F.R. and N.H. generated figures. M.F.R., N.H., and T.A. wrote the manuscript. All authors read, revised, and approved the manuscript.

## Declaration of Generative AI and AI-Assisted Technologies

During the preparation of this work, the author(s) used Codex for reference management using article-specific URLs collected by the authors, Claude for validating the reference list, Claude for formatting and testing code in our GitHub repository, and Codex for checking grammar. After using these tools, the authors reviewed and edited the content as needed and take full responsibility for the content of the publication.

## Supplemental Information

Document S1.pdf **Figures S1–S6.**

Document S2.xlsx **Tables S1-S7.**

## STAR★Methods

### Key Resources Table

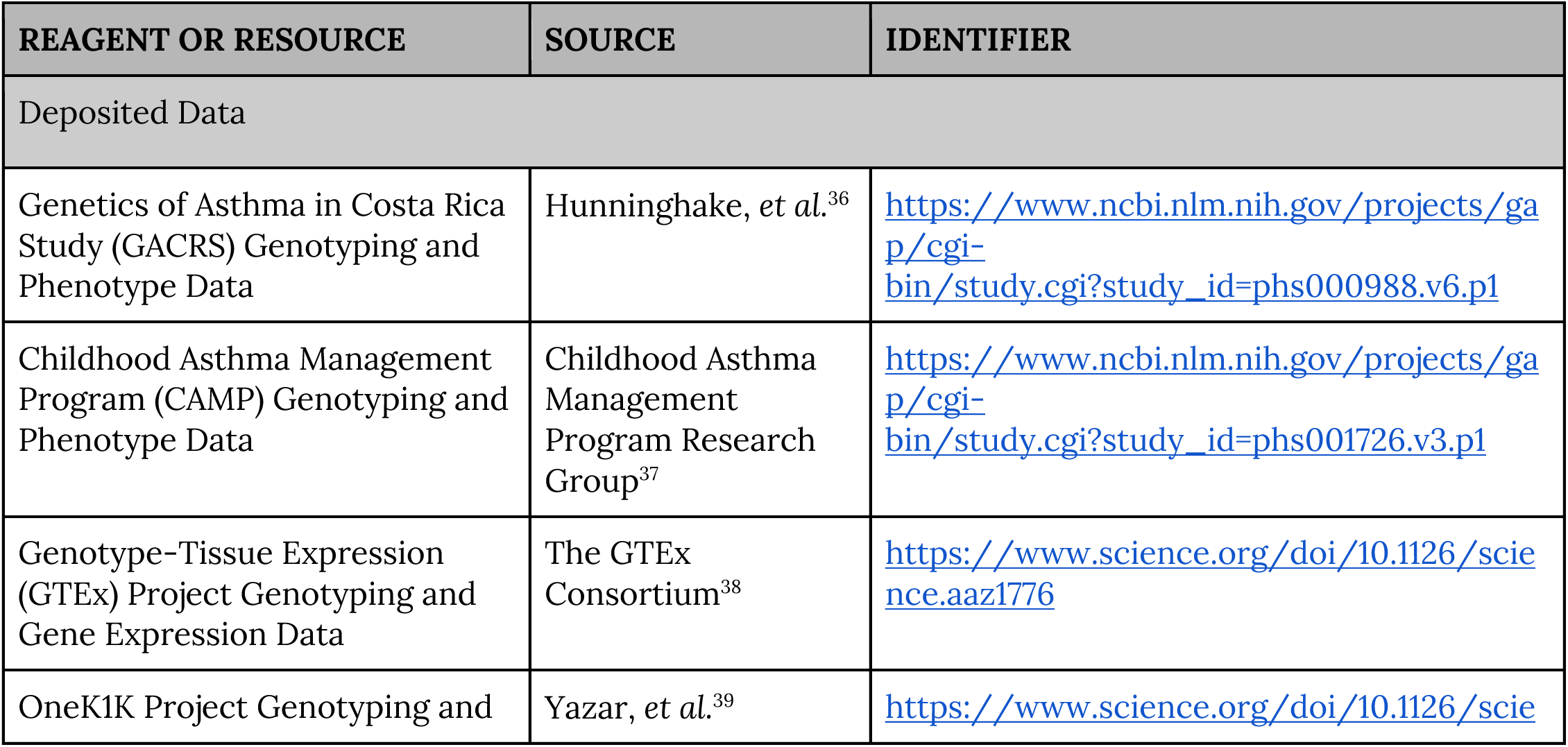
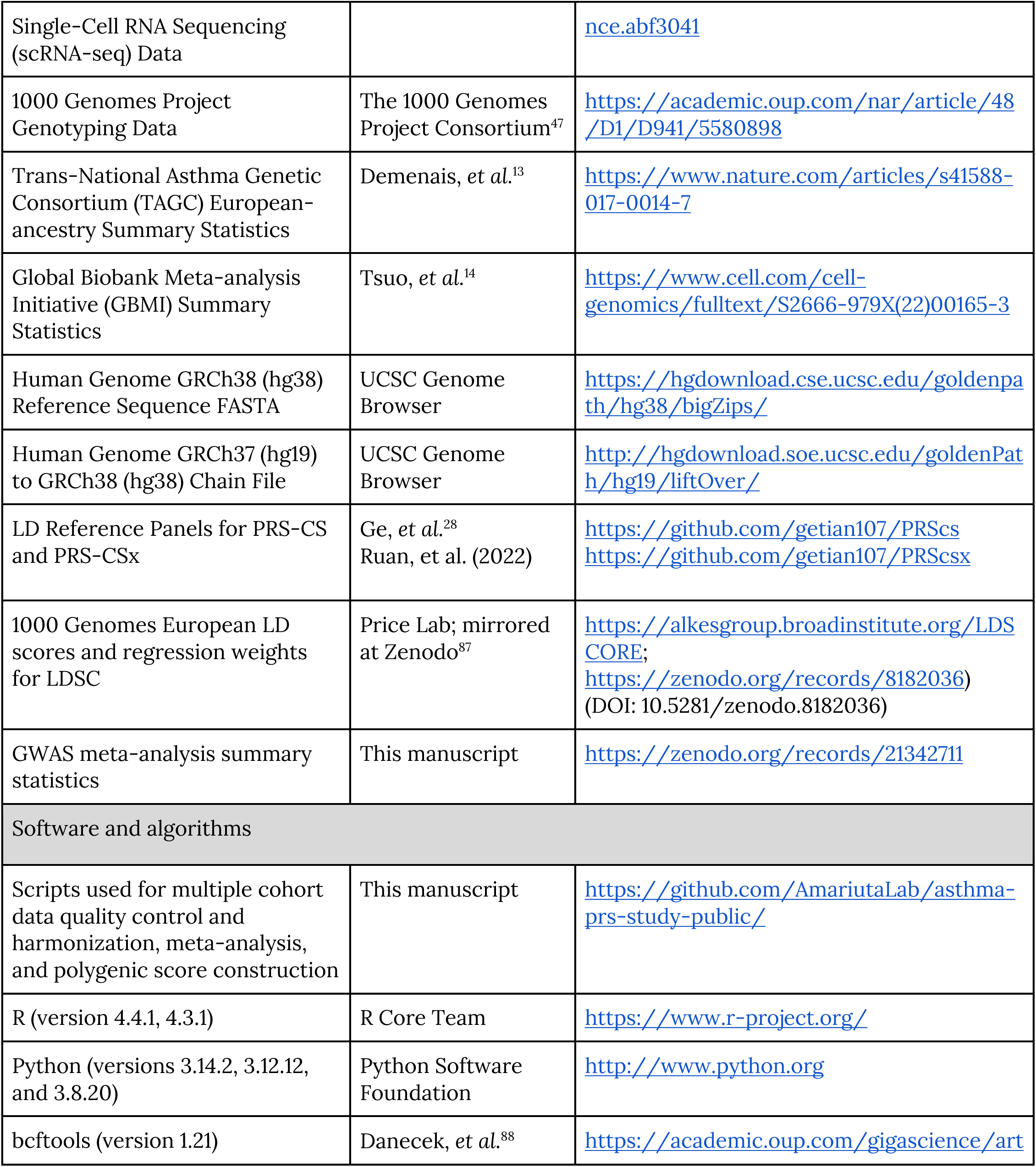
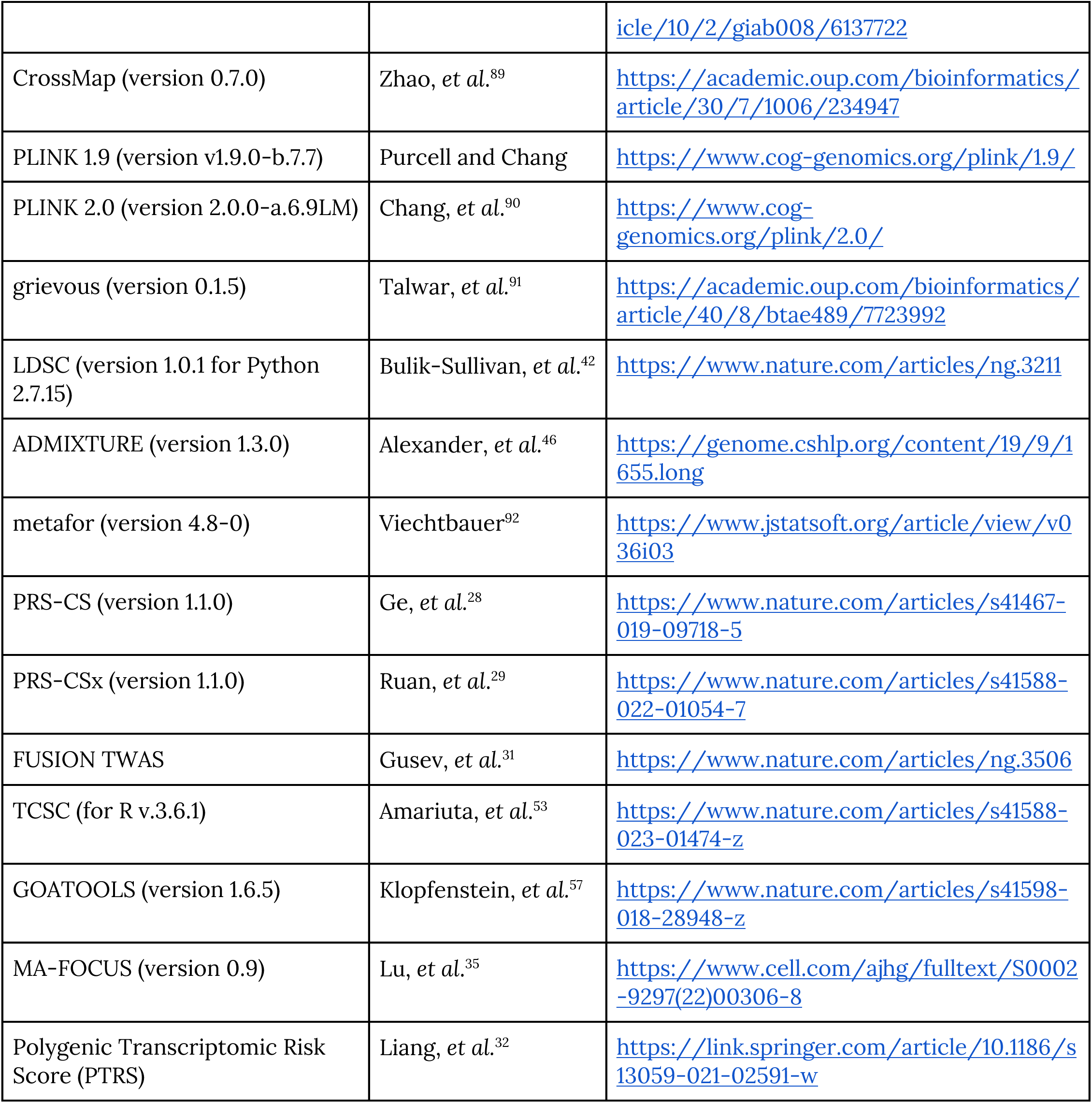

Data sources and access

Individual-level genotyping data for the Adult Genotype-Tissue Expression (GTEx) Project^38^ (https://www.gtexportal.org/home/aboutAdultGtex) were obtained through approved controlled-access requests to the GTEx Consortium through the database of Genotypes and Phenotypes (dbGaP Accession: phs000424.v11.p2). GTEx gene models were obtained from our previous study^53^.

Individual-level genotyping data for the OneK1K Project^39^ were obtained through an approved secondary use of the data obtained through another study^54^. The data was analyzed in accordance with the data use agreements governing the originating study.

Individual-level genotyping data for the 1000 Genomes Project^47^ were obtained from the project’s public data access portal (https://www.internationalgenome.org/data); all data is publicly available and were accessed without restriction.

Individual-level genotyping data for the Genetics of Asthma in Costa Rica Study (GACRS; dbGaP Accession: phs000988.v6.p1)^36^ and the Childhood Asthma Management Program (CAMP; dbGaP Accession: phs001726.v3.p1)^37^ were obtained for this study.

## METHOD DETAILS

### Meta-analysis of European-ancestry asthma GWAS

The Trans-National Asthma Genetic Consortium (TAGC)^13^ comprises individuals of European, African, East Asian, and Latin American ancestry, whereas the Global Biobank Meta-analysis Initiative (GBMI)^14^ includes participants assigned to five major superpopulations present in the 1000 Genomes Project^47^: African (AFR), Admixed American (AMR), East Asian (EAS), European (EUR), and South Asian (SAS) ancestries. In this study, we meta-analyzed European-ancestry genome-wide association study (GWAS) summary statistics from TAGC (19,954 cases, 107,715 controls) and GBMI (121,940 cases, 1,254,131 controls) to create a unified European-ancestry summary statistics dataset for downstream analyses. In total, this dataset included 141,894 cases and 1,361,846 controls (total N=1,503,740 individuals).

Prior to meta-analysis, standard quality-control procedures were applied after merging the summary statistics from each contributing study. These included removal of variants with missing or incomplete effect size or standard error information and harmonization of allele alignment across datasets. After quality control and harmonization, a total of 28,277,483 variants were available across both studies, including 1,616,499 shared variants and study-specific variants unique to each dataset. Of these, 23,828,684 variants with valid allele annotations were retained for downstream analyses.

For variants present in both TAGC and GBMI, we performed a fixed-effect meta-analysis assuming a shared underlying genetic effect across studies with an inverse-variance-weighted (IVW) approach as implemented in *metafor::rma*^92^. For each variant, let β̂_i_ and *SE_i_* denote the estimated effect size and standard error, respectively, from study *i*. Each study was weighted by the inverse of its variance, 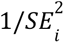. The combined effect size was then computed as:

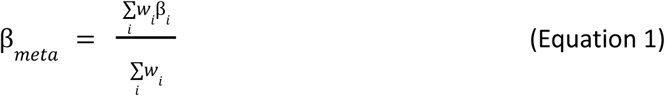

with the corresponding standard error calculated as:

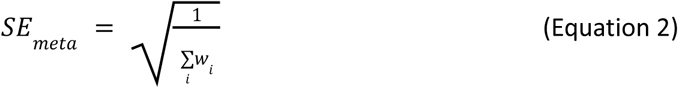

For variants observed in only a single study, we retained the study-specific effect size and standard error without meta-analysis. The resulting summary statistics, therefore, represent a unified set of association results, combining IVW meta-analyzed estimates for shared variants and single-study estimates for study-specific variants, and were used in all downstream analyses.

### SNP-heritability estimation of asthma with LD score regression

We estimated the SNP-based heritability 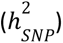 of asthma in our European-ancestry meta-analysis using linkage disequilibrium score regression (LDSC)^42^. Meta-analysis summary statistics were processed using the munge_sumstats.py utility from the LDSC software package (version 1.0.1) built for Python 2.7.15. The effective sample size was specified as the combined number of European-ancestry individuals meta-analyzed across TAGC and GBMI (N=1,503,740). For computational efficiency, we processed data in batches of 500,000 variants using the --chunk-size flag.

We performed SNP-heritability estimation on the liability scale using the --h2 flag. We used precomputed and publicly available European-ancestry LD scores and regression weights for each autosomal chromosome (Key Resources Table). The mean chi-squared statistic (χ²) was 1.795, the LDSC intercept was 1.153 (SE = 0.014), and the ratio was 0.193 (SE = 0.017), indicating that the majority of inflation (approximately 80%) is attributable to real polygenic signal rather than confounding or population stratification. The remaining inflation may represent residual population stratification between TAGC and GBMI, or subtle technical artifacts in the meta-analysis summary statistics. For liability-scale conversion, we specified a sample prevalence (--samp-prev) of 0.0944 and a population prevalence (--pop-prev) of 0.0528. The sample prevalence corresponds to the ratio of asthma cases present in our meta-analysis (141,894 cases, 1,361,846 controls), whereas the population prevalence was based on global estimates of asthma prevalence in European-ancestry individuals^43^, rather than country-specific estimates.

#### Determination of index variants and genome-wide significant loci

To define independent index variants (i.e., “lead” variants) in our combined TAGC-GBMI meta-analysis, we performed linkage disequilibrium (LD)-based clumping in PLINK v1.9 with a European-ancestry LD reference panel constructed from 1000 Genomes Project (Phase 3) individuals. We performed clumping using a genome-wide significance threshold of p ≤ 5 × 10^-8^ to identify index variants, and a secondary threshold of p ≤ 1 × 10^-3^ to assign linked variants to each index variant. We applied an LD threshold of r² = 0.1 within a 1 Megabase (Mb) window. Additionally, we annotated our clumped variants with the effect size (β), standard error (SE), and Z-score statistic (Z) from our meta-analysis. The resulting clumped output was used to define independent genome-wide significant loci for downstream comparison with known asthma-associated variants.

### Pediatric asthma sample selection

Pediatric asthma cases were obtained from two cohorts: the Genetics of Asthma in Costa Rica Study (GACRS)^36^ and the Childhood Asthma Management Program (CAMP)^37^. For both cohorts, we defined pediatric asthma cases as unrelated children with physician-diagnosed asthma; probands were originally selected for these cohorts as previously described in other studies^36,37^. Unrelated child and adult controls without a history of asthma were then selected from the same populations to provide a comparative baseline for downstream analyses. Only one individual per family was included to avoid confounding due to family structure. Following sample pre-filtering, we included 974 cases and 844 controls from the GACRS cohort, as well as 597 cases and 64 controls from the CAMP cohort.

### Data harmonization and pre-processing

Genotype data for the GACRS and CAMP were subjected to quality control and harmonization procedures prior to any downstream analyses.

#### Metadata inspection and sample annotation

Cohort-specific metadata were first reviewed to verify subject identifiers, sample relationships, and phenotype associations, including cases where multiple samples were available per individual subject. Individual-level identifiers, parental identifiers, sex, and phenotype labels were updated and standardized across cohorts. Within-cohort duplicate variants were removed prior to harmonization.

#### Reference genome alignment and variant normalization

To ensure consistency across cohorts, all variant coordinates were aligned to a common reference build (GRCh38). When necessary, genomic coordinates were lifted over from earlier builds with CrossMap^89^. To minimize strand and reference mismatches across cohorts, multiallelic variants were first decomposed into biallelic records with bcftools^88^ (norm), then aligned to the reference genome (+fixref plugin).

Non-ambiguous variants were flipped or swapped as needed, while variants that could not be unambiguously resolved were excluded from downstream steps.

#### Ancestry inference and population structure adjustment

To characterize per-sample ancestry composition and adjust for population stratification, we performed principal component analysis (PCA) for the GACRS and CAMP cohorts by projecting samples onto principal components computed from 1000 Genomes Project reference samples^47^), representing the five major super-populations: African (AFR), Admixed American (AMR), East Asian (EAS), European (EUR), and South Asian (SAS). The resulting per-sample super-population membership scores were used as additive covariates in the variant-level PRS-CS and PRS-CSx baseline logistic regressions (see PRS-CS and PRS-CSx subsections below).

### Ancestry analysis with ADMIXTURE

We estimated individual ancestry proportions for CAMP and GACRS separately with ADMIXTURE, which fits a maximum-likelihood model of *K* ancestral populations to genotype data under the assumption of Hardy-Weinberg equilibrium within populations^46^. Each cohort was separately merged with samples from the 1000 Genomes Project^47^ reference panel through our data harmonization and pre-processing pipeline (see previous subsection). Additionally, we restricted analyses to HapMap 3^93^ SNPs for robust, high-quality inference.

Prior to running ADMIXTURE, we performed quality control and LD pruning separately for each cohort with PLINK 1.9. First, we filtered variants with genotype missingness exceeding 5% (--geno 0.05) or minor allele frequency below 1% (--maf 0.01). Hardy-Weinberg equilibrium filtering was omitted as each cohort had been merged with 1000 Genomes reference samples, for which population stratification and admixture would inflate HWE test statistics and lead to inappropriate variant removal. Finally, we performed LD pruning with a sliding window of 50 variants, a step size of 10 variants, and an r^2^ threshold of 0.1 (--indep-pairwise 50 10 0.1), retaining only the pruned set of approximately independent variants. We then ran ADMIXTURE in the unsupervised mode for values of *K* ranging from 1 to 5. To select the optimal K, we performed 5-fold cross-validation as implemented in ADMIXTURE (--cv). Ancestry results were then used to inform downstream analyses.

#### Polygenic risk score construction

We constructed PRS using two methods of increasing sophistication. First, we applied PRS-CS, a Bayesian continuous shrinkage framework that places a globally learned prior on SNP effect sizes to improve estimation accuracy relative to P+T^28^. We also applied PRS-CSx, which extends PRS-CS to a multi-ancestry framework by jointly modeling effect sizes across populations using coupled continuous shrinkage priors^29^. For both PRS-CS and PRS-CSx, we used pre-computed LD reference panels (Key Resources Table) derived from the 1000 Genomes Project (Phase 3) superpopulations (African (AFR), Admixed American (AMR), European (EUR), East Asian (EAS), and South Asian (SAS)) to ensure compatibility with the ancestrally diverse composition of GACRS and CAMP. The panel choice differs between methods. Global shrinkage parameters were selected through automatic relevance determination (ϕ = auto) as implemented in the PRS-CS and PRS-CSx software^28,29^. For PRS-CS, LDREF = EUR was the only LDREF option available for the single-population model. For PRS-CSx, we selected the cross-population meta-combined LD panel (LDREF = META), consistent with its recommended use for multi-ancestry PRS deployment in cohorts of heterogeneous ancestry composition^29^. Final PRS were computed as the weighted sum of allele dosages across genome-wide variants, with weights derived from the posterior-mean effect size estimates. For downstream integration with PTRS features, both PRS columns were z-standardized using GACRS-train statistics only and merged with the out-of-fold (OOF)-transformed PTRS feature matrices on Sample_ID via inner-join.

#### PRS-CS

PRS-CS was applied to our IVW meta-analysis of individuals from GBMI and TAGC to represent

EUR-ancestry individuals and used the precomputed 1000 Genomes (Phase 3) LD reference panel for EUR-ancestry individuals mentioned above.

After obtaining the posterior effect size estimates, we constructed a PRS for each individual *i* as:

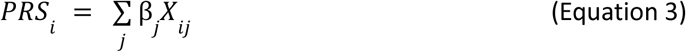

where β̂_j_ denotes the posterior SNP effect size estimate at site *j* produced by PRS-CS and *X_ij_* represents the allele count for individual *i* at site *j*.

#### PRS-CSx

We also computed a multi-ancestry PRS with PRS-CSx, which simultaneously fits multiple sets of input GWAS summary statistics while modeling population-specific LD to produce posterior SNP effect size estimates for each individual population^29^. We supplied available asthma summary statistics from the GBMI cohort for African (AFR), Admixed American (AMR), East Asian (EAS), and South Asian (SAS) individuals. To represent EUR-ancestry individuals, we input our IVW meta-analysis of individuals from GBMI and TAGC.

After obtaining the posterior SNP effect size estimates for each population, we constructed our PRS by training the best linear combination of each single-population PRS that was constructed by PRS-CSx in each cohort as:

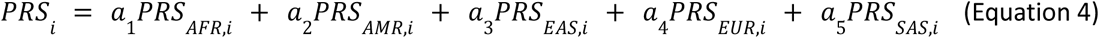

where *a*_1_, *a*_2_, *a*_3_, *a*_4_, and *a*_5_ represent mixing weights and *PRS_AFR_*_,*i*_, *PRS_AMR_*_,*i*_, *PRS_EAS_*_,*i*_, *PRS_EUR_*_,*i*_, and *PRS_SAS_*_,*i*_represent the PRS calculated by PRS-CSx based on the supplied AFR, AMR, EAS, EUR, and SAS summary statistics, respectively, for each individual *i*.

#### PRS Post Processing

Per-chromosome PLINK --score outputs for the default configurations (PRS-CS: ϕ = auto, LDREF = EUR; PRS-CSx: ϕ = auto, LDREF = META) were consolidated into standardized cohort-level tables for downstream classifier-based evaluation. Because PRS-CS and PRS-CSx generate posterior SNP-effect samples through Gibbs sampling, PLINK --score was applied separately to each retained MCMC iteration, yielding chromosome-specific PRS contributions for each individual and iteration. For each method and cohort, SCORESUM values were summed across the 22 autosomes to obtain genome-wide, per-individual, per-iteration PRS values. These data were reshaped to long format using pandas.melt, with one row per individual and MCMC iteration, and joined by IID with sample-level 1000 Genomes superpopulation admixture proportions estimated using ADMIXTURE^46^. The resulting tables served as inputs to downstream PRS evaluations. PRS values were averaged across MCMC iterations to obtain posterior means, standardized using GACRS training-set statistics, and then joined with the PTRS features.

#### PRS-CS and PRS-CSx baseline evaluation

For each of PRS-CS and PRS-CSx, the posterior-mean individual-level PRS described above, standardized using GACRS training-set statistics, was combined with the five 1000 Genomes superpopulation admixture proportions (AFR, AMR, EAS, EUR, and SAS; see *Ancestry inference and population structure adjustment*) to form a six-feature input vector. An L2-regularized logistic regression classifier (scikit-learn LogisticRegression) was trained on the GACRS training set and applied to the held-out GACRS test set and the CAMP-Balanced cohort.

### Transcriptome-wide association study (TWAS) with FUSION

To evaluate gene-level associations from our European-ancestry asthma meta-GWAS, we performed transcriptome-wide association studies (TWAS) using the FUSION framework^31^ in summary statistics mode. In this mode, FUSION does not impute genetically regulated expression in individual participants; rather, for each gene-tissue or gene-cell-type pair, it combines a precomputed *cis*-genetic gene expression prediction model with the GWAS meta-analysis effect sizes and a reference LD panel to estimate the association between the gene’s genetically regulated expression and asthma risk^31^. We applied FUSION to two complementary precomputed *cis*-eQTL prediction-weight panels: (1) 39 GTEx v8 bulk tissues from postmortem donors including asthma-relevant tissues (lung, esophagus mucosa, esophagus muscularis, whole blood) for which we previously computed FUSION gene model weights^53^; and (2) 17 OneK1K pseudobulked peripheral blood mononuclear cell (PBMC) types, using cell-type-specific FUSION weights^39,54^. Briefly, we performed log_2_(TPM+1) normalization before pseudobulking single-cell gene expression profiles across donors and cell types and then standardized gene expression to be compatible with FUSION. TPM (transcripts per million) refers to the process of normalizing each gene’s expression by its length in kilobases and then using a scaling factor such that each cell’s total expression sums to one million^95^. OneK1K has a larger donor sample size than per-tissue GTEx (982 donors vs. 320 for most analyzed GTEx tissues) but generally weaker gene expression prediction models, due to the sparsity and noise of single-cell expression data, including highly variable numbers of cells per donor; this asymmetry is empirically reflected in the number of TWAS associations detected by each modality.

By default, FUSION filters out genes whose expression levels are not *cis*-heritable (p > 0.01), determined by GCTA^94^. We additionally require that the heritability estimate be non-negative. Then, for each *cis*-heritable gene-tissue or gene-cell-type pair, FUSION generates weight files containing the default four candidate *cis*-eQTL prediction models: top1, defined as a single SNP model with the largest squared effect size; LASSO regression; elastic net regression; and best linear unbiased prediction (BLUP). Each file also includes cross-validation performance statistics for the available models. For each gene, FUSION selects the candidate model with the lowest cross-validation p-value for association with the trait, while genes whose minimum p-value is > 0.05 are excluded from further analysis, ensuring that all reported TWAS Z-scores derive from genes whose expression can be nominally explained by *cis*-genetic variation.

The European GWAS meta-analysis summary statistics were used as input to both tissue- and cell-type-specific TWAS panels. SNP-level LD was modeled using the 1000 Genomes Project (Phase 3) European reference panel distributed with FUSION (LDREF/1000G.EUR.*). The resulting per-gene, per-context (tissue or cell type) TWAS Z-scores (and corresponding two-sided p-values) were then carried forward to downstream quality-control filtering, probabilistic fine-mapping with FOCUS, and the alternative LD-clumping + p-value-thresholding gene-prioritization pipeline.

### Gene Ontology (GO) term enrichment analysis with GOATOOLS

To confirm the biological relevance of FUSION TWAS-identified gene sets prior to probabilistic fine-mapping with FOCUS, we performed Gene Ontology (GO) term enrichment analysis with GOATOOLS^55–57^ separately for genes identified from gene-tissue and gene-cell-type associations.

Gene-tissue and gene-cell-type association p-values from FUSION TWAS were corrected for multiple testing using the Benjamini-Hochberg false discovery rate (FDR) procedure applied separately within each modality. From 226,122 tested gene-tissue pairs, there were 24,866 unique genes, and from 19,055 tested gene-cell-type pairs, there were 3,571 unique genes. Unique genes with FDR < 5%, e.g., dropping tissue or cell type pairing, were used as the study set for enrichment analysis: 3,524 for tissue and 479 for cell types. The background gene set for each enrichment analysis was restricted to all genes tested in FUSION TWAS within the corresponding modality rather than the full set of protein-coding human genes to avoid inflating enrichment statistics with genes never evaluated by TWAS.

GO annotations were obtained from the National Center for Biotechnology Information (NCBI) gene2go database for Homo sapiens (Taxonomy ID 9606), and the GO directed acyclic graph (DAG) was parsed from the resulting go-basic.obo file^55,56^. Enrichment significance was assessed using a two-sided Fisher’s exact test with Benjamini-Hochberg correction across all tested GO terms as implemented in GOATOOLS^57^. Enriched terms with FDR < 5% were retained for further interpretation.

### Identification of tissue- and cell-type-specific genes with asthma (FOCUS / MA-FOCUS)

We leverage gene fine-mapping with the single-ancestry FOCUS mode^34^ from a locally modified fork of the MA-FOCUS codebase^35^. FOCUS assumes shared causal genes across populations, enhancing fine-mapping accuracy and enabling cross-ancestry transferability of gene-level signals. A subset of our FUSION weight panels stores only a single model per gene and omits the cross-validation performance table that FOCUS uses to choose among multiple candidate models. This subset corresponds to the meta-analyzed bulk-tissue panels^53^. Original FOCUS fails to import these panels because it assumes every weight file carries the standard multi-model FUSION layout. We therefore made a minor modification to FOCUS such that it accepts this single-model format; weight files with the standard multi-model layout continue to be handled by the original code path. The modification does not alter any downstream FOCUS behavior, including region selection, cross-population fine-mapping, or credible-set construction. The modified version is available within our analysis code.

For GTEx tissue-level feature selection, we retained gene tissue pairs with PIP ≥ 0.1, approximately the upper quartile of PIPs among the 17,712 pairs in GTEx 90% credible sets. This yielded 4,033 final retained pairs. For OneK1K, we retained all 1,065 gene cell-type pairs in the 90% credible sets without applying an additional PIP threshold. This asymmetric criterion reflects the substantially smaller OneK1K feature pool. Applying PIP ≥ 0.1 to OneK1K would have retained only 688 pairs, leaving several cell types with too few features for stable downstream PTRS construction.

### Building LD clumping and p-value thresholding-based polygenic risk score models

For each gene-context pair that passed the heritability filter, we obtained the TWAS Z-score *Zt*_,*g*_ from the FUSION summary-statistics association and the genetically predicted expression vector Ê*t*_,*g*_ (*j*) across individuals *j* in the target cohort, where *t* indexes tissues and *g* indexes genes, and individual genotypes *Xt*_,*g*_(*j*) were used to predict gene expression using the same eQTL weights *wt*_,*g*_ used in the TWAS step:

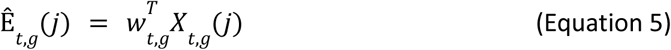

Because predicted expression of nearby genes becomes correlated through shared eQTLs and LD, we performed the equivalent of LD clumping across genes within each tissue using the empirical Pearson correlation of Ê*t*,*g* across GACRS individuals using r² < 0.1, keeping the gene-tissue pair with the smallest TWAS p-value in each correlated block. We then thresholded the clumped tissue-specific gene list at four TWAS p-value cutoffs θ ∈ {5 × 10^-5^, 5 × 10^-4^, 5 × 10^-3^, 0.05}, yielding tissue-specific and threshold-specific gene sets *St*,θ .

For each *(t, θ),* we computed a Z-weighted polygenic transcriptomic risk score, as previously done in Liang et al., 2022:

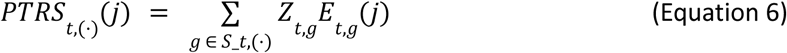

implemented in our analysis pipeline as the per-individual scores produced by the PTRS construction script.

We split the GACRS cohort into 60/20/20 training, validation, and test sets. The validation split was drawn first (20%, random seed 42); the remaining 80% was further split 75/25 into training and test sets (random seed 0). The per-individual *PTRSt*_,θ_ was computed once per (t, θ) for every sample using the fitted FUSION eQTL weights across all three GACRS splits (train, validation, and test) and the external CAMP-Balanced cohort. There is no cohort-specific fitting at this step.

Feature selection then proceeded in two stages. First, for each (t, θ), we retained only those features and configurations with higher mean PTRS in cases than in controls in the GACRS validation set. Second, a cross-cohort consistency filter was applied to the surviving configurations to identify (t, θ) pairs with stable performance across cohorts (specific criteria detailed in Two-stage modality-specific PTRS selection and evaluation), producing the final (t, θ) shortlist used to compare with the FOCUS PTRS. The primary evaluation metric used throughout the study is the area under the curve (AUC); for CAMP-Balanced, we report the mean AUC ± SD across 100 case-matched bootstraps. This matches the linear-model TWAS P+T formulation used in the original PTRS framework^32^.

### Theoretical common variant AUC ceiling

We estimated the theoretical maximum AUC attainable by a predictor that perfectly captures the common-variant genetic liability to asthma using the liability-threshold framework of Wray et al. (2010).

Let *K* denote disease prevalence, *T* = Φ^−1^ (1 − *K*) the liability threshold, and **φ**(*T*) the standard normal density at *T*. The standardized selection intensities for cases and controls are:

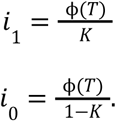

Assuming a standardized liability with common variant SNP heritability 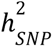, the conditional means of genetic liability among cases and controls are:

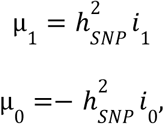

and their corresponding variances are:

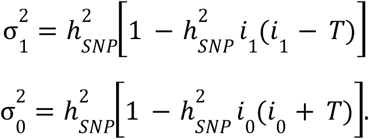

The maximum AUC is therefore:

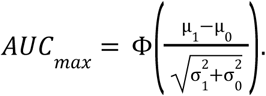

Using the population prevalence applied in our LDSC analysis (*K* = 0. 0528) gives *T* = 1. 6183, φ(*T*) = 0. 1077, *i*_1_ = 2. 0398, and *i*_0_ = 0. 1137. With 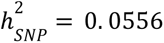, the resulting conditional parameters are µ_1_ = 0. 1134, µ_0_ =− 0. 0063, 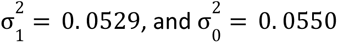. Consequently:

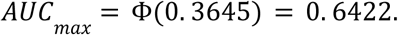

Propagating the 95% confidence interval of the LDSC heritability estimate alone (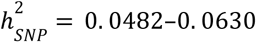) produced an approximate AUC-ceiling interval of 0.633–0.651. This interval does not incorporate uncertainty in population prevalence or deviations from the liability-threshold assumptions. The estimate should therefore be interpreted as an approximate model-based benchmark rather than a fixed empirical upper bound, particularly because the heritability estimate was derived from an all-age European-ancestry asthma meta-analysis whereas model performance was evaluated in pediatric cohorts.

### Assembling an asthma phenotype-aware polygenic transcriptome risk score

The variant-level PRS-CS and PRS-CSx baselines summarize the marginal SNP-level signal under a Bayesian shrinkage prior, but do not leverage how these variants may impact gene expression regulation, which has been proposed to represent more transferable features across cohorts^32^. To this end, we built per-individual Polygenic Transcriptome Risk Scores (PTRS) in which an individual’s genetic risk for asthma is estimated by the TWAS-Z-weighted sum of genetically predicted expression across a prioritized gene set, separately within each tissue and cell type.

Our baseline gene-level PTRS approach was the TWAS LD clumping + p-value thresholding (TWAS P+T) approach described above. Our first proposed prioritization strategy used FOCUS probabilistic fine-mapping. GTEx tissue gene models were retained if they were included in a 90% credible set and had PIP ≥ 0.1, whereas OneK1K gene cell-type models were retained based solely on inclusion in a 90% credible set. We ensured that both approaches used the same input gene-level features, including a consistent best FUSION model per gene-context pair, and followed the same gene expression prediction approach described above when estimating Ê*t*_,*g*_(*j*). We further required that the variance of the predicted expression value must be nonzero across individuals within each cohort. Then, we either applied the pruning and thresholding procedure described above for the baseline approach or the FOCUS PIP and credible set thresholding for the proposed approach. Each strategy resulted in a different set of gene models supplied to Equation 6 above. Specifically, we use the FUSION SCORESUM column from the PLINK profile scoring procedure and perform gene-wise multiplication with the TWAS Z-score and sum these values across the model-specific gene set. Any non-additive structure or feature-specific calibration is learned downstream by binary classifiers, such as random forest.

For the GTEx tissue panel, these steps yield a per-individual PTRS vector of length 39 tissues, and for the OneK1K cell type panel, the vector is of length 17 cell types. These per-feature PTRS values are computed once on every individual in GACRS (across all three train/validation/test splits) and on every individual in the external CAMP evaluation cohort, applying the same per-feature gene sets and the same FUSION eQTL weights to every cohort learned from the European asthma GWAS meta-analysis. We emphasize that these gene-level features were never determined based on any information from the GACRS or CAMP cohorts.

### Two-stage modality-specific PTRS selection and evaluation

The per-feature PTRS catalogue included 39 GTEx tissues and 17 OneK1K cell types, evaluated across FOCUS and TWAS P+T strategies. For TWAS P+T, four p-value thresholds were tested. This catalogue was reduced to a shortlist through a directional filter in Stage 1 and a cross-cohort AUC consistency filter in Stage 2.

Stage 1 applied a case-and-control directionality filter. For each (t, θ) configuration, we retained only features with a higher mean PTRS in cases than in controls in the GACRS validation set. The retention was based solely on the direction of the case and control mean difference.

Stage 1 used a cross-cohort consistency filter defined in the section “Building LD clumping and p-value thresholding-based polygenic risk score models.”

Stage 2 applied a cross-cohort AUC consistency filter. Stage 1 surviving features were required to achieve AUC > 0.53 in GACRS-test and mean AUC > 0.52 in CAMP-Balanced, calculated across 100 bootstrap iterations of 64 cases and 64 controls. Both thresholds were set slightly above the null AUC of 0.50; the lower CAMP-Balanced threshold accommodates the greater sampling variability of its smaller evaluation samples. Requiring both criteria prioritizes features showing concordant discrimination across two demographically distinct cohorts while retaining features with modest individual effects.

For TWAS P+T configurations, candidate classifiers were restricted to penalized linear models, consistent with the linear scoring formulation of the original PTRS framework^32^. We evaluated ridge logistic regression at three regularization strengths C ∈ {0.01, 0.1, 1.0}, LASSO-regularized logistic regression at C = 0.1, and elastic-net logistic regression tuned via 5-fold cross-validation over the mixing parameter L1_ratio ∈ {0.1, 0.3, 0.5, 0.7, 0.9} and regularization strength C ∈ {0.001, 0.01, 0.1, 1.0}. C denotes the inverse-regularization parameter in scikit-learn, with smaller values corresponding to stronger regularization. All classifiers used class_weight="balanced". Restricting TWAS P+T models to this family maintained consistency with the linear-scoring paradigm in which PTRS was introduced^32^. For FOCUS configurations, we additionally evaluated nonlinear classifiers, including random forest, gradient boosting, and a stacking ensemble with a logistic-regression meta-learner. These models were included to assess whether fine-mapped gene sets contain nonlinear relationships among prioritized gene features that cannot be captured by additive linear models.

To quantify the joint predictive contribution of identified tissues or cell types beyond any single feature and model potential nonlinear relationships among them, we aggregated the MA-FOCUS features passing the Stage 2 filter into per-individual unified PTRS. Separate unified tissue- and cell-type-level PTRS were constructed by fitting the same seven-classifier used at the per-feature level (three ridge logistic regressions, one LASSO logistic, one CV-tuned elastic net, gradient boosting, and a 5-fold cross-validated grid-searched random forest) to the corresponding per-feature scores. We reported the top-performing classifier for each modality (grid-searched random forest for the tissue-level unified PTRS; gradient boosting for the cell-type-level unified PTRS). Furthermore, within the GACRS training set, 5-fold stratified cross-validation with random_state=42 was used to generate out-of-fold predictions.

These out-of-fold predictions provided leakage-free unified PTRS values on the GACRS train as inputs to evaluate the unified PTRS and variant-level PRS integration model.

Per-feature classifiers and the unified PTRS were evaluated across two cohorts that capture distinct forms of generalization. The first was the GACRS-test, the 20% within-cohort test split generated using a stratified random split. The second was CAMP-Balanced, the repeated balanced-subsampling evaluation of the pediatric CAMP cohort described in the final-model evaluation on CAMP. Each CAMP-Balanced iteration comprised 64 sampled cases and all 64 controls. For each strategy, feature space, classifier, and evaluation cohort, we report AUC, top-quartile case enrichment via odds ratio (OR), and case–control Welch’s t-test p-value.

### Comprehensive and cross-modal pediatric asthma genetic risk score

Variant-level PRS and gene-level PTRS capture complementary axes of risk. PRS represents marginal genome-wide SNP-level signal estimated under a Bayesian shrinkage prior, whereas PTRS represents tissue- or cell-type-resolved eQTL-weighted predicted expression. The integration stage combines these complementary signals into a single per-individual risk score. This provided the within-modality model class shown in **Figure 4B**. Building on this baseline, we then tested a cross-modal architecture using the top cell type feature (naïve CD4+ T cell PTRS) and top tissue feature (esophageal mucosa PTRS) and modeled these jointly with z-standardized variant-level PRS from either PRS-CS or PRS-CSx. No further feature selection or reduction was applied.

Each selected feature’s raw PTRS was standardized using GACRS training-set statistics. Scores were then transformed into predicted asthma probabilities using the best per-feature classifier identified during individual feature evaluation. Within GACRS-train, 5-fold stratified cross-validation generated out-of-fold probabilities using sklearn.model_selection.cross_val_predict with shuffled folds and random_state=42. Each sample was predicted by a classifier fitted to the other four folds, preventing its label from being used to fit the model generating its prediction. The resulting per-feature probabilities replaced the raw PTRS columns in downstream cross-modal feature matrices. Accordingly, for every individual *j*, the cross-modal integration input contains the three following features:

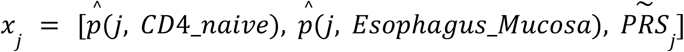

where p̂(*j*, *f*) is the per-feature per-individual predicted probability, and 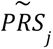 is the z-standardized PRS score (PRS-CS or PRS-CSx). While we did not model ancestry-specific principal components or other covariates at this stage, ancestry is implicitly modeled by the PRS-CSx algorithm when estimating population-specific weights for each input GWAS.

### Integration strategies

For integrating each PTRS configuration (unified-PTRS, cross-modal) with each PRS variant (PRS-CS or PRS-CSx), we trained on the GACRS training samples and evaluated on the GACRS-test and CAMP-Balanced cohorts. All grid searches used 5-fold stratified cross-validation on GACRS-train (sklearn.StratifiedKFold, shuffle = True, random_state = 42). The tree/ensemble strategies used AUC as the selection criterion, while the LogisticRegressionCV Elastic Net used the sklearn default score for its inner C selection. We evaluated seven integration strategies.

First, logistic regression with L2 (ridge) regularization at fixed C = 1.0 (sklearn.LogisticRegression, solver = ’liblinear’) served as the canonical additive baseline to integrate the input features. Second, elastic net logistic regression used a fixed L1/L2 mixing ratio l1_ratio = 0.5, equal LASSO and ridge contributions, with the regularization strength C selected by 5-fold inner cross-validation over the sklearn default log-scale grid (sklearn.LogisticRegressionCV, penalty = ’elasticnet’, solver = ’saga’, cv = 5). Third, Random Forest was grid-searched over n_estimators ∈ {50, 100, 200}, max_depth ∈ {2, 3, 5, None}, min_samples_leaf ∈ {10, 20, 50}, and max_features ∈ {1, ’sqrt’, ’log2’}. Fourth, Gradient Boosting was grid-searched over n_estimators ∈ {50, 100}, max_depth ∈ {2, 3}, and learning_rate ∈ {0.05, 0.1}.

Fifth, a linear support vector machine (sklearn.LinearSVC, fixed C = 0.1) was evaluated with Platt-scaled probability calibration. The raw SVM decision-function output *f(x)* is passed through a logistic sigmoid where *A* and *B* are fitted by maximum likelihood on 3-fold held-out predictions (sklearn.CalibratedClassifierCV, method = ’sigmoid’, cv = 3). Sixth, Stacking-Fixed used a two-layer ensemble in which logistic regression (C = 0.1), Random Forest (n_estimators = 100, max_depth = 3, min_samples_leaf = 20), and Gradient Boosting (n_estimators = 50, max_depth = 2, learning_rate = 0.05) base learners generated 5-fold out-of-fold predicted probabilities, which fed into a logistic-regression meta-learner with solver=’liblinear’ and otherwise default hyperparameters (sklearn.StackingClassifier, base cv = 5). Seventh, Rank-Add was included as a training-free, non-parametric baseline that sums the within-cohort empirical ranks of the input components. This baseline provided a robustness check against integration models that might overfit the small CAMP control set.

The frozen integration models were then evaluated on GACRS-test and CAMP-Balanced, as detailed below.

### CAMP-Balanced evaluation

Final model evaluation used the full pediatric CAMP cohort, comprising 597 cases and 64 controls. Although AUC is invariant to case prevalence, the limited number of CAMP controls motivated a balanced resampling procedure that standardized the evaluation sample size and quantified variability across case subsets. In each of 100 iterations, 64 cases were sampled without replacement and combined with all 64 controls, yielding a balanced sample of 128 individuals. AUC is reported as the mean ± SD across these iterations.

Case–control separation of predicted probabilities was evaluated using Welch’s two-sided t-test (scipy.stats.ttest_ind, equal_var=False) on the full CAMP cohort rather than the balanced subsets. Using the full cohort preserves all available observations and provides greater precision for estimating the difference in mean predicted probability.

Pairwise comparisons used identical CAMP-Balanced subsets across models. For each iteration *i*, we calculated

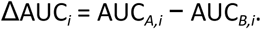

Empirical one-sided resampling support was summarized as the proportion of iterations in which ΔAUC*_i_* ≤ 0. Because the resampled subsets overlap and use the same controls, these quantities describe the consistency of the observed AUC difference across case subsets rather than formal independent-sample P values.

We also calculated top-quartile case enrichment. Within each CAMP-Balanced iteration, the 25th and 75th percentiles of predicted probability defined the bottom and top quartiles. We calculated the case odds ratio as

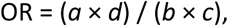

where *a* and *b* are the numbers of cases and controls in the top quartile, and *c* and *d* are the corresponding counts in the bottom quartile. Iterations with no controls in the top quartile or no cases in the bottom quartile produced undefined ORs and were excluded, so no zero-cell correction was applied.

## QUANTIFICATION AND STATISTICAL ANALYSIS

Statistical analyses were performed using the software listed in the Key Resources Table. Figure legends and the main text include the descriptions of statistical tests used to assess the significance of results. Nominal significance refers to p-value < 0.05; Benjamini-Hochberg refers to the calculation of false discovery rate, typically assessed at 5%. Individuals (cases) in the CAMP cohort were randomized to create the CAMP Balanced cohort and bootstrapped versions. Exclusion of individuals based on genetic relatedness is described in the main text and in the Methods above.

