## Supplementary Figures for "Multimodal gene prioritization reveals nonlinear regulatory architecture in childhood-onset asthma"

Supplementary Materials Document

### Multimodal causal gene prioritization and nonlinear tissue and cell-type interaction risk modeling reveal the regulatory architecture of childhood-onset asthma

Nan Huang[*1,2], Michelle F. Ragsac[*1,2], Xiaoyu Gui[1,2], Kelan G. Tantisira[3,4], Tiffany Amariuta[⊗1,2]

*Author Affiliations*

1. Halıcıoğlu Data Science Institute, University of California, San Diego, La Jolla, CA, USA;
2. Division of Biomedical Informatics, Department of Medicine, University of California, San Diego, La Jolla, CA, USA;
3. Division of Pediatric Respiratory Medicine, University of California, San Diego, La Jolla, CA, USA;
4. Department of Pediatrics, Rady Children's Hospital, San Diego, CA, USA;

* These authors contributed equally to this work.

### Supplementary Figures

#### Figure S1. FUSION TWAS Miami plots of gene-tissue associations across bulk GTEx tissues.

Miami plots displaying FUSION TWAS Z-scores across all autosomes for 39 bulk GTEx Project v8 tissues. Each point represents a gene-tissue pair, with alternating silver and dark grey coloring denoting adjacent chromosomes. Highlighted points (teal) exceed the Bonferroni-corrected significance threshold (*p*-value $< 2.21 \times 10^{-7}$) across 226,122 gene-tissue pairs. The dashed navy-blue line indicates $|Z| > 1.96$. Labeled gene names indicate the most significant associations within each tissue.

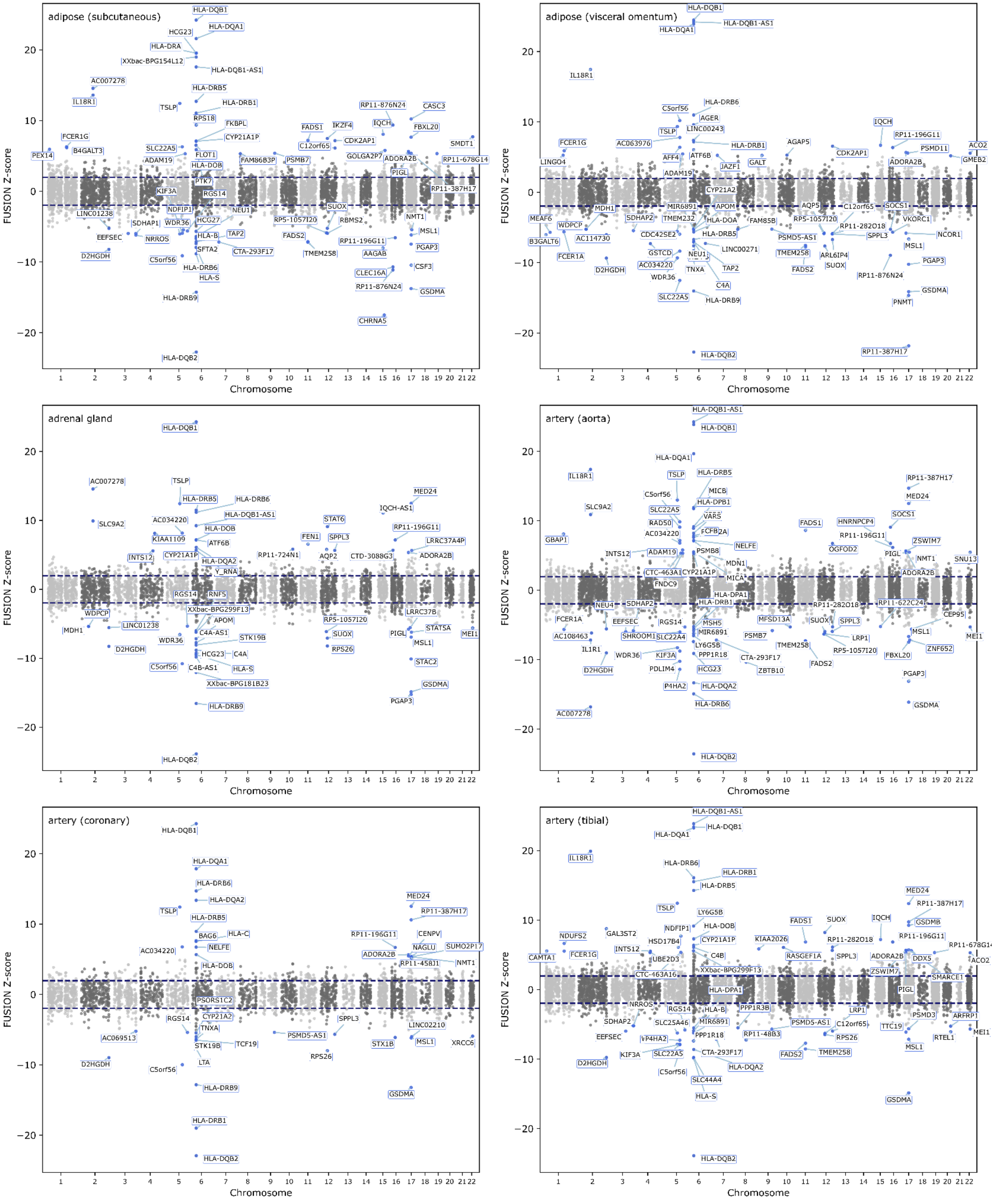

adipose (subcutaneous)
HLA-DQB1
HCG23
HLA-DQA1
HLA-DRA
XXbac-BPG154L12
HLA-DQB1-AS1
AC007278
HLA-DRB5
IL18R1
TSLP
HLA-DRB1
RPS18
RP11-876N24
CASC3
FKBPL
FADS1
IKZF4
IQCH
FBXL20
FCER1G
CYP21A1P
C12orf65
CDK2AP1
SMDT1
B4GALT3
SLC22A5
FLOT1
PEX14
ADAM19
HLA-DOB
FAM86B3P
PSMB7
GOLGA2P7
ADORA2B
RP11-678G14
PIGL
PTK7
KIF3A
RGS14
RP11-387H17
NDFIP1
NEU1
SUOX
LINC01238
NMT1
SDHAP1
WDR36
HCG27
RP5-1057I20
RBMS2
MSL1
EEFSEC
HLA-B
TAP2
FADS2
RP11-196G11
NRROS
PGAP3
SFTA2
CTA-293F17
TMEM258
AAGAB
D2HGDH
C5orf56
CSF3
HLA-DRB6
CLEC16A
HLA-S
RP11-876N24
GSDMA
HLA-DRB9
CHRNA5
HLA-DQB2
FUSION Z-score
Chromosome
adipose (visceral omentum)
HLA-DQB1
HLA-DQB1-AS1
HLA-DQA1
IL18R1
HLA-DRB6
C5orf56
AGER
IQCH
LINC00243
RP11-196G11
TSLP
FCER1G
AC063976
HLA-DRB1
AGAP5
PSMD11
ACO2
CDK2AP1
AFF4
ATF6B
LINGO4
GALT
ADORA2B
GMEB2
JAZF1
ADAM19
CYP21A2
MDH1
MIR6891
APOM
AQP5
C12orf65
SOCS1
MEAF6
SDHAP2
TMEM232
HLA-DOA
FAM85B
RP5-1057I20
VKORC1
WDPCP
HLA-DRB5
RP11-282O18
CDC42SE2
PSMD5-AS1
SPPL3
NCOR1
B3GALT6
AC114730
LINC00271
MSL1
GSTCD
TMEM258
ARL6IP4
FCER1A
NEU1
PGAP3
AC034220
SUOX
D2HGDH
TAP2
FADS2
TNXA
RP11-876N24
WDR36
C4A
GSDMA
SLC22A5
HLA-DRB9
PNMT
HLA-DQB2
RP11-387H17
FUSION Z-score
Chromosome
adrenal gland
HLA-DQB1
AC007278
TSLP
MED24
HLA-DRB5
HLA-DRB6
IQCH-AS1
SLC9A2
AC034220
HLA-DQB1-AS1
STAT6
RP11-196G11
HLA-DOB
FEN1
SPPL3
KIAA1109
LRRC37A4P
ATF6B
RP11-724N1
INTS12
CYP21A1P
AQP2
CTD-3088G3
ADORA2B
HLA-DQA2
Y_RNA
RGS14
RNF5
XXbac-BPG299F13
LRRC37B
WDPCP
APOM
RP5-1057I20
STAT5A
LINC01238
MDH1
C4A-AS1
SUOX
PIGL
MEI1
STK19B
WDR36
MSL1
RPS26
D2HGDH
HCG23
C4A
STAC2
C5orf56
C4B-AS1
HLA-S
XXbac-BPG181B23
GSDMA
PGAP3
HLA-DRB9
HLA-DQB2
FUSION Z-score
Chromosome
artery (aorta)
HLA-DQB1-AS1
HLA-DQB1
HLA-DQA1
IL18R1
TSLP
HLA-DRB5
RP11-387H17
C5orf56
MICB
MED24
SLC9A2
SLC22A5
HLA-DPB1
VARS
SOCS1
FADS1
HNRNPCP4
RAD50
FCFB2A
AC034220
RP11-196G11
GBAP1
ZSWIM7
NELFE
OGFOD2
INTS12
ADAM19
PSMB8
PIGL
NMT1
SNU13
MDN1
CTC-463A1
CYP21A1P
ADORA2B
MICA
FNDC9
HLA-DPA1
NEU4
SDHAP2
HLA-DRB1
RP11-282O18
RP11-622C24
CEP95
FCER1A
EEFSEC
RGS14
MSH5
MFSD13A
SUOX
SPPL3
MSL1
AC108463
SHROOM1
SLC22A4
MIR6891
PSMB7
LRP1
MEI1
IL1R1
LY6G5B
TMEM258
RP5-1057I20
ZNF652
WDR36
KIF3A
PPP1R18
FBXL20
CTA-293F17
D2HGDH
PDLIM4
HCG23
FADS2
ZBTB10
PGAP3
P4HA2
HLA-DQA2
HLA-DRB6
GSDMA
AC007278
HLA-DQB2
FUSION Z-score
Chromosome
artery (coronary)
HLA-DQB1
HLA-DQA1
HLA-DRB6
HLA-DQA2
MED24
TSLP
HLA-DRB5
RP11-387H17
BAG6
HLA-C
RP11-196G11
CENPV
AC034220
NELFE
NAGLU
SUMO2P17
ADORA2B
RP11-458J1
NMT1
HLA-DOB
PSORS1C2
CYP21A2
SPPL3
RGS14
TNXA
LINC02210
AC069513
PSMD5-AS1
STX1B
MSL1
XRCC6
STK19B
TCF19
RPS26
LTA
D2HGDH
C5orf56
HLA-DRB9
GSDMA
HLA-DRB1
HLA-DQB2
FUSION Z-score
Chromosome
artery (tibial)
HLA-DQB1-AS1
HLA-DQA1
HLA-DQB1
IL18R1
HLA-DRB6
HLA-DRB1
HLA-DRB5
MED24
RP11-387H17
TSLP
LY6G5B
FADS1
SUOX
IQCH
GSDMB
HLA-DOB
GAL3ST2
NDFIP1
RP11-196G11
NDUFS2
HSD17B4
CYP21A1P
KIAA2026
RP11-282O18
RP11-678G14
FCER1G
INTS12
C4B
RASGEF1A
SPPL3
ADORA2B
DDX5
CAMTA1
UBE2D3
ACO2
ZSWIM7
XXbac-BPG299F13
CTC-463A16
SMARCE1
HLA-DPA1
PIGL
NRROS
RGS14
HLA-B
PPP1R3B
LRP1
SDHAP2
SLC25A46
MIR6891
PSMD5-AS1
PSMD3
ARFRP1
C12orf65
TTC19
MEI1
EEFSEC
PPP1R18
RP11-48B3
RPS26
VP4HA2
RTEL1
MSL1
KIF3A
SLC22A5
CTA-293F17
FADS2
TMEM258
D2HGDH
HLA-DQA2
C5orf56
SLC44A4
HLA-S
GSDMA
HLA-DQB2
FUSION Z-score
Chromosome

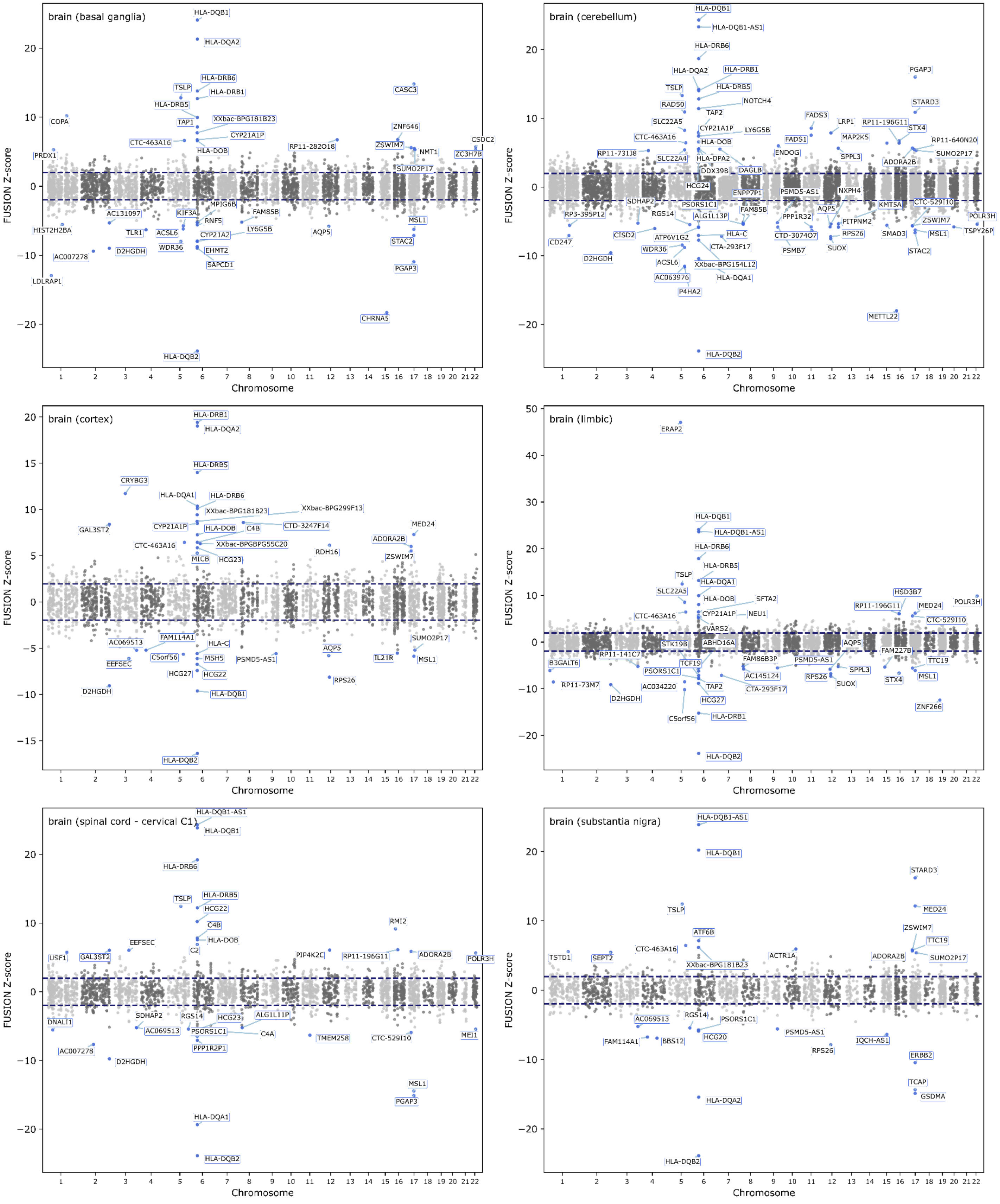
brain (basal ganglia)
HLA-DQB1
HLA-DQA2
HLA-DRB6
TSLP
HLA-DRB1
HLA-DRB5
TAP1
XXbac-BPG181B23
CYP21A1P
CTC-463A16
HLA-DOB
COPA
PRDX1
RP11-282O18
CASC3
ZNF646
ZSWIM7
NMT1
CSDC2
ZC3H7B
SUMO2P17
MPIG6B
KIF3A
AC131097
RNF5
FAM85B
LY6G5B
HIST2H2BA
TLR1
ACSL6
CYP21A2
WDR36
D2HGDH
EHMT2
SAPCD1
AC007278
LDLRAP1
AQP5
MSL1
STAC2
PGAP3
CHRNA5
HLA-DQB2
brain (cerebellum)
HLA-DQB1
HLA-DQB1-AS1
HLA-DRB6
HLA-DQA2
HLA-DRB1
TSLP
HLA-DRB5
RAD50
NOTCH4
TAP2
SLC22A5
CYP21A1P
LY6G5B
CTC-463A16
HLA-DOB
RP11-731J8
SLC22A4
HLA-DPA2
DDX39B
DAGLB
HCG24
ENPP7P1
SDHAP2
PSORS1C1
RGS14
ALG1L13P
FAM85B
RP3-395P12
CISD2
ATP6V1G2
HLA-C
CTA-293F17
CD247
WDR36
D2HGDH
ACSL6
XXbac-BPG154L12
AC063976
HLA-DQA1
P4HA2
HLA-DQB2
FADS3
FADS1
ENDOG
LRP1
MAP2K5
RP11-196G11
STX4
SPPL3
PSMD5-AS1
NXPH4
AQP5
PPP1R32
CTD-3074O7
PSMB7
RPS26
SUOX
PITPNM2
KMT5A
SMAD3
PGAP3
STARD3
RP11-640N20
SUMO2P17
ADORA2B
CTC-529I10
ZSWIM7
POLR3H
MSL1
TSPY26P
STAC2
METTL22
brain (cortex)
HLA-DRB1
HLA-DQA2
HLA-DRB5
CRYBG3
HLA-DQA1
HLA-DRB6
XXbac-BPG181B23
XXbac-BPG299F13
GAL3ST2
CYP21A1P
HLA-DOB
C4B
CTD-3247F14
MED24
CTC-463A16
XXbac-BPGBPG55C20
ADORA2B
RDH16
MICB
HCG23
ZSWIM7
FAM114A1
HLA-C
SUMO2P17
AC069513
C5orf56
MSH5
PSMD5-AS1
AQP5
TL2IR
MSL1
EEFSEC
HCG27
HCG22
RPS26
D2HGDH
HLA-DQB1
HLA-DQB2
brain (limbic)
ERAP2
HLA-DQB1
HLA-DQB1-AS1
HLA-DRB6
HLA-DRB5
TSLP
HLA-DQA1
SLC22A5
HLA-DOB
SFTA2
CYP21A1P
NEU1
CTC-463A16
VARS2
STK19B
ABHD16A
RP11-141C7
TCF19
B3GALT6
PSORS1C1
RP11-73M7
AC034220
TAP2
D2HGDH
HCG27
C5orf56
HLA-DRB1
HLA-DQB2
FAM86B3P
AC145124
CTA-293F17
PSMD5-AS1
RPS26
AQP5
SPPL3
SUOX
HSD3B7
RP11-196G11
MED24
POLR3H
CTC-529I10
FAM227B
TTC19
STX4
MSL1
ZNF266
brain (spinal cord - cervical C1)
HLA-DQB1-AS1
HLA-DQB1
HLA-DRB6
TSLP
HLA-DRB5
HCG22
C4B
HLA-DOB
C2
EEFSEC
USF1
GAL3ST2
RMI2
PIP4K2C
RP11-196G11
ADORA2B
POLR3H
DNALI1
SDHAP2
RGS14
HCG23
ALG1L11P
AC069513
PSORS1C1
C4A
TMEM258
CTC-529I10
MEI1
PPP1R2P1
AC007278
D2HGDH
MSL1
PGAP3
HLA-DQA1
HLA-DQB2
brain (substantia nigra)
HLA-DQB1-AS1
HLA-DQB1
STARD3
TSLP
MED24
ZSWIM7
TTC19
ATF6B
TSTD1
SEPT2
CTC-463A16
ACTR1A
ADORA2B
SUMO2P17
XXbac-BPG181B23
AC069513
RGS14
PSORS1C1
HCG20
PSMD5-AS1
FAM114A1
BBS12
IQCH-AS1
RPS26
ERBB2
TCAP
GSDMA
HLA-DQA2
HLA-DQB2
FUSION Z-score
Chromosome

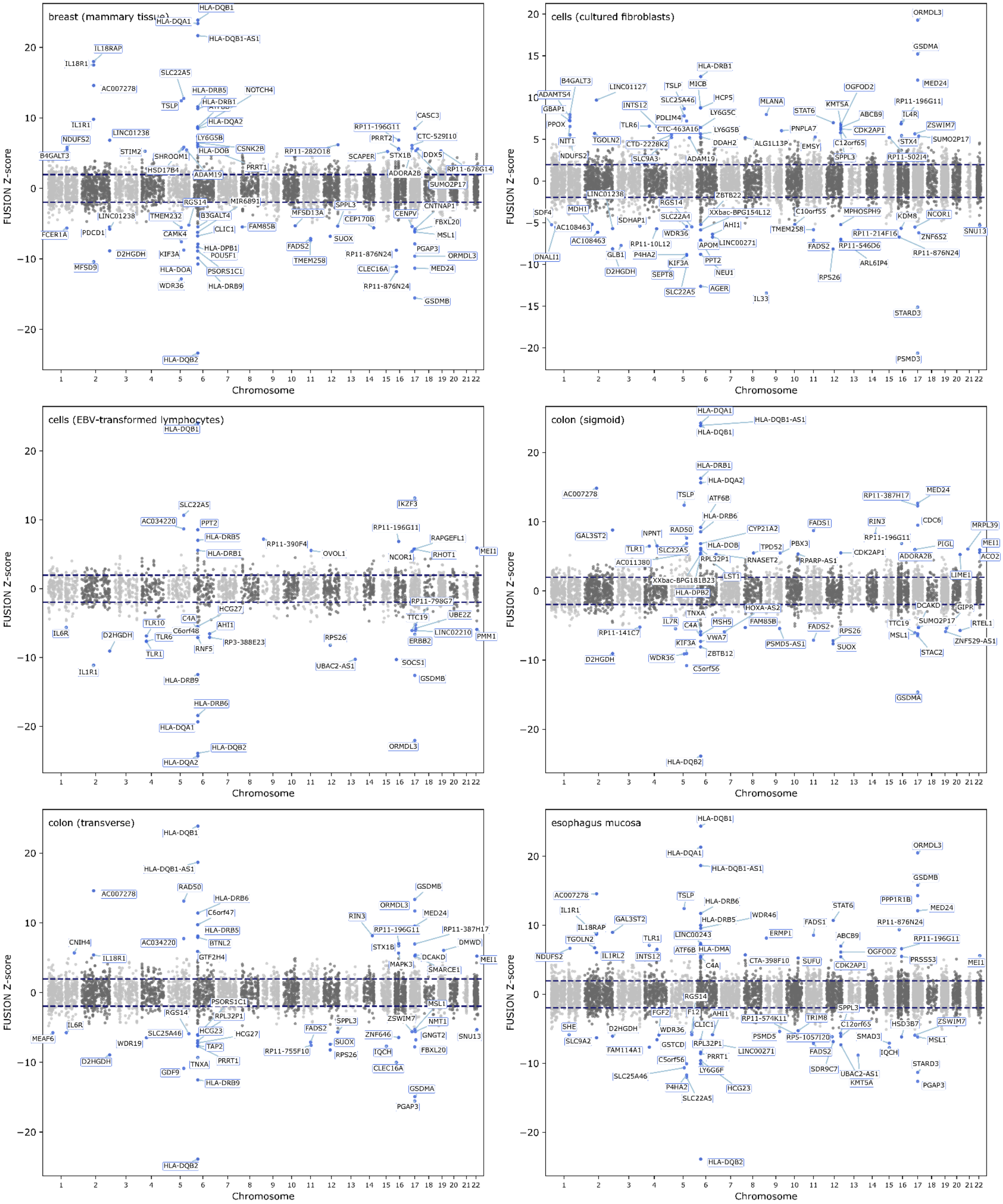

breast (mammary tissue)
cells (cultured fibroblasts)
cells (EBV-transformed lymphocytes)
colon (sigmoid)
colon (transverse)
esophagus mucosa
FUSION Z-score
Chromosome
HLA-DQB1
HLA-DQA1
HLA-DQB1-AS1
HLA-DQB2
HLA-DRB1
HLA-DRB5
HLA-DRB6
HLA-DQA2
ORMDL3
GSDMA
GSDMB
MED24
SLC22A5
TSLP
AC007278
IL18R1
IL18RAP
D2HGDH
PSMD3
STARD3
IKZF3
IL1R1

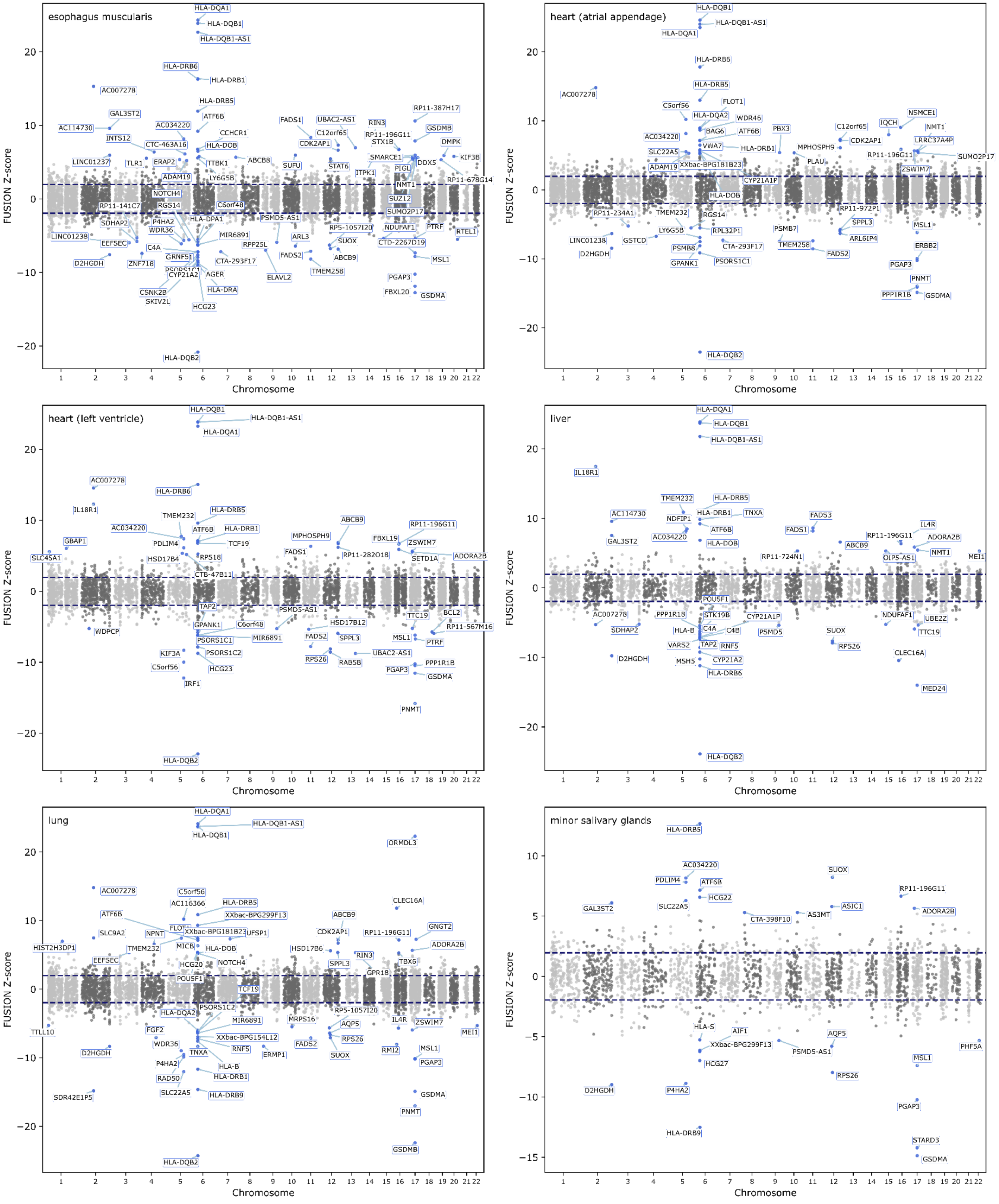

esophagus muscularis
heart (atrial appendage)
heart (left ventricle)
liver
lung
minor salivary glands
FUSION Z-score
Chromosome
HLA-DQA1
HLA-DQB1
HLA-DQB1-AS1
HLA-DRB6
HLA-DRB1
HLA-DRB5
HLA-DQB2
AC007278
GAL3ST2
AC114730
AC034220
D2HGDH
GSDMA
PGAP3
MSL1
PNMT
RP11-196G11
ADORA2B
ATF6B

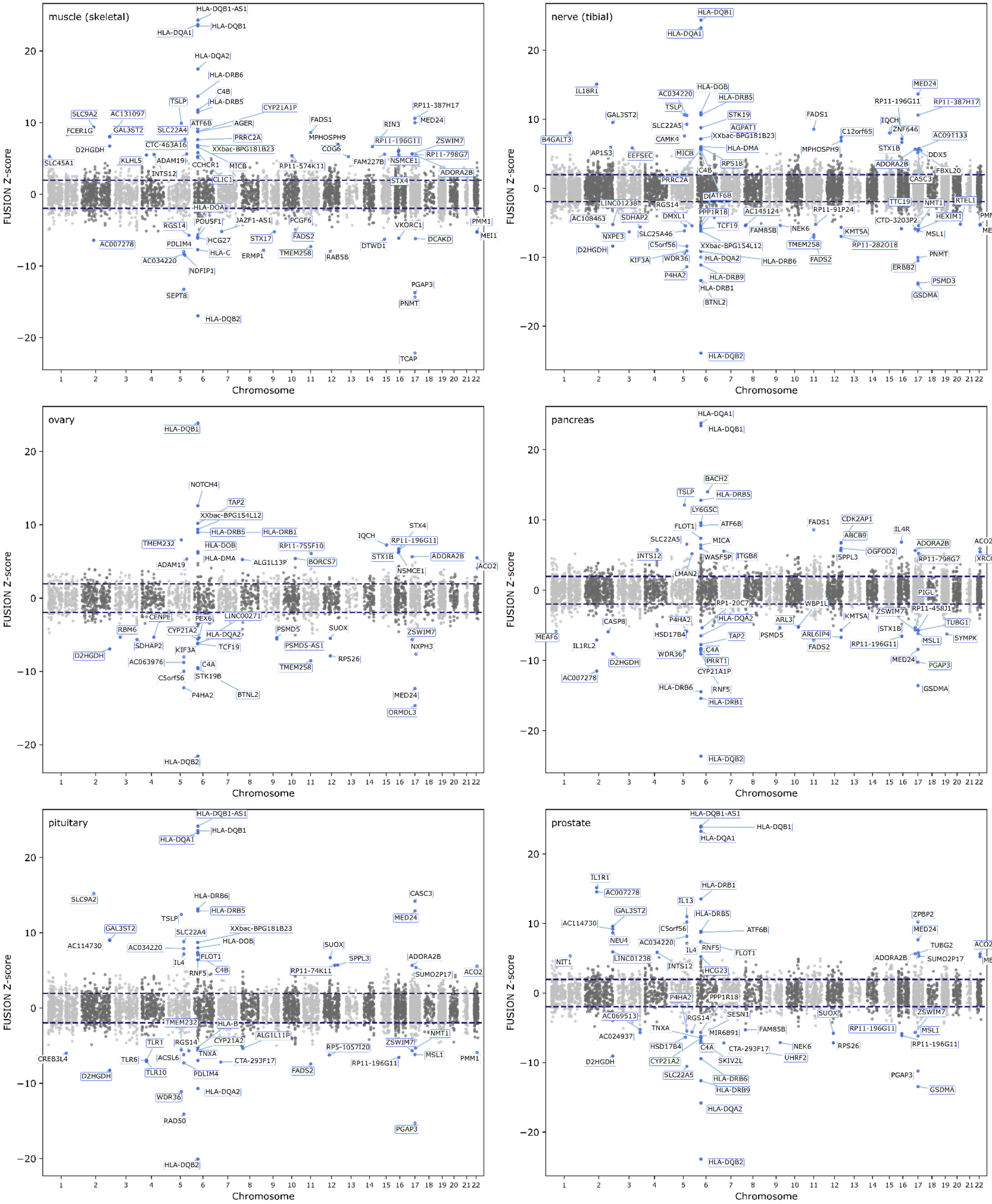
muscle (skeletal)
HLA-DQB1-AS1
HLA-DQB1
HLA-DQA1
HLA-DQA2
HLA-DRB6
C4B
HLA-DRB5
TSLP
SLC9A2
AC131097
FCER1G
GAL3ST2
SLC22A4
ATF6B
AGER
CYP21A1P
PRRC2A
CTC-463A16
XXbac-BPG181B23
D2HGDH
KLHL5
ADAM19
MICB
INTS12
CCHCR1
SLC45A1
CLIC1
FADS1
MPHOSPH9
COG6
RP11-574K11
FAM227B
RIN3
RP11-196G11
NSMCE1
STX4
RP11-387H17
MED24
ZSWIM7
RP11-798G7
ADORA2B
HLA-DOA
POU5F1
JAZF1-AS1
PCGF6
RGS14
HCG27
FADS2
STX17
AC007278
PDLIM4
HLA-C
ERMP1
TMEM258
RAB5B
DTWD1
VKORC1
DCAKD
PMM1
MEI1
AC034220
NDFIP1
SEPT8
PGAP3
PNMT
HLA-DQB2
TCAP
nerve (tibial)
HLA-DQB1
HLA-DQA1
IL18R1
HLA-DOB
AC034220
HLA-DRB5
TSLP
GAL3ST2
STK19
B4GALT3
SLC22A5
AGPAT1
CAMK4
XXbac-BPG154L12
AP1S3
EEFSEC
MICB
HLA-DMA
RPS18
C4B
PRRC2A
FADS1
MPHOSPH9
C12orf65
MED24
RP11-196G11
RP11-387H17
IQCH
ZNF646
AC091133
STX1B
DDX5
ADORA2B
FBXL20
CASC3
LINC01238
RGS14
ATF6B
AC108463
SDHAP2
DMXL1
PPP1R18
AC145124
RP11-91P24
NXPE3
SLC25A46
TCF19
FAM85B
NEK6
KMT5A
TTC19
NMT1
RTEL1
CTD-3203P2
HEXIM1
PMM1
MSL1
MEI1
D2HGDH
C5orf56
XXbac-BPG154L12
TMEM258
RP11-282O18
KIF3A
WDR36
HLA-DQA2
HLA-DRB6
FADS2
ERBB2
PNMT
P4HA2
HLA-DRB9
HLA-DRB1
PSMD3
GSDMA
BTNL2
HLA-DQB2
ovary
HLA-DQB1
NOTCH4
TAP2
XXbac-BPG154L12
HLA-DRB5
HLA-DRB1
TMEM232
HLA-DOB
RP11-755F10
IQCH
STX4
RP11-196G11
STX1B
ADORA2B
HLA-DMA
ALG1L13P
BORCS7
ADAM19
NSMCE1
ACO2
CENPE
LINC00271
PEX6
RBM6
CYP21A2
HLA-DQA2
PSMD5
SUOX
ZSWIM7
SDHAP2
TCF19
PSMD5-AS1
NXPH3
KIF3A
D2HGDH
RPS26
AC063976
C4A
TMEM258
STK19B
C5orf56
BTNL2
P4HA2
MED24
ORMDL3
HLA-DQB2
pancreas
HLA-DQA1
HLA-DQB1
BACH2
TSLP
HLA-DRB5
LY6G5C
FLOT1
ATF6B
FADS1
CDK2AP1
IL4R
SLC22A5
MICA
ABCB9
ADORA2B
ACO2
INTS12
WASF5P
ITGB8
OGFOD2
SPPL3
RP11-798G7
XRCC6
LMAN2
PIGL
RP1-20C7
WBP1L
RP11-458J1
ZSWIM7
KMT5A
CASP8
P4HA2
HLA-DQA2
ARL3
TUBG1
STX1B
HSD17B4
TAP2
PSMD5
ARL6IP4
MEAF6
MSL1
SYMPK
IL1RL2
C4A
FADS2
RP11-196G11
WDR36
D2HGDH
PRRT1
MED24
PGAP3
CYP21A1P
AC007278
HLA-DRB6
RNF5
GSDMA
HLA-DRB1
HLA-DQB2
pituitary
HLA-DQB1-AS1
HLA-DQB1
HLA-DQA1
SLC9A2
HLA-DRB6
CASC3
HLA-DRB5
TSLP
MED24
GAL3ST2
SLC22A4
XXbac-BPG181B23
AC114730
AC034220
HLA-DOB
SUOX
FLOT1
ADORA2B
IL4
SPPL3
C4B
RNF5
RP11-74K11
SUMO2P17
ACO2
TMEM232
HLA-B
TLR1
RGS14
CYP21A2
ALG1L11P
NMT1
ZSWIM7
RP5-1057I20
TNXA
CREB3L4
TLR6
ACSL6
CTA-293F17
MSL1
PMM1
RP11-196G11
FADS2
D2HGDH
TLR10
PDLIM4
WDR36
HLA-DQA2
RAD50
PGAP3
HLA-DQB2
prostate
HLA-DQB1-AS1
HLA-DQB1
HLA-DQA1
IL1R1
HLA-DRB1
AC007278
IL13
GAL3ST2
HLA-DRB5
ZPBP2
AC114730
C5orf56
ATF6B
MED24
NEU4
AC034220
TUBG2
ACO2
RNF5
FLOT1
NIT1
IL4
ADORA2B
SUMO2P17
LINC01238
MEI1
INTS12
HCG23
P4HA2
PPP1R18
ZSWIM7
AC069513
SUOX
RGS14
SESN1
TNXA
FAM85B
RP11-196G11
MSL1
MIR6891
AC024937
HSD17B4
C4A
NEK6
RPS26
CTA-293F17
RP11-196G11
UHRF2
CYP21A2
SKIV2L
D2HGDH
PGAP3
SLC22A5
HLA-DRB6
HLA-DRB9
GSDMA
HLA-DQA2
HLA-DQB2
FUSION Z-score
Chromosome
1 2 3 4 5 6 7 8 9 10 11 12 13 14 15 16 17 18 19 20 21 22
20
10
0
−10
−20

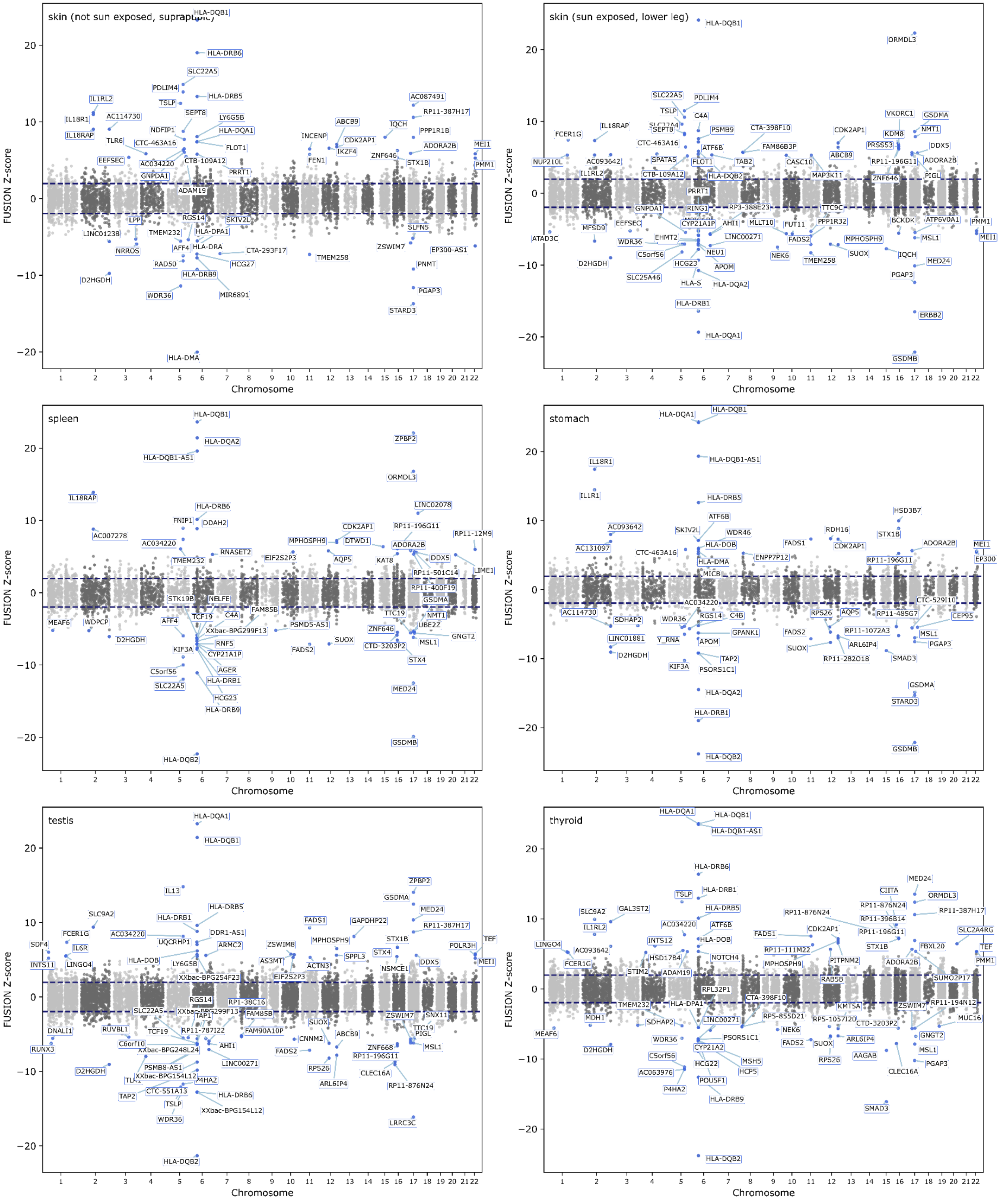
skin (not sun exposed, suprapubic)
skin (sun exposed, lower leg)
spleen
stomach
testis
thyroid
FUSION Z-score
Chromosome
HLA-DQB1
HLA-DRB6
SLC22A5
PDLIM4
HLA-DRB5
IL1RL2
TSLP
IL18R1
AC114730
SEPT8
LY6G5B
NDFIP1
HLA-DQA1
IL18RAP
TLR6
CTC-463A16
FLOT1
EEFSEC
AC034220
CTB-109A12
PRRT1
GNPDA1
ADAM19
RGS14
SKIV2L
LPP
HLA-DPA1
LINC01238
TMEM232
AFF4
HLA-DRA
NRROS
CTA-293F17
RAD50
HCG27
HLA-DRB9
D2HGDH
MIR6891
WDR36
HLA-DMA
INCENP
FEN1
ABCB9
CDK2AP1
IKZF4
TMEM258
AC087491
RP11-387H17
IQCH
PPP1R1B
ADORA2B
ZNF646
STX1B
MEI1
PMM1
SLFN5
ZSWIM7
EP300-AS1
PNMT
PGAP3
STARD3
ORMDL3
PSMB9
CTA-398F10
FAM86B3P
VKORC1
GSDMA
KDM8
NMT1
PRSS53
DDX5
C4A
ATF6B
TAB2
CASC10
MAP3K11
RP11-196G11
ADORA2B
ZNF646
PIGL
FCER1G
NUP210L
AC093642
SPATA5
HLA-DQB2
PRRT1
RING1
RP3-388E23
TTC9C
PPP1R32
BCKDK
ATP6V0A1
CYP21A1P
AHI1
MLLT10
FUT11
MFSD9
ATAD3C
EHMT2
LINC00271
FADS2
MPHOSPH9
MSL1
NEU1
SUOX
NEK6
APOM
TMEM258
MED24
SLC25A46
HLA-S
HLA-DQA2
PGAP3
HLA-DRB1
ERBB2
GSDMB
ZPBP2
HLA-DQA2
HLA-DQB1-AS1
ORMDL3
IL18RAP
LINC02078
FNIP1
DDAH2
RP11-196G11
AC007278
RP11-12M9
MPHOSPH9
DTWD1
RNASET2
EIF2S2P3
AQP5
KAT8
RP11-501C14
LIME1
RP11-400F19
STK19B
NELFE
FAM85B
MEAF6
WDPCP
TCF19
PSMD5-AS1
TTC19
UBE2Z
XXbac-BPG299F13
GNGT2
RNF5
FADS2
CTD-3203P2
KIF3A
CYP21A1P
C5orf56
AGER
HCG23
HLA-DRB9
HLA-DQB2
IL18R1
IL1R1
HSD3B7
AC131097
RDH16
STX1B
HLA-DOB
HLA-DMB
ENPP7P12
MICB
SDHAP2
RPS26
AQP5
RP11-485G7
CTC-529I10
CEP95
GPANK1
RP11-1072A3
LINC01881
Y_RNA
ARL6IP4
TAP2
RP11-282O18
SMAD3
PSORS1C1
SDF4
SLC9A2
FCER1G
IL6R
INTS11
LINGO4
IL13
DDR1-AS1
UQCRHP1
ARMC2
XXbac-BPG254F23
AS3MT
ACTN3
ZSWIM8
GAPDHP22
SPPL3
STX4
NSMCE1
POLR3H
TEF
RP1-38C16
TAP1
RP11-787I22
FAM90A10P
RUVBL1
DNALI1
RUNX3
C6orf10
XXbac-BPG248L24
PSMB8-AS1
XXbac-BPG154L12
P4HA2
CTC-551A13
CNNM2
ZNF668
CLEC16A
RP11-876N24
LRRC3C
GAL3ST2
INTS12
HSD17B4
NOTCH4
STIM2
RPL32P1
RP11-111M22
PITPNM2
RAB5B
SUMO2P17
RP11-194N12
MUC16
KMT5A
RP5-855D21
RP5-1057I20
CIITA
SLC2A4RG
FBXL20
MDH1
MSH5
HCP5
POU5F1
AC063976
AAGAB
SMAD3

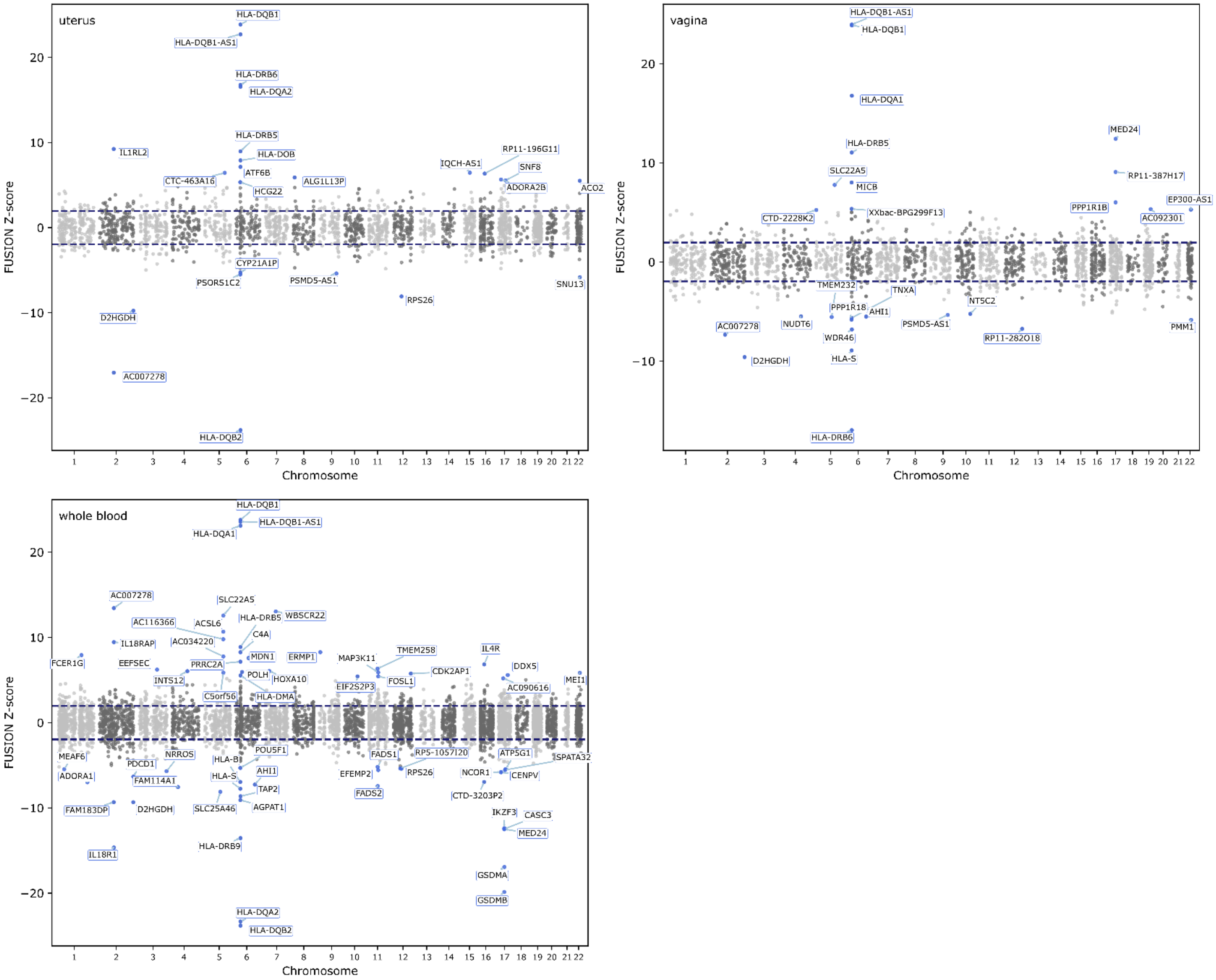
uterus
vagina
whole blood
FUSION Z-score
Chromosome
HLA-DQB1
HLA-DQB1-AS1
HLA-DRB6
HLA-DQA2
HLA-DRB5
HLA-DOB
IL1RL2
CTC-463A16
ATF6B
HCG22
ALG1L13P
IQCH-AS1
RP11-196G11
SNF8
ADORA2B
ACO2
CYP21A1P
PSORS1C2
PSMD5-AS1
RPS26
SNU13
D2HGDH
AC007278
HLA-DQB2
HLA-DQA1
MED24
RP11-387H17
EP300-AS1
PPP1R1B
AC092301
SLC22A5
MICB
XXbac-BPG299F13
CTD-2228K2
TMEM232
TNXA
PPP1R18
AHI1
NT5C2
NUDT6
PSMD5-AS1
WDR46
RP11-282O18
PMM1
HLA-S
AC116366
ACSL6
WBSCR22
IL18RAP
AC034220
C4A
FCER1G
EEFSEC
MDN1
ERMP1
MAP3K11
TMEM258
IL4R
PRRC2A
CDK2AP1
DDX5
INTS12
POLH
HOXA10
FOSL1
MEI1
C5orf56
HLA-DMA
EIF2S2P3
AC090616
MEAF6
PDCD1
NRROS
HLA-B
POU5F1
FADS1
RP5-1057I20
ATP5G1
SPATA32
ADORA1
FAM114A1
EFEMP2
RPS26
NCOR1
CENPV
TAP2
FADS2
CTD-3203P2
FAM183DP
SLC25A46
AGPAT1
IKZF3
CASC3
IL18R1
HLA-DRB9
GSDMA
GSDMB

#### Figure S2. FUSION TWAS Miami plots of gene-cell-type associations across pseudobulked OneK1K PBMC cell types.

Miami plots displaying FUSION TWAS Z-scores across all autosomes for each of the 17 pseudobulked cell types drawn from peripheral blood mononuclear cells (PBMC) from the OneK1K Project cohort. Each point represents a gene-cell-type pair, with alternating silver and dark grey coloring denoting adjacent chromosomes. Highlighted points (teal) exceed the Bonferroni-corrected significance threshold ($p$-value $< 2.92 \times 10^{-6}$) across 19,055 gene-cell-type pairs. The dashed navy-blue line indicates $|Z| > 1.96$. Labeled gene names indicate the most significant associations within each cell type.

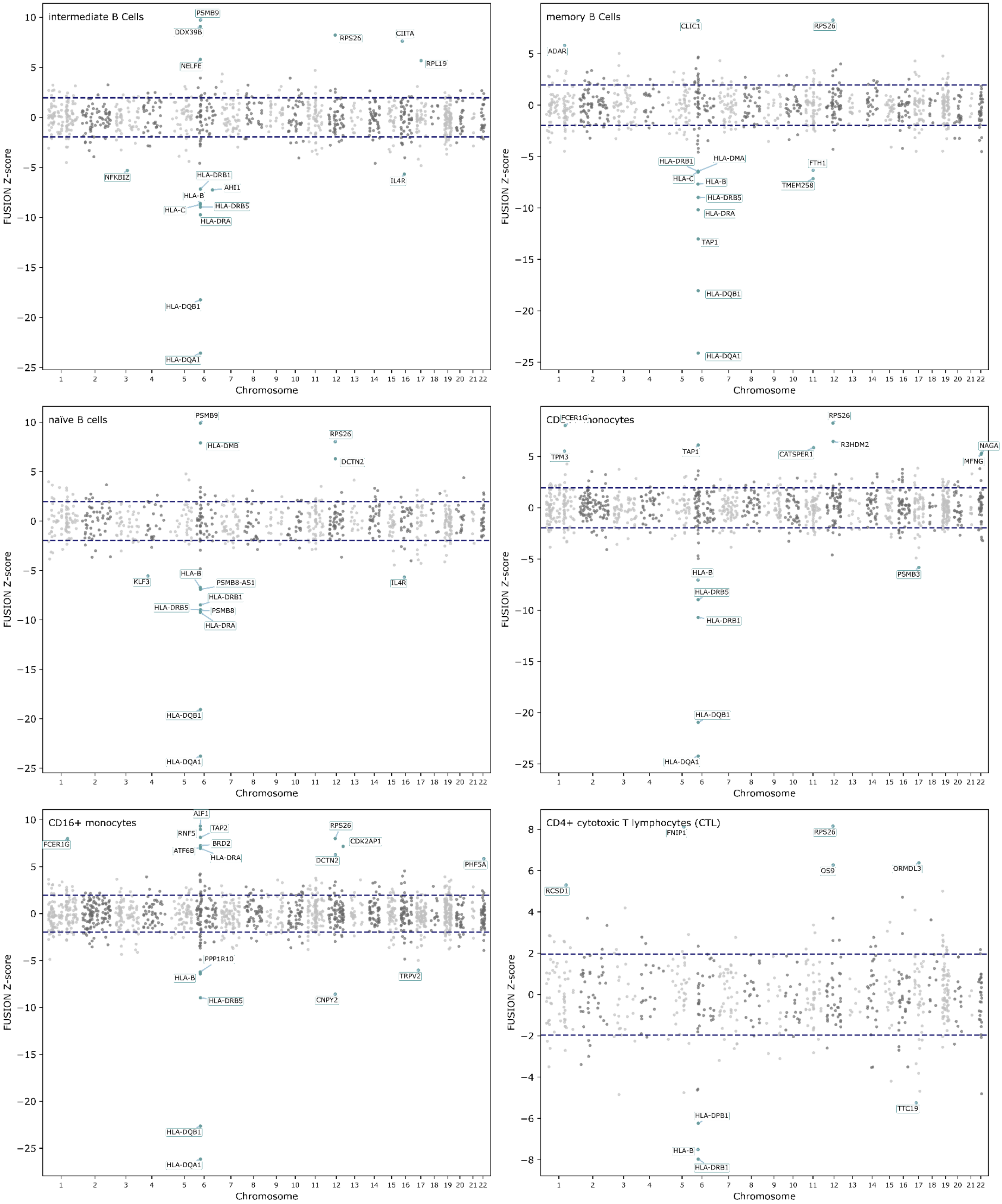

intermediate B Cells
PSMB9
DDX39B
NELFE
RPS26
CIITA
RPL19
NFKBIZ
HLA-DRB1
AHI1
HLA-B
HLA-C
HLA-DRB5
HLA-DRA
IL4R
HLA-DQB1
HLA-DQA1
memory B Cells
ADAR
CLIC1
RPS26
HLA-DRB1
HLA-DMA
HLA-C
HLA-B
HLA-DRB5
HLA-DRA
FTH1
TMEM258
TAP1
HLA-DQB1
HLA-DQA1
naïve B cells
PSMB9
HLA-DMB
RPS26
DCTN2
KLF3
HLA-B
PSMB8-AS1
HLA-DRB1
HLA-DRB5
PSMB8
HLA-DRA
IL4R
HLA-DQB1
HLA-DQA1
monocytes
FCER1G
TPM3
TAP1
CATSPER1
RPS26
R3HDM2
NAGA
MFNG
HLA-B
HLA-DRB5
HLA-DRB1
PSMB3
HLA-DQB1
HLA-DQA1
CD16+ monocytes
FCER1G
AIF1
RNF5
TAP2
BRD2
ATF6B
HLA-DRA
RPS26
CDK2AP1
DCTN2
PHF5A
PPP1R10
HLA-B
TRPV2
HLA-DRB5
CNPY2
HLA-DQB1
HLA-DQA1
CD4+ cytotoxic T lymphocytes (CTL)
FNIP1
RPS26
RCSD1
OS9
ORMDL3
HLA-DPB1
TTC19
HLA-B
HLA-DRB1
FUSION Z-score
Chromosome

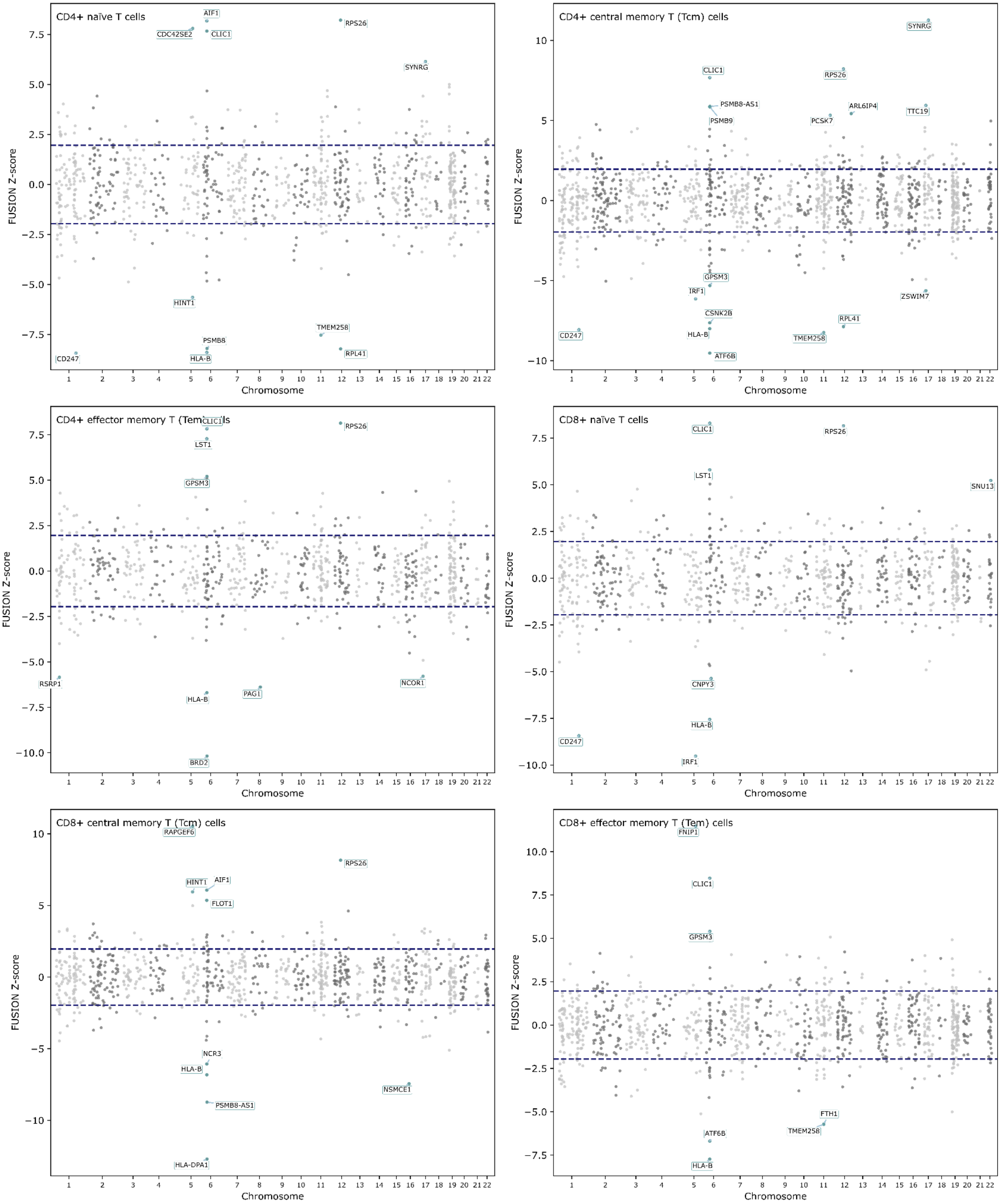
CD4+ naïve T cells
AIF1
CDC42SE2
CLIC1
RPS26
SYNRG
HINT1
TMEM258
PSMB8
CD247
HLA-B
RPL41
FUSION Z-score
Chromosome
CD4+ central memory T (Tcm) cells
SYNRG
CLIC1
RPS26
PSMB8-AS1
PSMB9
ARL6IP4
PCSK7
TTC19
GPSM3
IRF1
ZSWIM7
CSNK2B
RPL41
CD247
HLA-B
TMEM258
ATF6B
CD4+ effector memory T (Tem) cells
CLIC1
RPS26
LST1
GPSM3
RSRP1
NCOR1
HLA-B
PAG1
BRD2
CD8+ naïve T cells
CLIC1
RPS26
LST1
SNU13
CNPY3
HLA-B
CD247
IRF1
CD8+ central memory T (Tcm) cells
RAPGEF6
RPS26
HINT1
AIF1
FLOT1
NCR3
HLA-B
NSMCE1
PSMB8-AS1
HLA-DPA1
CD8+ effector memory T (Tem) cells
FNIP1
CLIC1
GPSM3
FTH1
TMEM258
ATF6B
HLA-B

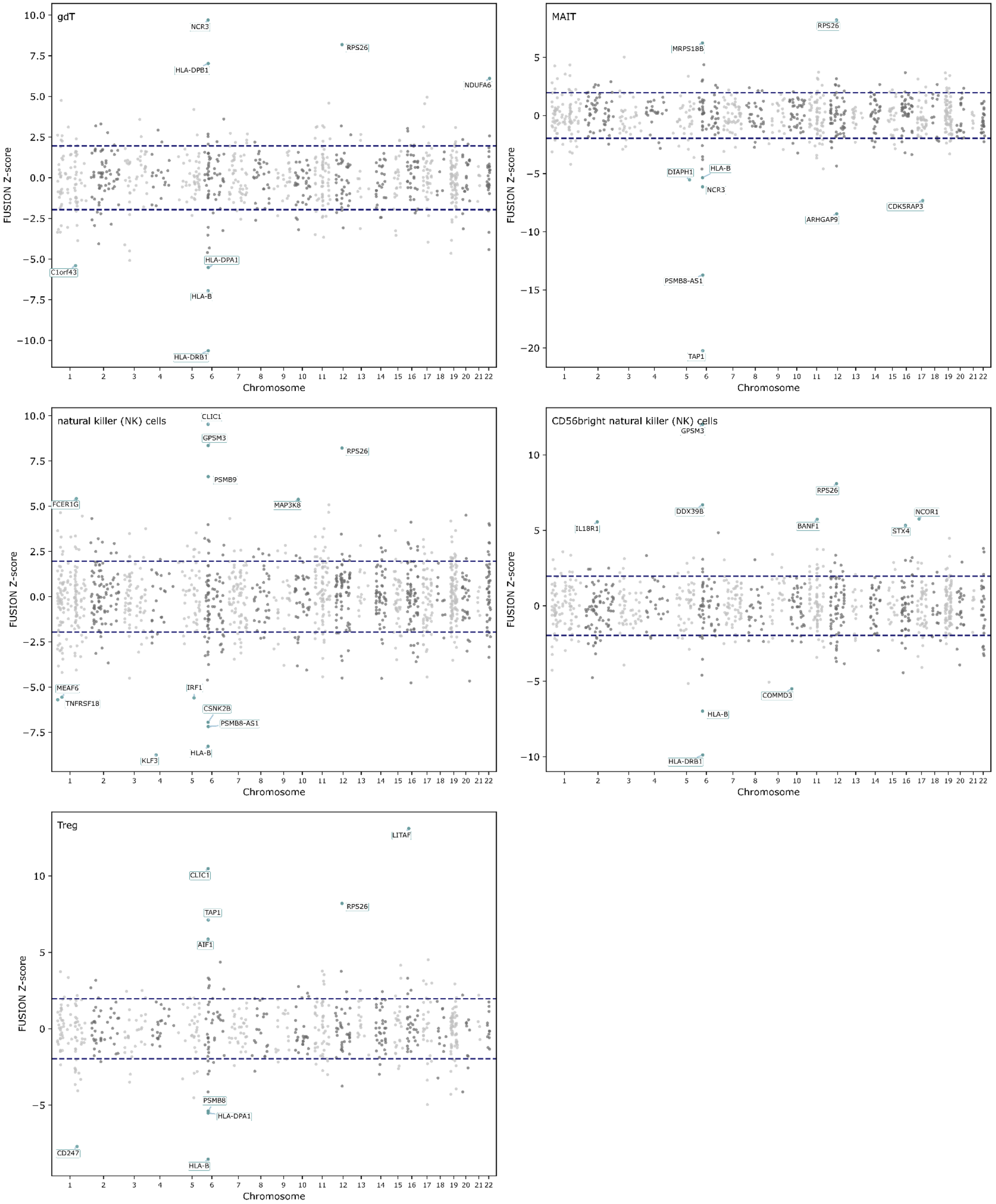
gdT
NCR3
HLA-DPB1
RPS26
NDUFA6
HLA-DPA1
C1orf43
HLA-B
HLA-DRB1
FUSION Z-score
Chromosome
MAIT
RPS26
MRPS18B
DIAPH1
HLA-B
NCR3
ARHGAP9
CDK5RAP3
PSMB8-AS1
TAP1
FUSION Z-score
Chromosome
natural killer (NK) cells
CLIC1
GPSM3
RPS26
PSMB9
FCER1G
MAP3K8
MEAF6
TNFRSF18
IRF1
CSNK2B
PSMB8-AS1
HLA-B
KLF3
FUSION Z-score
Chromosome
CD56bright natural killer (NK) cells
GPSM3
RPS26
DDX39B
IL18R1
BANF1
NCOR1
STX4
COMMD3
HLA-B
HLA-DRB1
FUSION Z-score
Chromosome
Treg
LITAF
CLIC1
RPS26
TAP1
AIF1
PSMB8
HLA-DPA1
CD247
HLA-B
FUSION Z-score
Chromosome

#### Figure S3. FUSION TWAS gene-tissue and gene-cell-type associations tested versus passing nominal filters across bulk GTEx tissues and pseudobulked OneK1K PBMC cell types.

Bar plots of (**A**) the total number of gene-tissue pairs tested (grey) and passing nominal filters (blue) across 39 bulk GTEx Project v8 tissues, and (**B**) the total number of gene-cell-type pairs tested (grey) and passing nominal filters (teal) across 17 pseudobulked cell types derived from peripheral blood mononuclear cells (PBMC) from the OneK1K Project cohort. In both panels, associations passing nominal filters met the following criteria: TWAS $p$-value < 0.05, TWAS Z-score ≠ 0, and SNP-heritability $h^2 > 0$. Tissues and cell types are ordered by the total number of tested pairs in descending order. Nominally significant associations were retained as input for probabilistic fine-mapping with MA-FOCUS and *do not* reflect a multiple-testing-corrected significance threshold.

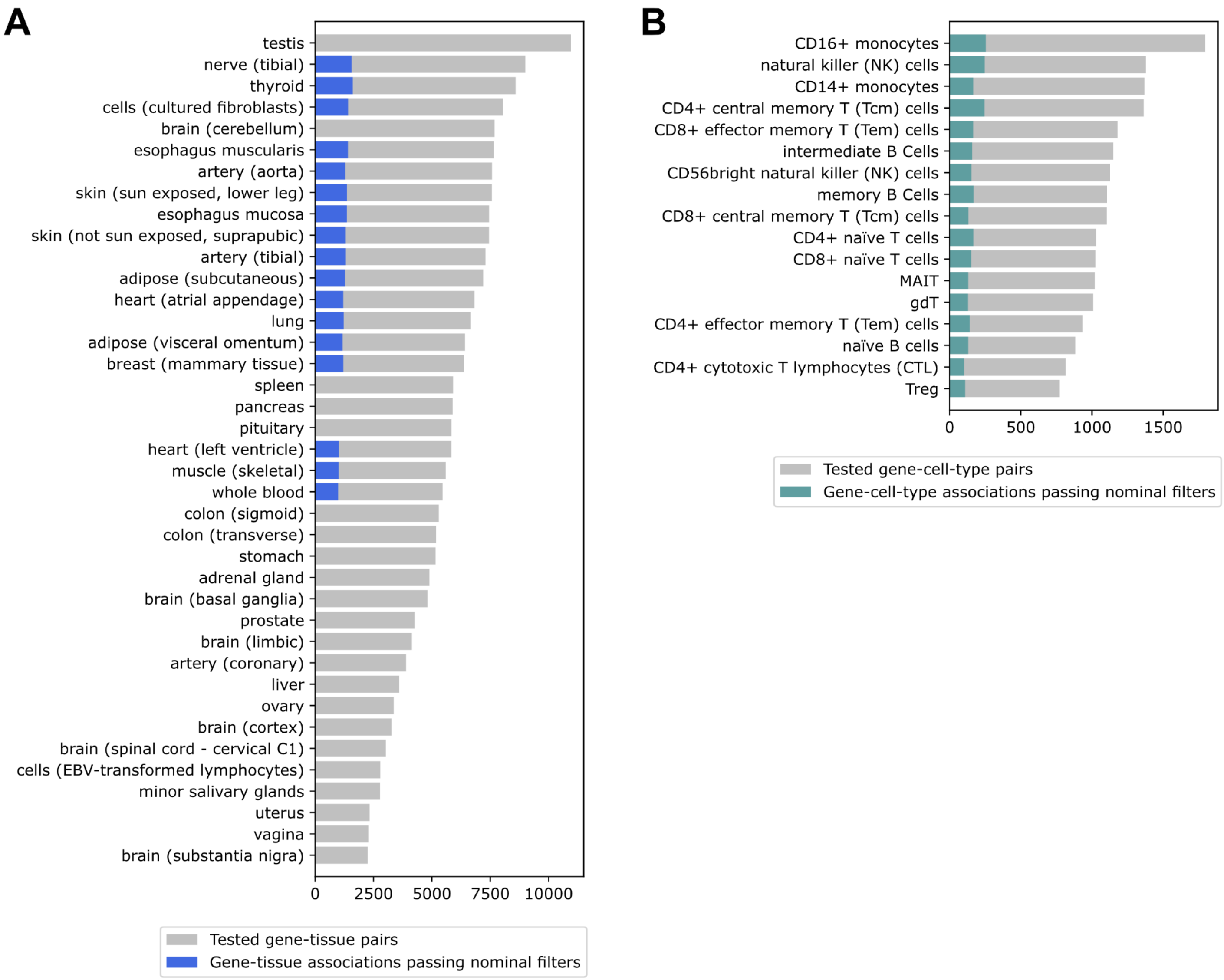
A
testis
nerve (tibial)
thyroid
cells (cultured fibroblasts)
brain (cerebellum)
esophagus muscularis
artery (aorta)
skin (sun exposed, lower leg)
esophagus mucosa
skin (not sun exposed, suprapubic)
artery (tibial)
adipose (subcutaneous)
heart (atrial appendage)
lung
adipose (visceral omentum)
breast (mammary tissue)
spleen
pancreas
pituitary
heart (left ventricle)
muscle (skeletal)
whole blood
colon (sigmoid)
colon (transverse)
stomach
adrenal gland
brain (basal ganglia)
prostate
brain (limbic)
artery (coronary)
liver
ovary
brain (cortex)
brain (spinal cord - cervical C1)
cells (EBV-transformed lymphocytes)
minor salivary glands
uterus
vagina
brain (substantia nigra)
0
2500
5000
7500
10000
Tested gene-tissue pairs
Gene-tissue associations passing nominal filters
B
CD16+ monocytes
natural killer (NK) cells
CD14+ monocytes
CD4+ central memory T (Tcm) cells
CD8+ effector memory T (Tem) cells
intermediate B Cells
CD56bright natural killer (NK) cells
memory B Cells
CD8+ central memory T (Tcm) cells
CD4+ naïve T cells
CD8+ naïve T cells
MAIT
gdT
CD4+ effector memory T (Tem) cells
naïve B cells
CD4+ cytotoxic T lymphocytes (CTL)
Treg
0
500
1000
1500
Tested gene-cell-type pairs
Gene-cell-type associations passing nominal filters

#### Figure S4. GO term enrichment analysis of FDR-significant genes from FUSION TWAS gene-tissue and gene-cell-type associations.

Bar plots of significant Gene Ontology (GO) term enrichment results (False Discovery Rate, FDR < 5%) among (**A**) 3,524 unique genes identified from gene-tissue associations across 39 bulk GTEx Project v8 tissues, and (**B**) 479 unique genes identified from gene-cell-type associations across 17 pseudobulked cell types derived from peripheral blood mononuclear cells (PBMC) from the OneK1K Project cohort. Bar lengths represent the enrichment significance as $-\log_{10}(q\text{-value})$ and fold-enrichment values are displayed to the right of each bar. Terms are grouped by ontology namespace: Biological Process (BP), Cellular Component (CC), or Molecular Function (MF). Only enriched GO terms as determined by GOATOOLS are shown; depleted (i.e., purified) terms are excluded. We defined gene-tissue and gene-cell-type gene sets by applying a 5% FDR threshold to the FUSION TWAS $p$-values separately within each modality. Study gene sets were then evaluated against FUSION TWAS-tested backgrounds (24,866 unique genes for gene-tissue associations; 3,571 unique genes for gene-cell-type associations) with GOATOOLS. Fold-enrichment values are not directly comparable across panels due to differences in background gene set size.

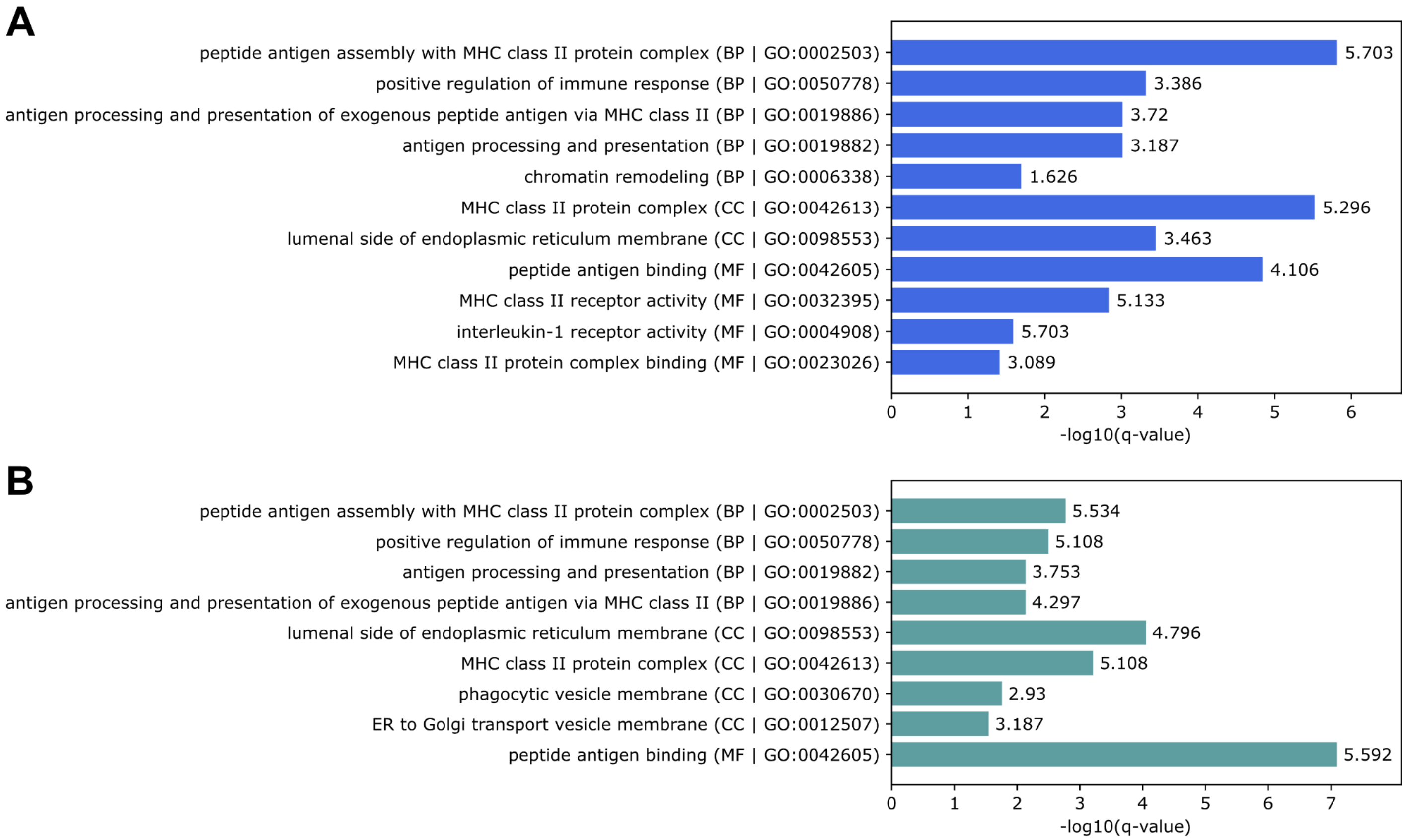

#### Figure S5. Number of genes in the FOCUS 90% credible set per tissue/cell type.

Horizontal bar plots show the number of gene–context pairs included in FOCUS 90% credible sets for each GTEx tissue (**A**) and OneK1K cell type (**B**). Contexts are ordered by decreasing feature count, with the number of retained pairs shown beside each bar. GTEx tissues contained substantially more credible-set pairs than OneK1K cell types, consistent with broader and more diffuse posterior support across correlated bulk-tissue expression models. These raw counts also reflect differences in the number and composition of expression models available in the two reference panels and should not be interpreted as direct measures of biological importance.

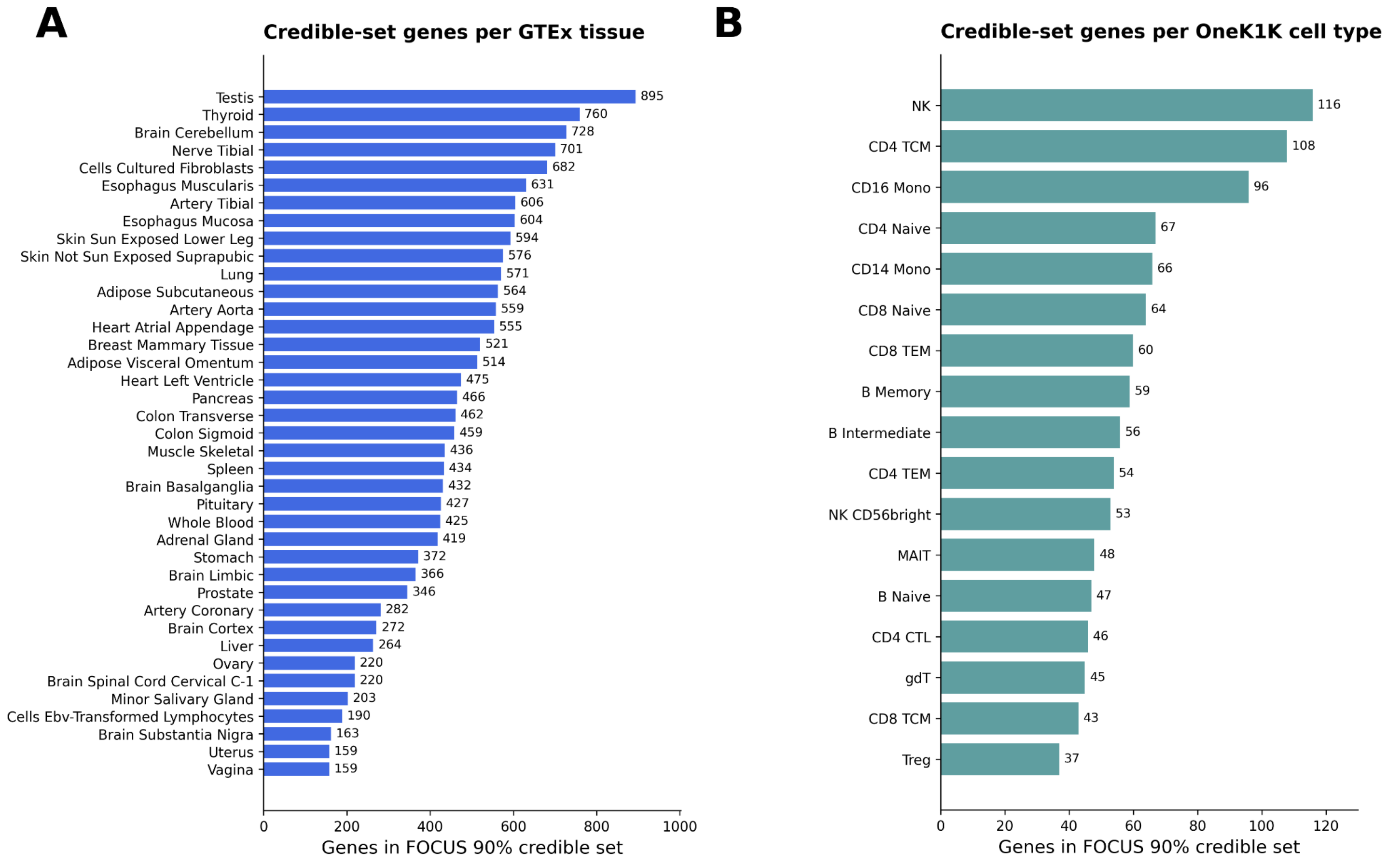

##### Figure S6. FOCUS posterior support is more concentrated in OneK1K cell types than in GTEx tissues.

Stacked bar plots show the distribution of FOCUS posterior inclusion probabilities (PIPs) among gene–context pairs in 90% credible sets across 39 GTEx tissues (left) and 17 OneK1K cell types (right). The x-axis represents the proportion of pairs within each context, and colors indicate PIP intervals. GTEx credible sets were dominated by low-PIP gene–tissue pairs, whereas OneK1K cell types contained larger proportions of intermediate- and high-PIP pairs. This difference indicates more concentrated posterior support in the cell-type-level analysis and broader, more diffuse support across bulk tissues, potentially reflecting greater cross-tissue co-regulation.

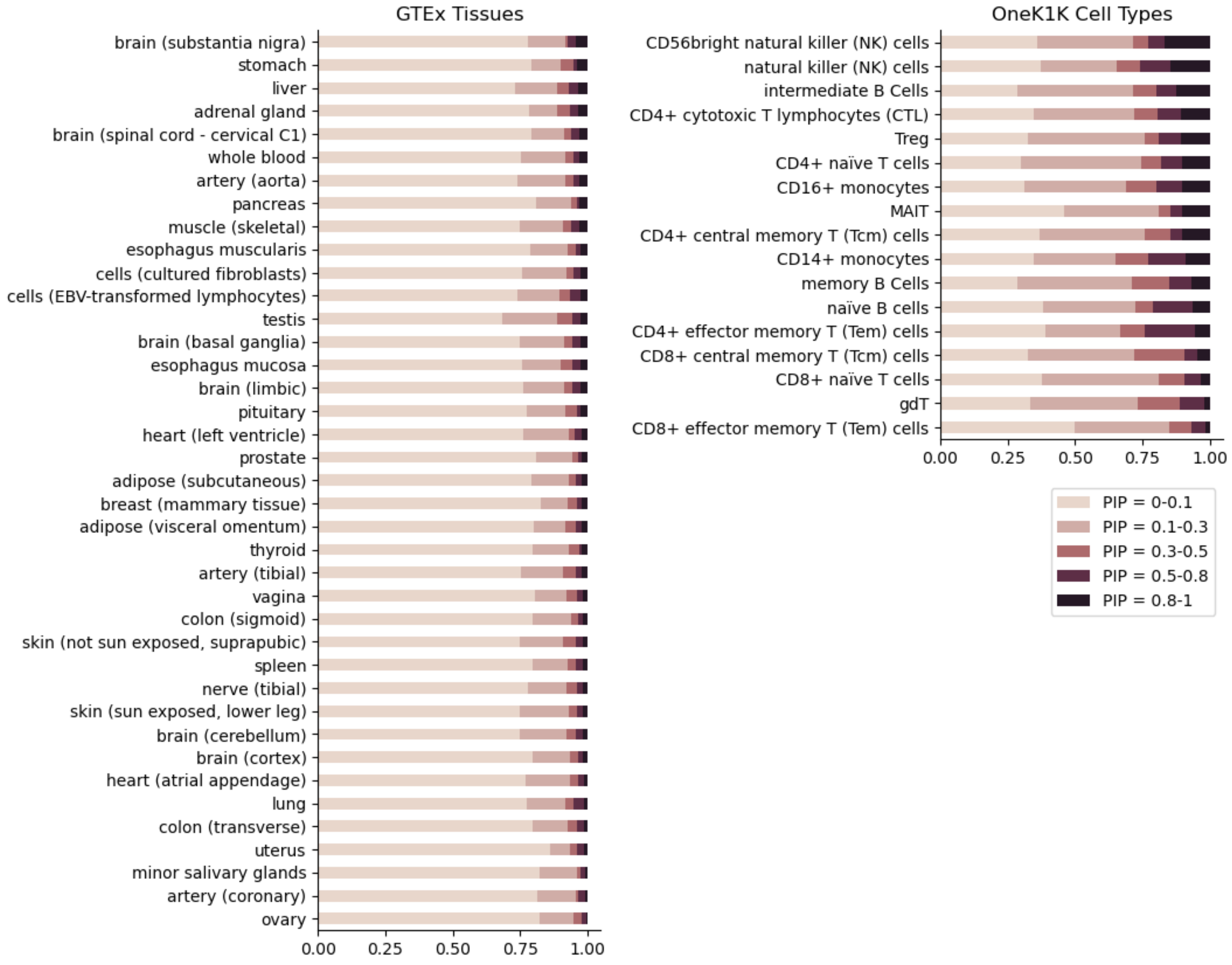
